# FAMEWS: a Fairness Auditing tool for Medical Early-Warning Systems

**DOI:** 10.1101/2024.02.08.24302458

**Authors:** Marine Hoche, Olga Mineeva, Manuel Burger, Alessandro Blasimme, Gunnar Rätsch

**Author notes:** These authors contributed equally (Marine Hoche, Olga Mineeva).

## Abstract

Machine learning applications hold promise to aid clinicians in a wide range of clinical tasks, from diagnosis to prognosis, treatment, and patient monitoring. These potential applications are accompanied by a surge of ethical concerns surrounding the use of Machine Learning (ML) models in healthcare, especially regarding fairness and non-discrimination. While there is an increasing number of regulatory policies to ensure the ethical and safe integration of such systems, the translation from policies to practices remains an open challenge. Algorithmic frameworks, aiming to bridge this gap, should be tailored to the application to enable the translation from fundamental human-right principles into accurate statistical analysis, capturing the inherent complexity and risks associated with the system. In this work, we propose a set of fairness impartial checks especially adapted to ML early-warning systems in the medical context, comprising on top of standard fairness metrics, an analysis of clinical outcomes, and a screening of potential sources of bias in the pipeline. Our analysis is further fortified by the inclusion of event-based and prevalence-corrected metrics, as well as statistical tests to measure biases. Additionally, we emphasize the importance of considering subgroups beyond the conventional demographic attributes. Finally, to facilitate operationalization, we present an open-source tool FAMEWS to generate comprehensive fairness reports. These reports address the diverse needs and interests of the stakeholders involved in integrating ML into medical practice. The use of FAMEWS has the potential to reveal critical insights that might otherwise remain obscured. This can lead to improved model design, which in turn may translate into enhanced health outcomes.

## 1. Introduction

We are witnessing the rise of Machine Learning (ML) models targeting the healthcare domain. The increasing availability of electronic health record (EHR) datasets enables the development of AI-based monitoring systems in the hospital. For instance, Yèche et al. (2021) propose benchmark models for early detection of organ failure based on the HiRID dataset (Faltys et al., 2021). These prognosis early-warning systems aim to raise the alarm in case of a high risk of organ failure within the next 12 hours. These systems are meant to be applied to critically ill patients and could have a tremendous impact on their health outcomes. As with every ML model, these systems can be biased (Coeckelbergh, 2020) and could lead to unfair health disadvantages for some patient groups (Vayena et al., 2018). Governments worldwide have expressed concern about the ethics and safe integration of ML systems. For instance, the proposed EU AI Act^1,2^ aims to answer to the urgency of framing the models with strict regulatory policies. Regarding the fairness of such models, the draft of the act promotes audits of algorithms and datasets to ensure non-discrimination and non-violation of human rights. To this end, they require developers to provide documentation about the model’s general characteristics, capabilities, and limitations. However, no further details are provided on how to audit fairness in practice. As highlighted in the review of algorithmic fairness (Pagano et al., 2023), this task is challenging as there is no consensus on how to measure the fairness of an algorithm.

To fully comprehend the issue of bias in medical ML, we conducted exploratory work with ethics professionals and clinicians analyzing early detection of circulatory failure as developed in the HiRID benchmark (Yèche et al., 2021). In this first attempt (to the best of our knowledge) to design a fairness auditing framework for early-warning systems, we acknowledge the necessity to not only check for classical notions of fairness but also to investigate the fairness of the early-warning system’s real-world consequences (McCradden et al., 2020). We question various system’s design choices from a fairness perspective as bias can be introduced at many stages of the Machine Learning pipeline (Rajkomar et al., 2018). We summarize our learnings in an open-source tool FAMEWS which primarily complements the HiRID benchmarks (Yèche et al., 2021), but is applicable to a wide range of early-warning systems.

Our main contributions are:

1. **A flexible fairness-auditing framework tailored for clinical early-warning systems**. The framework is depicted in Figure 1. In the clinical context, patient grouping based on medical attributes such as admission type, comorbidities, or patient consciousness helps to spot model biases and identify disadvantaged subgroups beyond static demographic attributes (like race or gender). We propose grouping definitions for the HiRID dataset, but the user may change and augment them (Figure 1A). The tool is not restricted to any specific dataset, model type, or prediction task. If lacking some inputs, the user can run only part of the analysis (Figure 1B).
2. **Evaluating ML models, not only through standard metrics but also through comparison of clinical outcomes and screening of the potential sources of bias**. Available analyses are listed in Figure 1C and described in Section 3. We focus on prognosis models estimating future risk and providing early alarms, differing from classification setup by including a time dimension. Differences in timing lead to unfair outcomes as well as discrepancies in alarm’s accuracy. Also, as to capture an event, it is enough to have only one alarm, we need to measure recall from the event point of view (in addition to a conventional timestep-based recall). Medical variables serve as input signals and define prediction targets. Differences in their levels and missingness patterns, even if initially clinically justified, can mislead model selection and obscure fairness measurements. Differences in feature ranking across cohorts can also result in an unrepresentative model, especially while implementing a submodel reduced to the most important features. We address these concerns with the screening stages in the framework.
3. **Proposing the automatic generation of a PDF report that is easily shareable with various stakeholders and comprises the detailed fairness analysis and insightful summaries of each audit stage**. Provisioned stakeholder’s needs and interests are described in Section 4.2. We don’t differentiate between users while generating the report. By including all levels of analysis detail, we aim to ease communication as every stakeholder is viewing the same version of the report, and in addition, we do not hide any potentially critical information. An example of the produced report is given in Appendix D, and the insights derived from it are in Appendix C.

**Figure 1:**
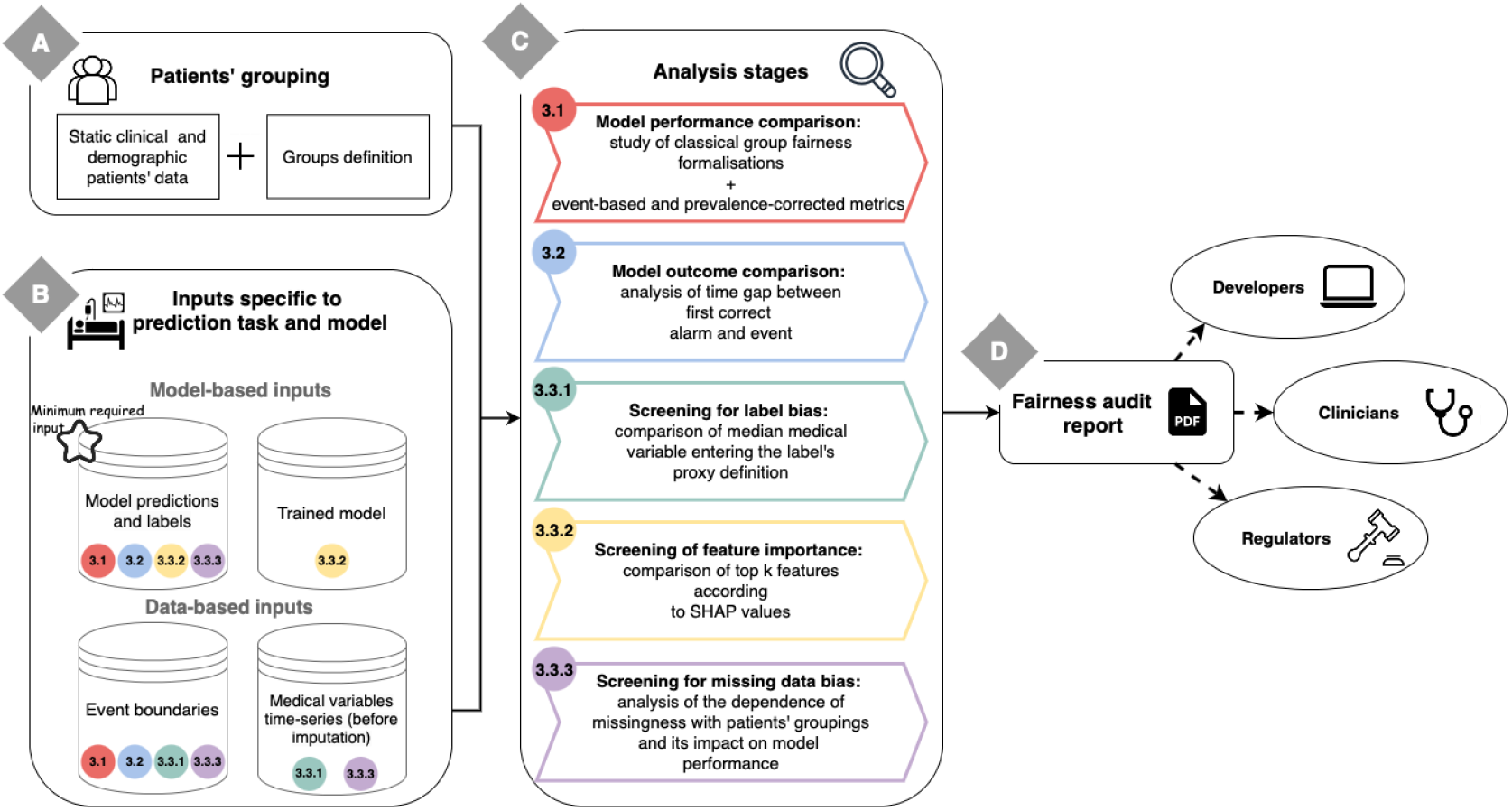
Schema summarising FAMEWS workflow. The user first needs to provide the patients’ groupings (A), which can be based on demographics (like gender or race) or static clinical attributes (like admission reason). Then, for each prediction task and model, the user has to provide model and data-based inputs that are specific to the ML system to audit (B). Afterwards, the different analytical stages can be run (C). Their numbering indicates the corresponding section in the paper. Each analysis stage requires a specific set of inputs depicted in block B by its numbered colored dot. The results of the analyses are gathered in a PDF report that can be shared with the different stakeholders (D).

## 2. Related work

In recent years, with the rise of concern surrounding the fairness of Machine Learning algorithms, tools to detect bias in these models have emerged (Bellamy et al., 2018; Weerts et al., 2023; Cabrera et al., 2019; Wexler et al., 2019; Saleiro et al., 2018; Hertweck et al., 2023). In Table 1, we summarise the characteristics of popular fairness auditing tools and compare them to our framework.

**Table 1:** Comparison of fairness auditing tools

| Characteristic | AI Fairness 360 | Fairlearn | FairVis | What-if Tool | Aequitas | FairnessLab | Our tool |
| --- | --- | --- | --- | --- | --- | --- | --- |
| Other task than binary classification | ✓ | ✓ | ✓ | ✓ | ✗ | ✗ | ✓ |
| Flexible grouping (Not only binary) | ✓ | ✓ | ✓ | ✓ | ✓ | ✗ | ✓ |
| Classical fairness metrics | ✓ | ✓ | ✓ | ✓ | ✓ | ✓ | ✓ |
| Source of bias screening | ✗ | ✗ | ✓ | ✓ | ✗ | ✗ | ✓ |
| Comprehensive fairness assessment | ✗ | ✗ | ✓ | ✗ | ✓ | ✓ | ✓ |
| Robust statistical analysis | ✓ | ✗ | ✗ | ✗ | ✗ | ✗ | ✓ |
| Visual interface | ✗ | ✓ | ✓ | ✓ | ✓ | ✓ | ✓ |

Previous works focus on fair decision-making and as such support binary classifiers. Nonetheless, some of these tools extend to multiclass classifiers or regressors, as shown in the first row of Table 1.

Group fairness can be described as the absence of systematic disadvantages towards a group of individuals that share a common attribute. The type of supported grouping is an important tool characteristic that we outline in Table 1. In the algorithmic fairness literature, classical groupings are based on protected features such as ethnicity, gender, or age and there exists a notion of a privileged and an unprivileged category. We follow the most recent tools and expand this precept by letting the user define their own grouping, which can be multicategorical. FairVis (Cabrera et al., 2019) even proposes to scan the set of possible features to find the most discriminated intersectional group.

In order to assess the fairness of a model, the Machine Learning community relies on formalizations of fairness (Makhlouf et al., 2021). They can be defined as a mathematical condition on the individual’s attributes and the model output, that when satisfied ensures the model’s compliance with a certain vision of fairness. To approximate these formalizations, fairness auditing tools propose to compare common performance metrics from one group to another.

The detection of unfair model outputs opens the question of where the bias is coming from. The source of bias screening is another comparison characteristic in Table 1. In Meng et al. (2022), authors explore how interpretability techniques can be used to grasp the underlying mechanics of detected biases in an ML model. For the same purpose, What-if Tool (Wexler et al., 2019) offers an interactive platform to explore trained models. For instance, they support counterfactual analysis to investigate which attributes have an unjustified effect on the prediction. While What-if tool offers a lot of capabilities to examine the model’s robustness (exploration of feature importance, data distribution, and missingness), it lacks the possibility to perform these analyses per subgroup. Moreover, the counterfactual analysis on protected attributes is quite intricate to perform for medical applications as some of these attributes (such as age and sex) have direct clinically justified impacts on the label.

Finally, a couple of frameworks like FairnessLab (Hertweck et al., 2023) and Aequitas (Saleiro et al., 2018) go beyond the classical bias analysis tools by providing a more comprehensive fairness assessment. They output an intuitive summary with explanations related to relevant ethics and justice concepts, in this way becoming usable by developers as well as regulators and guiding the users to the most adequate fairness metric. For instance, the Aequitas framework (Saleiro et al., 2018) presents an interesting solution for generating fairness reports. It outputs detailed plots to compare different formalizations of fairness across groups as well as summary assessment to easily comprehend for which groups and metrics the model is biased. However, this framework is only suitable for classical binary classification, lacking event-based metrics which are key for evaluating early-warning systems. They also don’t propose outcome-based metrics or screening of potential sources of bias. Moreover, the details about the statistical methodology of their work are missing.

Discussed frameworks are available as libraries and some (Table 1) also embed convenient automatic visualization functionalities like a dashboard or report generation.

Focus on the medical context and early-warning systems differentiate our work from others, that are more general, but missing some essential details for this particular application.

## 3. Tool description

FAMEWS aims to facilitate systematic fairness audits of ML-based alarm systems in the medical field. We designed our tool to widen the usual fairness auditing scope: we assess classical fairness metrics but we also examine the fairness of clinical outcomes and investigate the potential sources of bias. Its main functionalities are summarized in Figure 1.

We consider alarm systems that take as input time-series of medical variables (lab measurements, medications, etc.) and return for each time step a score indicating how likely is the patient to undergo an event within the next X hours.

Our audit is based on comparing key statistics across cohorts of patients. The cohorts can be formed with usual demographics and static clinical information (in Figure 1A). For instance, for the HiRID dataset, the framework includes clinically relevant groupings, such as admission reasons (like trauma or cardiovascular). In the generated PDF report, we display the cohorts’ composition (total number of patients and number of patients undergoing an event). We also give the possibility for the users to filter out cohorts that don’t have enough patients with events (by default this parameter is set to 1), as the analysis would not be statistically significant for them.

An overview of the required inputs for each of the stages is indicated in Figure 1B by a colored dot with a section number. The minimum required input is the model’s predictions and true labels for each timestep. Additionally, the time boundaries of the target events extend the audit to the assessment of performance metrics from the event scope and alarm timing comparison. Access to the trained model (or directly SHAP feature importance values) and the time series dataset allows FAMEWS to run screenings of potential sources of bias.

We recommend providing predictions from models trained with different random seeds, as this will reduce the impact of model randomness on audit results. For each stage of our audit pipeline (in Figure 1C), we run a detailed statistical analysis, that conforms to best practices, and we generate aggregated views to summarize key takeaways. These elements are gathered in a PDF report (Figure 1D). In the following paragraphs, we present the goal and motivation of each analysis stage, the metrics and statistical techniques used to capture disparities between cohorts, and outline generated visualizations and aggregated views for the fairness report.

### 3.1. Classical formalizations of fairness: comparison of model performance across cohorts

#### Goal

In this stage, we compare the model’s performance and the validity of the threshold choice across different patient cohorts through classical fairness notions (Makhlouf et al., 2021; Chen et al., 2023). An example can be found in section 2 of the sample report (Appendix D).

#### Metrics

For each cohort of patients, we compute the metrics related to a set of adequate fairness notions (they are listed in Appendix A together with the definitions of the performance metrics). We implemented binary (recall, precision, FPR, and NPV) and score-based metrics (AUROC, AUPRC, average score on positive and negative classes, calibration error) as they are relevant at different phases of model development. For instance, while tuning the model, score-based metrics are valuable, whereas a deployed model with binary outputs is evaluated using relevant binary metrics. For our targeted medical application, it is beneficial to consider event-based metrics such as event-based recall (number of predicted events over the total number of events) and event-based AUPRC (area under the precision / event-based recall curve). We added the possibility of comparing the precision, NPV, and AUPRC after correction for prevalence (due to the imbalance of positive labels across cohorts). This is equivalent to comparing the original version of these metrics assuming the cohorts have equal prevalence (more details are given in Appendix B).

#### Statistical methodology

We compare the metrics for each cohort to the rest of the patients. To ensure the statistical robustness of this comparison, we first bootstrap the patient population of the test set (we draw with replacement 100 random samples of the test set size) and compute for each cohort in each sample the metrics listed above. We then perform the Mann-Whitney U test with Bonferroni correction. From these statistical tests we obtain for each metric the categories of patients which are significantly worse off compared to the rest of the population. We then quantify these disparities by computing the absolute difference in median metric (taken over the bootstrapped samples) between patients of the category and patients outside it:

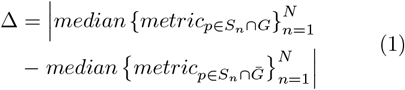

with *S*_*n*_ the *n*^*th*^ bootstrapped sample, *N* the total number of bootstrapped samples drawn, *G* the studied cohort and 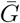 the rest of patients.

#### Visualizations

The results of the comparison are presented as tables in the report. We display box plots for each metric with the median, first quartile, and third quartile over the bootstrapped samples. Cohorts that are significantly worse off are highlighted with a star. For the score-based metrics, we report performance curves: calibration, ROC, and precision-recall (also event-based) curves. The colored error area represents the standard deviation computed over the bootstrap samples. To ease comparison, we keep the same scale for each metric across the entire report.

#### Aggregated views

3 aggregated views are proposed for this stage:

1. Summary statistics for each metric and grouping: it is composed of the macro-average, the minimum over the grouping’s categories, and the metric value for the minority category.
2. Summary view based on the ratio of significantly worse metrics: For each cohort, we report the ratio of significantly worse metrics over the total number of analyzed metrics. We highlight which category of patients within the grouping and across all groupings is the worst in terms of ratio. The largest delta, as defined in Equation (1), for this category is stated.
3. Table displaying for each metric the 3 cohorts with the largest delta that are significantly worse off than the rest of the population. They are also flagged with a red star on the corresponding metric box plot.

### 3.2. Checking for bias of outcomes: comparison of the time gap between first correct alarm and event across cohorts

#### Goal

One outcome of the early-warning system is to direct additional clinical attention to specific patients to prevent the forecasted events. We analyze whether the alarm is triggered sufficiently in advance for the different cohorts of patients. An example is in section 3 of the sample report (Appendix D).

#### Metrics

For each detected event, we compute the time gap between the first correct alarm and the event. The bigger the time gap the better off a patient is.

To not bias this analysis, we first split the events with respect to how much time in advance the alarm could be triggered. For the sake of clarity, let us consider an alarm system with a 12-hour horizon. If an event happens three hours after the start of the stay, the alarm can be triggered at most 3 hours in advance; while if it occurs after 24 hours, the alarm can be raised 12 hours in advance. It is thus not equitable to compare these two categories of events. To overcome this issue, we propose to split the possible alarm window into 4 (configurable) parts: 0-3h, 3-6h, 6-12h, and more than 12h. For each of our alarm window splits and cohort of patients, we then compute the median time gap.

#### Statistical methodology

We draw 100 bootstrap samples (as for the previous stage in Section 3.1). For each bootstrapped sample, each alarm window split, and each cohort of patients, we compute the median time gap. We then use the Mann-Whitney U test with Bonferroni correction to determine which cohorts are significantly worse off than the rest of the population. We quantify the disparity by computing the difference between the median (taken over the bootstrapped samples) time gap for patients belonging to a cohort and patients not belonging to it, for each window split. This is equivalent to computing Δ in Equation (1) with *metric* being the median time gap for the events falling into a specific window split for a selected cohort.

#### Visualizations

The comparison results are outlined in tables and visually displayed in box plots, in the same fashion as for our first analysis (Section 3.1).

#### Aggregated views

2 aggregated views are proposed for this stage:

1. Summary statistics for each alarm window split and grouping of patients composed of the macro-average,the minimum metric value over all the grouping’s categories, and the value for the minority category.
2. Table displaying for each alarm window split the 3 cohorts with the biggest delta that are significantly worse-off than the rest of the population. These cohorts are also flagged with a red star on the corresponding box plot.

### 3.3. Assessing level of bias for potential sources

#### 3.3.1. Comparison of some medical variables across cohorts

##### Goal

It is quite common in clinical contexts to rely on proxy labels instead of ground truth to depict a medical phenomenon. For instance, circulatory failure can be defined through arterial lactate and blood pressure levels. This analysis has been specially designed to tackle the problem of label bias (Wick et al., 2019; Rateike et al., 2022) that can occur in these settings. We want to check whether the proxy used to define the label is correct for all cohorts. An ill-defined label can create degradation in performance and unfair outcomes. We thus propose to compare the distribution of medical variables used in the proxy definition across the different cohorts of patients. Nonetheless, this stage can also be used to study other time-series variables that are relevant to the user. An example can be found in section 4 of the sample report (Appendix D).

##### Metrics

For each cohort, we compare the distribution of chosen medical variables to the rest of the population. For each patient, we compute the median value over the entire stay. According to this stage’s goal, we expect that undergoing an event has a strong influence on the variable value. We thus also inspect separately periods of stay free of events and patients without events.

##### Statistical methodology

We draw 100 bootstrap samples from the train set in the same fashion as in Section 3.1. For each sample and each cohort, we end up with three different median values (for all data points, not during events, and for patients free of events) for the selected medical variables. We compare the distribution of each median from one cohort to the rest of the population using the Mann-Whitney U test with Bonferroni correction. We quantify the difference in median values by computing the absolute difference in medians (median taken over the bootstrapped samples of the different medians) between patients belonging to a cohort and patients not belonging to it. This is equivalent to computing Δ in Equation (1) with *metric* being one of the three median values for a medical variable and a selected cohort.

##### Visualizations

We report the results in tables and with box plots. The star on these plots flags the categories of patients with a significantly different median value compared to the rest of the patients.

##### Aggregated views

We outline, for each of the selected medical variables and the median computation methods, the 3 cohorts with the biggest delta in median value that are significantly different from the rest of the population. These cohorts are also signaled with a red star on the corresponding variable box plot.

#### 3.3.2. Comparing the top k features across cohorts

##### Goal

Regarding explainability concerns, it is essential for the stakeholders to know the features that drive the prediction process. We check whether feature importance deviates across patient cohorts. We consider this to be of special interest for two scenarios. First, while considering a submodel developers usually keep only the most important features from the validation set (Hyland et al., 2020), however in this process, they can disregard features that are important to minority cohorts, losing predictive power for them (Zong et al., 2023). Then, to check the clinical relevance of the model, it can be useful to show medical practitioners, not only the global top features but also the top features for the different subcohorts. Indeed they might want to review how the medical variables impact the model prediction depending on the various patient profiles. An example is in section 5 of the sample report (Appendix D).

##### Metrics

To study the feature importance, we will rely on SHAP values (Lundberg and Lee, 2017). This is a local explanation method, allowing us to obtain the feature importance for each data point. We can thus obtain the feature importance for each patient and aggregate them per cohort. Furthermore, this method aligns better with human intuition than other feature importance estimation techniques (Lundberg and Lee, 2017), such as LIME (Ribeiro et al., 2016). Nonetheless, this framework can yield inaccurate feature importance values when features are dependent or correlated. (Aas et al., 2021).

For each patient and a given feature, we thus quantify its importance with the absolute mean SHAP value over the stay. Then we derive a feature ranking for a cohort based on the mean feature importance over all of its patients. We compare the feature ranking of each cohort to the global feature ranking using a similarity measure on lists called the rank-biased overlap (RBO) (Webber et al., 2010). This measure has the particularity of giving more weight to the head compared to the tail (weighting parameter *p* = 0.935). This aspect is particularly suitable to the comparison of feature importance rankings as we care more about differences for the top features (Sarica et al., 2022). Nonetheless, this property highly depends on the weighting parameter, which can be challenging to tweak properly. For each feature ranking, we flag the features that significantly changed rank compared to the global ranking.

##### Statistical methodology

To establish the statistical relevance of our analysis, we compute the RBO for feature ranking on random simulated patient cohorts. This yields an upper bound,

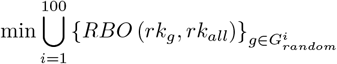

(with *G*^*i*^ the *i*^*th*^ random grouping, *rk*_*g*_ the ranking obtained on one cohort of *G*^*i*^ and *rk*_*all*_ the overall ranking) below which the RBO testifies of significantly different feature rankings. From these random groupings, we compute for each feature, the delta of inverse rank 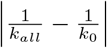 (with *k*_*all*_ the global rank and *k*_0_ the rank we want to compare to) and obtain a lower bound,

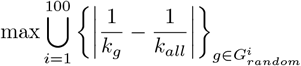

(with *G*^*i*^ the *i*^*th*^ random grouping, *k*_*g*_ the rank of the studied feature for one cohort of *G*^*i*^ and *k*_*all*_ its global rank) above which the delta of inverse rank indicates that the feature has a significantly different rank compared to the global ranking.

##### Visualizations

For each cohort, we outline the top *k* features, we print the feature name in red when it isn’t part of the global top *k* ranking and in blue when it changes rank within the top *k* ranking from global to cohort-based. We only color the names when the change of rank is significant. However, for each feature that changes rank, we put in parenthesis the difference in rank and the direction of change.

##### Aggregated views

We display the RBO for each cohort, colored in red when it is significantly low.

#### 3.3.3. Comparing the missingness of key medical variables and its impact across cohorts

##### Goal

The intensity of measurement of medical variables highly depends on their nature and the health status of the patient. As such, data used for medical applications aren’t missing at random. We thus investigate how the intensity of measurement for relevant variables correlates with patients’ attributes. From a fairness perspective, we can wonder whether disparities in the intensity of measurement across cohorts of patients are purely motivated by medical reasons or whether some forms of discrimination are present. We thus inspect the impact of missingness on the model performance (Getzen et al., 2023). The results could hint at adapting the data collection or the imputation practices. An example can be found in section 6 of the sample report (Appendix D).

##### Metrics

For this analysis, the user needs to provide, for each patient, the time series of medical variables resampled on a fixed time-step grid before data imputation. For each of the selected medical variables, we forward propagate the measurement value according to its usual sampling interval (that has been indicated by the user).

First, we measure the intensity of measurements *I* for each patient that has at least one valid value:

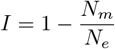

with *N*_*e*_ the number of expected measurements and *N*_*m*_ the number of missed measurements. *N*_*e*_ is defined as 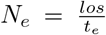 with *los* the patient’s length of stay and *t*_*e*_ the expected sampling interval. *N*_*m*_ is obtained by summing the number of measurements that could have been done during each period 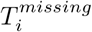 without valid measurements (even after propagation): 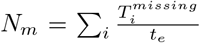. The user provides categorization for the intensity of measurement values. For our example report, we class values below 90% as *insufficient* and above as *enough*. We put apart patients without any measurement. Then, we assess the impact of missing values on performance. The methodology is similar to the stage in Section 3.1: we measure classical metrics but instead of grouping the data points per cohort of patients, we group them based on their missingness status. Data points without valid value after propagation are grouped in the *missing_msrt* category, those belonging to patients without measurement in *no_msrt* and the rest in *with_msrt*. For this analysis, we don’t measure event-based metrics. For variables used in the label’s definition, it is not possible to run the analysis on the *no_msrt* category.

##### Statistical methodology

We run the Chi-squared independence test to assess the dependence between the patients’ grouping and the intensity of measurement categories.

The statistical tests for the impact of performance analysis are run in the same fashion as in Section 3.1. However, instead of comparing each cohort to the rest of the population, we compare the missingness categories *no_msrt* and *missing_msrt* against the *with_msrt* category.

##### Visualizations

For the intensity of measurements analysis, we provide for each cohort a bar plot displaying the percentage of patients belonging to each intensity category. The dotted lines show the percentage over the entire population of patients as references. For the impact on performance, we present the results in tables and box plots as in Section 3.1.

##### Aggregated views

For each of the selected medical variables, if the grouping and the intensity of measurements are dependent, the grouping is outlined in a table. Also, the category for the corresponding grouping with the biggest rate of patients without measurement and the one with an insufficient number of measurements are indicated. To summarize the impact on the performance, the ratio of metrics that are significantly worse than the *with_msrt* metrics is displayed for each missingness category as well as the worst delta in metrics.

## 4. Discussion

In this paper, we described FAMEWS – a fairness auditing tool tailored for medical early-warning systems. Our approach extends the scope of classical fairness assessment tools by including an analysis of fairness of outcomes, screening of potential sources of bias, and proposing to consider clinical attributes on top of classical demographic features for fairness analysis. We will now discuss the flexibility of our tool, how our generated report can be used by the different stakeholders as well as the strengths and limitations of our work.

### 4.1. Flexibility of the tool

We primarily built our tool to audit the fairness of an LGBM (Light Gradient-Boosting Machine) early-warning system detecting circulatory failures in the intensive care unit on the HiRID dataset (Yèche et al., 2021). Nonetheless, we conceived it with a certain level of flexibility, allowing it to be extended to a broader range of applications. We tested our framework on other alarm systems (early detection of respiratory failure (Hüser et al., 2024)) with different alarm-to-event horizon lengths, on other datasets like MIMIC-III (Johnson et al., 2016), and on other types of models (Long Short-Term Memory networks). The users can define their own patients’ groupings depending on the available attributes, provide processed inputs rather than raw data, or run only a subset of the stages if they don’t have access to some input data. Moreover, some stages can be adapted to audit other types of binary classifiers; for instance, where the model outputs for each patient a single prediction instead of a time series. Finally, our tool is open-source, offering the possibility to the users to further extend its functionalities.

However, to complete this fairness audit, the user needs a minima access to some test data and the capability to generate predictions from the model (see Figure 1B).

### 4.2. Intended use of the produced report

We designed our report as a conveniently exchangeable document that can be understood and used by different stakeholders. We decided against an interactive dashboard that, although more convenient for exploratory work, would have required the technical skills of the end user, secured access to medical data, and would not have been easily exportable. We now list the provisioned use of the report for the identified stakeholders:

#### Developers

- Compare different model design choices (model type, preprocessing, feature engineering) in order to choose the best model from a fairness point of view. A quick glimpse of how the model is evolving can be obtained by comparing the aggregated views of the respective reports.
- Identify targets for bias mitigation and measure the impacts of different debiasing methods. The aggregated views can be used to facilitate model comparison and choose the best bias mitigation.
- Monitor the behavior of the model, from a fairness point of view, while using the model on new data samples or retraining it (after the deployment for instance).

#### Clinicians

- Adapt their reliance on the model by learning about its main biases, which are highlighted in the aggregated views. For instance, if the practitioners are aware that the model is performing worse for a specific patient cohort then they will not overly rely on the model to monitor these patients, avoiding falling into an automation bias (Rajkomar et al., 2018).
- Provide developers feedback and help them to comprehend certain disparities, especially in the screening of sources of bias analyses. For instance, the results of the label bias screening can be used to discuss the validity of the label proxy definition for all patients. Their feedback can then guide the developers in choosing adequate bias mitigation techniques.

#### Regulators

- Get informed about the model limitations in terms of bias and obtain a brief overview of the demographics.
- Check that the model complies with actual regulations in terms of fairness and non-discrimination.

### 4.3. Strengths and limitations of our framework

The resulting audit report might seem cumbersome to apprehend. We nonetheless believe it is necessary to present the entire analysis in the report, as selecting relevant results is subjective and might hide relevant disparities to the end users. We facilitate its navigation with a table of contents, a glossary, and aggregated views for each analysis stage. These views help in grasping the main takeaways of the report. However, like every summary, it is not self-sufficient and we insist on the necessity to refer to the more detailed analyses to fully understand the extent of potential biases.

Despite its size, our report is rather limited in the range of screened sources of bias. We tackle the ones that we deem crucial for our prime use case. However, depending on the system’s design choices, other sources are also valuable to explore. We acknowledge similar limitations on our exploration of bias of outcomes. Indeed, this issue is deeply dependent on the application and some are not measurable without access to the actual real-world consequences of the ML system. We thus encourage the users to extend the fairness audit to the inspection of post-deployment biases. Then, our tool proposes a limited set of fairness metrics, contrary to other tools. Nonetheless, we implemented evaluation with event-based metrics and prevalence correction which we didn’t find in other fairness auditing tools, but we consider them important for early-warning systems auditing. Finally, we enforced best statistical practices to bring an adequate level of robustness to our audit results. We realized that this aspect was missing in existing fairness analysis frameworks.

In summary, we propose FAMEWS to assess the fairness of ML-based early-warning systems. We believe that the wide adoption of such auditing tools could ease the communication between regulators, developers, and clinicians and could assist in developing both accurate and ethical applications.

## Data Availability

In this study, we primarily experiment with HIRID dataset, which is available for download on PhysioNet: https://physionet.org/content/hirid/1.1.1/, and with the benchmark models for early-detection of organ failure whose code base is available at https://github.com/ratschlab/HIRID-ICU-Benchmark. The FAMEWS open-source tool is available at: https://github.com/ratschlab/famews.

## Data and Code Availability

In this study, we primarily experiment with HIRID dataset (Faltys et al., 2021), which is publicly available for download on PhysioNet (Goldberger et al., 2000), and with the benchmark models for early-detection of organ failure developed by Yèche et al. (2021) whose code base is available at https://github.com/ratschlab/HIRID-ICU-Benchmark/.

The FAMEWS open-source tool is available at: https://github.com/ratschlab/famews.

## Institutional Review Board (IRB)

The institutional review board (IRB) of the Canton of Bern approved the study on retrospective ICU (BASEC 2016 01463). The need for obtaining informed patient consent for patient data from our institution was waived owing to the retrospective and observational nature of the study.

## Authors’ contributions

M.H. conceptualized the study, defined the methodology, developed the tool, conducted all the computational experiments, interpreted the results and prepared the manuscript.

O.M. conceptualized the study, defined the methodology, assisted in interpreting the results, reviewed the tool’s code base, prepared the manuscript, provided supervision and feedback and coordinated the project.

M.B. assisted in data preprocessing, provided the technical support, created the Python package and reviewed the manuscript.

A.B. provided supervision and feedback and reviewed the manuscript.

G.R. conceptualized the study, secured funding, provided supervision, provided technical and conceptual feedback, and resources, reviewed the manuscript.

## 5. Acknowledgments

This project was supported by grant #2022-278 of the Strategic Focus Area “Personalized Health and Related Technologies (PHRT)” of the ETH Domain (Swiss Federal Institutes of Technology), and ETH core funding (to GR). OM is also supported by the Max Planck ETH Center for Learning Systems. We acknowledge discussions with David Berger, Martin Faltys and Effy Vayena. Computational analyses were performed at the LeonhardMed Trusted Research Environment at ETH Zurich (https://sis.id.ethz.ch/services/sensitiveresearchdata/).

## Appendix A. Formalizations of fairness and performance metrics

In this section, we define in Table 2 the different performance metrics available in FAMEWS. We show in Table 3 the formalizations of fairness that we thought important to consider while auditing alarm systems in clinical settings and we link them to their corresponding performance metrics. Precision-recall curve and AUPRC aren’t present in this table, as checking together for equal precision and recall across cohorts doesn’t match one of the conventional notions of fairness. Nonetheless, we still include them in our audit pipeline as they are valuable performance metrics for our use-case.

**Table 2:**
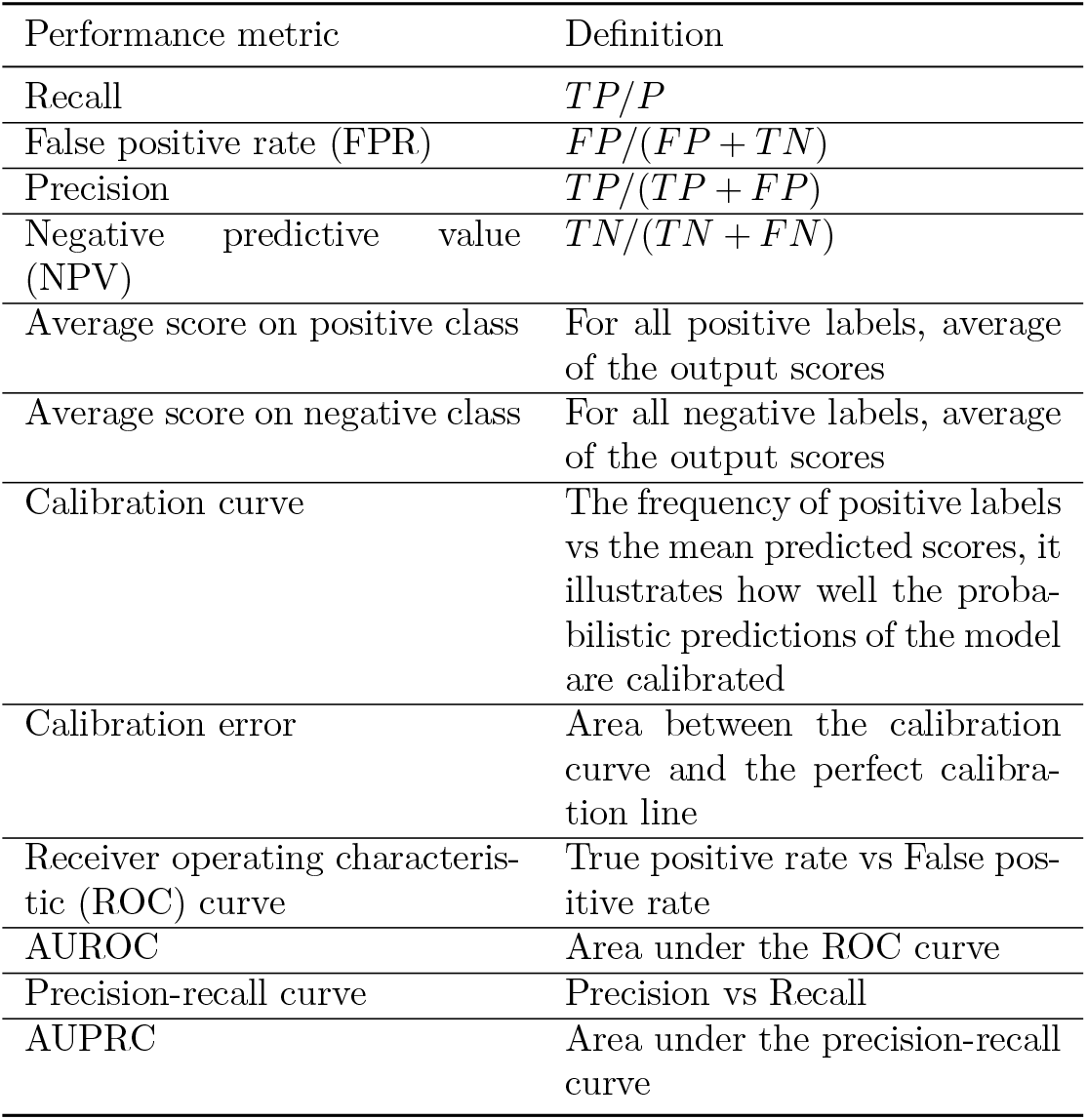
Performance metrics definitions. Definition of each of the model’s performance metrics used in the first step of our fairness analysis. In the formulas, *P* stands for the number of positive labels, *TP* the number of correctly predicted positive labels, *TN* the number of correctly predicted negative labels, *FP* the number of instances with true negative labels that were incorrectly predicted as positive by the model, and *FN* the number of instances with true positive labels that were incorrectly predicted as negative by the model.

**Table 3:** Relation between popular formalizations of fairness and performance metrics. We selected a set of formalizations of fairness that we deemed relevant for our use-case. In this table, we outline for each formalization the corresponding metrics we inspected. We consider that a notion of fairness is respected when the corresponding metric is equal across cohorts. When we use the symbol ‘&’ that means that both metrics have to be equal. For curves, we inspect visually whether they are similar across cohorts and use their respective error metrics to assess more precisely the disparities.

| Formalisation of fairness | Related performance metric |
| --- | --- |
| Equality of opportunity | Recall |
| Predictive equality | FPR |
| Equalized odds | AUROC, ROC curve, recall & FPR |
| Predictive parity | Precision |
| Conditional use accuracy | NPV & precision |
| Balance on positive class | Average score on positive class |
| Balance on negative class | Average score on negative class |
| Calibration | Calibration curve, calibration error |

## Appendix B. Proof prevalence correction

Consider *C* to be a random binary classifier. It assigns class 0 and class 1 with equal probability. Let 𝒟_1_ and 𝒟_2_ be two datasets with different prevalence *pv*_1_ and *pv*_2_, w.l.o.g. we assume *pv*_1_ *< pv*_2_.

This classifier being random, we expect it to have the same performance on 𝒟_1_ and 𝒟_2_. Let us express the recall, FPR, precision, and NPV on both datasets.

We denote by *P*_1_ (resp. *P*_2_) the number of positive labels in 𝒟_1_ (resp. 𝒟_2_), *N*_1_ (resp. *N*_2_) the number of negative labels in 𝒟_1_ (resp. 𝒟_2_), *TP*_1_ (resp. *TP*_2_) the number of correctly predicted positive labels in 𝒟_1_ (resp. 𝒟_2_), *TN*_1_ (resp. *TN*_2_) the number of correctly predicted negative labels in 𝒟_1_ (resp. 𝒟_2_), *FP*_1_ (resp. *FP*_2_) the number of negative labels wrongly predicted as positives in 𝒟_1_ (resp. 𝒟_2_) and *FN*_1_ (resp. *FN*_2_) the number of positive labels wrongly predicted as negatives in 𝒟_1_ (resp. 𝒟_2_).

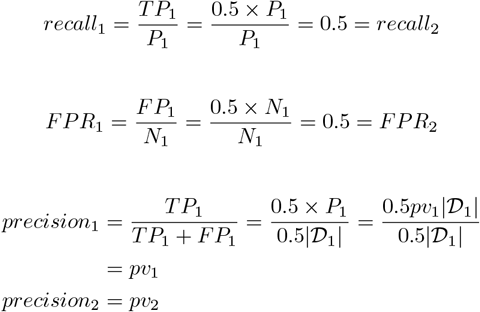

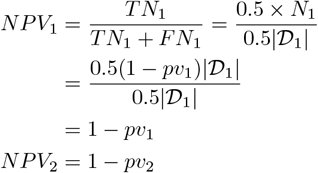

Recall and FPR are equal for both datasets as expected. However, this is not the case for precision and NPV. Let us find a way to modify the formula of precision and NPV such that they are equal for both datasets.

### Correction of precision

We want *c_precision*_1_ = *c_precision*_2_ (with *c_precision* the corrected precision.) We keep the higher prevalence *pv*_2_ as a reference and we want to correct for *pv*_1_. We denote by *s* the correction factor. We will artificially modify the number of false positives for 𝒟_1_ by the factor *s*.

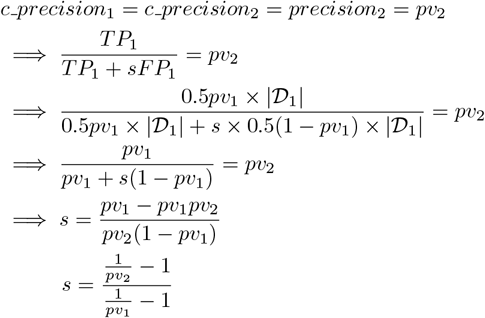

### Correction of NPV

We want *c_NPV*_1_ = *c_NPV*_2_ (with *c_NPV* the corrected NPV). We keep the smaller prevalence *pv*_1_ as a reference and we want to correct for *pv*_2_. We denote by *s* the correction factor. We will artificially modify the number of false negatives for 𝒟_1_ by the factor *s*.

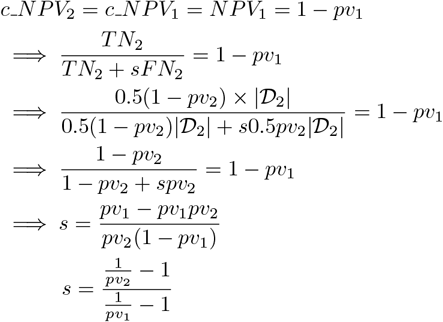

This correction allows us to have the same precision and NPV for both datasets. It is equivalent to considering the precision and NPV in the case the prevalences of both datasets are equal. All stages have be run on the test set, except for the missingness analysis that have been run of the training set.

## Appendix C. Main findings from the example report

We will now outline the key takeaways from the fairness audit of the circulatory failure early-warning system (Yèche et al., 2021) that we infer from the sample report (Appendix D). This report was obtained by running FAMEWS on the averaged predictions from 10 LGBM models trained with different random seeds on the HiRID dataset. It can serve as an example of how to interpret such an analysis account.

### C.1. Systematic performance discrepancy for male patients

In the summary table **Summarized performance metrics per grouping** (2.1.1.a), we can notice that for almost every metric (except one) the model performs worse on male patients than on female patients. Moreover, in the next aggregated view, it is highlighted that an important part of these metrics is statistically significantly worse. However, looking at the more detailed analysis grouping by sex (section 2.2.1), we realized that the discrepancy in performance (delta value) seems relatively small. The feature ranking doesn’t vary significantly between females and males.

### C.2. Minority categories aren’t always worse off

In the summary tables (from 2.1.1.a to 2.1.1.d) **Summarized performance metrics per grouping**, we can notice that the worst-performing category rarely aligns with the minority category.

### C.3. The effect of prevalence correction

If a cohort has a higher prevalence than the others then its performance is decreased by the prevalence correction, while if it has a lower prevalence its performance will be pushed. Thus, it is not surprising to observe that the gap between female and male patients is increased after the correction of AUPRC (Figure 2.2.1.a). In contrast to the effect on neurological patients, where the performance discrepancy in AUPRC has vanished after the correction, as the prevalence of events is the lowest for the neurological cohort (Figure 2.2.3.a). However, one can wonder whether it makes sense to correct for prevalence, i.e. whether we should compare these cohorts under the assumption that they have similar prevalences. It is then important to discuss with clinicians to gain an understanding of how a specific patient attribute impacts the prevalence.

### C.4. Label bias for neurological patients

In the **Summary view based on the ratio of significantly worst metrics** (subsection 2.1.1), it is underlined that the worst performance discrepancy over the entire set of cohorts is for neurological patients on event-based recall. They also appear a lot in the table **Top 3 categories with biggest performance metric discrepancies** (2.1.3.a), emphasizing that the model is biased against them.

This is also reflected in the bias of outcomes analysis where neurological patients have, by far, the biggest disparity in the time gap between correct alarm and event (section 3).

The **Medical variable analysis** (section 4) can hint at an explanation for these discrepancies. Indeed, neurological patients have a much higher median value for mean arterial pressure (MAP) than other cohorts (see subsection 4.2.3). This variable is used to construct the label for circulatory failure. We can then wonder whether the label definition is correct for these patients. These results trigger discussions with clinicians in order to adapt the model design and use for neurological patients.

### C.5. Dependence of the intensity of measurements on patients’ cohorts

We run the **Missingness analysis** (section 6) for arterial lactate (*a_Lac*) and peak inspiratory pressure (*Spitzendruck*). For both of these medical variables, the intensity of measurements is dependent on the patients’ groupings, both demographic and clinical. Recall which is a critical metric for our type of application, since we don’t want to miss a patient in circulatory failure, is significantly worse when the measurement is missing and the delta values seem quite important. This suggests that missingness has a critical impact on model performance. This sparks processes to improve the imputation strategy and also to dialogue with clinicians in order to gain a better understanding of these patterns of missingness.

## Appendix D. Example of the report

In the following sample report, *APACHE group* refers to the admission reason. To understand the meaning of the medical variables, please refer to the data description table of the HiRID benchmark (Yèche et al., 2021): https://github.com/ratschlab/HIRID-ICU-Benchmark/blob/master/preprocessing/resources/varref.tsv.

## Fairness Analysis Report

### 1. Information about test dataset

#### Grouping by sex

**Table 1.a.**
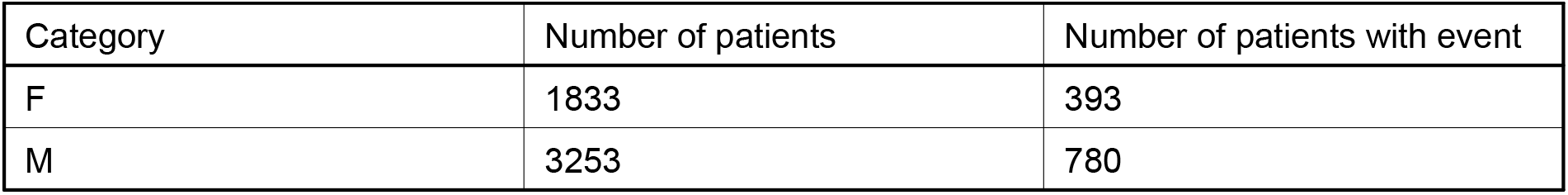

| Category | Number of patients | Number of patients with event |
| --- | --- | --- |
| F | 1833 | 393 |
| M | 3253 | 780 |

#### Grouping by age_group

**Table 1.b.**
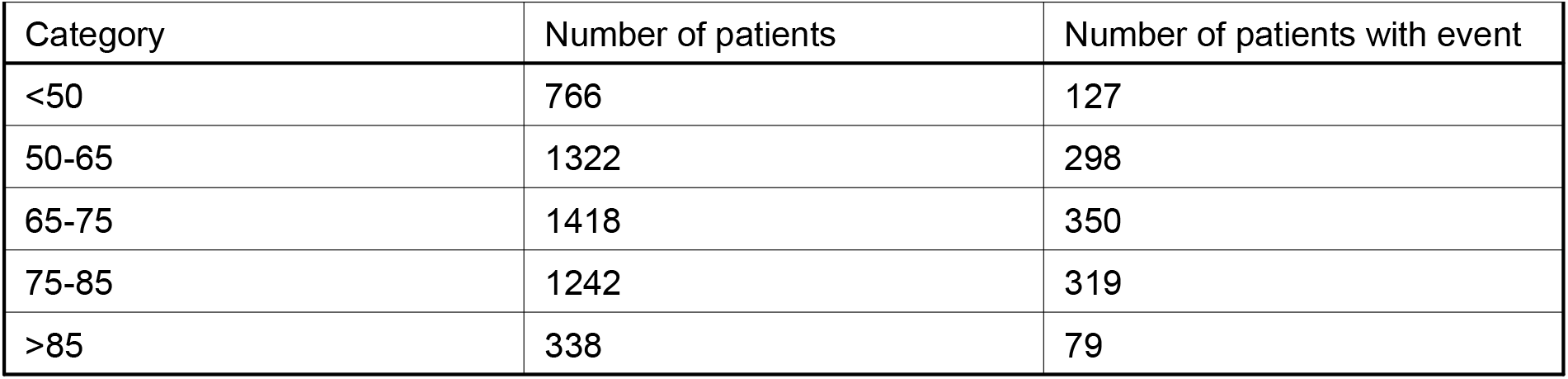

| Category | Number of patients | Number of patients with event |
| --- | --- | --- |
| <50 | 766 | 127 |
| 50-65 | 1322 | 298 |
| 65-75 | 1418 | 350 |
| 75-85 | 1242 | 319 |
| >85 | 338 | 79 |

#### Grouping by APACHE_group

**Table 1.c.**

| Category | Number of patients | Number of patients with event |
| --- | --- | --- |
| Cardiovascular | 1891 | 666 |
| Neurological | 1468 | 92 |
| Gastrointestinal | 522 | 151 |
| Respiratory | 471 | 102 |
| Other | 325 | 76 |
| Trauma | 279 | 61 |
| Metabolic | 98 | 19 |

#### Grouping by surgical_status

**Table 1.d.**
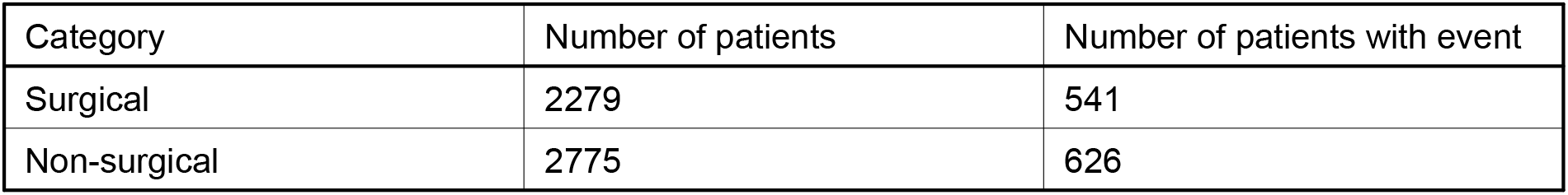

| Category | Number of patients | Number of patients with event |
| --- | --- | --- |
| Surgical | 2279 | 541 |
| Non-surgical | 2775 | 626 |

### 2. Model Performance Analysis

***Goal: Comparing the model performance across cohorts of patients***

Binary metrics computed with a threshold on score of 0.445.

#### 2.1. Aggregated views

##### 2.1.1. Summarized performance metrics per grouping

###### Grouping by sex

The minority category is F.

**Table 2.1.1.a.**

| Metric | Macro-average | Worst value (category) | For minority category |
| --- | --- | --- | --- |
| Recall ↑ | 0.201 | 0.198 (M) | 0.204 |
| Precision ↑ | 0.557 | 0.557 (M) | 0.558 |
| NPV ↑ | 0.965 | 0.962 (M) | 0.967 |
| FPR ↓ | 0.007 | 0.008 (M) | 0.007 |
| Corrected precision ↑ | 0.576 | 0.557 (M) | 0.596 |
| Corrected NPV ↑ | 0.967 | 0.967 (M) | 0.967 |
| Event-based recall ↑ | 0.805 | 0.793 (M) | 0.816 |
| Calibration error ↓ | 0.032 | 0.037 (F) | 0.037 |
| Avg. score on positive class | 0.255 | 0.252 (M) | 0.257 |
| Avg. score on negative class | 0.033 | 0.035 (M) | 0.031 |
| AUROC ↑ | 0.914 | 0.908 (M) | 0.921 |
| AUPRC ↑ | 0.39 | 0.385 (M) | 0.396 |
| Corrected AUPRC ↑ | 0.406 | 0.385 (M) | 0.427 |
| Event-based AUPRC ↑ | 0.694 | 0.674 (M) | 0.715 |
| Corrected event-based AUPRC ↑ | 0.707 | 0.674 (M) | 0.741 |

###### Grouping by age_group

The minority category is >85.

**Table 2.1.1.b.**

| Metric | Macro-average | Worst value (category) | For minority category |
| --- | --- | --- | --- |
| Recall ↑ | 0.199 | 0.184 (<50) | 0.204 |
| Precision ↑ | 0.598 | 0.522 (50-65) | 0.708 |
| NPV ↑ | 0.963 | 0.953 (>85) | 0.953 |
| FPR ↓ | 0.007 | 0.01 (75-85) | 0.006 |
| Corrected precision ↑ | 0.67 | 0.564 (75-85) | 0.719 |
| Corrected NPV ↑ | 0.98 | 0.98 (50-65) | 0.98 |
| Event-based recall ↑ | 0.793 | 0.751 (<50) | 0.795 |
| Calibration error ↓ | 0.054 | 0.082 (>85) | 0.082 |
| Avg. score on positive class | 0.257 | 0.253 (75-85) | 0.266 |
| Avg. score on negative class | 0.034 | 0.041 (75-85) | 0.038 |
| AUROC ↑ | 0.915 | 0.885 (75-85) | 0.916 |
| AUPRC ↑ | 0.408 | 0.372 (75-85) | 0.479 |
| Corrected AUPRC ↑ | 0.475 | 0.393 (75-85) | 0.489 |
| Event-based AUPRC ↑ | 0.717 | 0.654 (75-85) | 0.82 |
| Corrected event-based AUPRC ↑ | 0.768 | 0.676 (75-85) | 0.828 |

###### Grouping by APACHE_group

The minority category is Metabolic.

**Table 2.1.1.c.**

| Metric | Macro-average | Worst value (category) | For minority category |
| --- | --- | --- | --- |
| Recall ↑ | 0.18 | 0.058 (Neurological) | 0.164 |
| Precision ↑ | 0.552 | 0.386 (Neurological) | 0.709 |
| NPV ↑ | 0.959 | 0.944 (Cardiovascular) | 0.952 |
| FPR ↓ | 0.008 | 0.015 (Gastrointestinal) | 0.004 |
| Corrected precision ↑ | 0.679 | 0.58 (Gastrointestinal) | 0.766 |
| Corrected NPV ↑ | 0.99 | 0.989 (Neurological) | 0.991 |
| Event-based recall ↑ | 0.755 | 0.48 (Neurological) | 0.716 |
| Calibration error ↓ | 0.075 | 0.119 (Neurological) | 0.113 |
| Avg. score on positive class | 0.24 | 0.132 (Neurological) | 0.244 |
| Avg. score on negative class | 0.035 | 0.056 (Cardiovascular) | 0.032 |
| AUROC ↑ | 0.908 | 0.878 (Cardiovascular) | 0.93 |
| AUPRC ↑ | 0.383 | 0.172 (Neurological) | 0.466 |
| Corrected AUPRC ↑ | 0.491 | 0.415 (Cardiovascular) | 0.524 |
| Event-based AUPRC ↑ | 0.676 | 0.418 (Neurological) | 0.808 |
| Corrected event-based AUPRC ↑ | 0.77 | 0.707 (Cardiovascular) | 0.843 |

###### Grouping by surgical_status

The minority category is Surgical.

**Table 2.1.1.d.**

| Metric | Macro-average | Worst value (category) | For minority category |
| --- | --- | --- | --- |
| Recall ↑ | 0.205 | 0.194 (Non-surgical) | 0.217 |
| Precision ↑ | 0.554 | 0.553 (Non-surgical) | 0.555 |
| NPV ↑ | 0.963 | 0.961 (Surgical) | 0.961 |
| FPR ↓ | 0.008 | 0.009 (Surgical) | 0.009 |
| Corrected precision ↑ | 0.571 | 0.557 (Surgical) | 0.557 |
| Corrected NPV ↑ | 0.966 | 0.965 (Non-surgical) | 0.966 |
| Event-based recall ↑ | 0.802 | 0.799 (Non-surgical) | 0.804 |
| Calibration error ↓ | 0.033 | 0.033 (Surgical) | 0.033 |
| Avg. score on positive class | 0.258 | 0.248 (Non-surgical) | 0.268 |
| Avg. score on negative class | 0.035 | 0.04 (Surgical) | 0.04 |
| AUROC ↑ | 0.912 | 0.911 (Surgical) | 0.911 |
| AUPRC ↑ | 0.388 | 0.385 (Non-surgical) | 0.39 |
| Corrected AUPRC ↑ | 0.4 | 0.391 (Surgical) | 0.391 |
| Event-based AUPRC ↑ | 0.687 | 0.682 (Surgical) | 0.682 |
| Corrected event-based AUPRC ↑ | 0.698 | 0.682 (Surgical) | 0.682 |

##### 2.1.2. Summary view based on the ratio of significantly worse metrics

###### We first show an overview of this analysis over all groupings

<u>Worst ratio</u>: 73.3% for category 75-85 (age_group) with the biggest delta 0.053 on Corrected event-based AUPRC.

<u>Worst delta</u>: 0.346 on Event-based recall for category Neurological (APACHE_group).

In the following tables, we display the ratio of significantly worse metrics (over the total number of analysed performance metrics) for each category of patients.

###### Grouping by sex

<u>Worst ratio</u>: 60.0% for category M with the biggest delta 0.068 on Corrected event-based AUPRC.

<u>Worst delta</u> is the same as above.

**Table 2.1.2.a.**
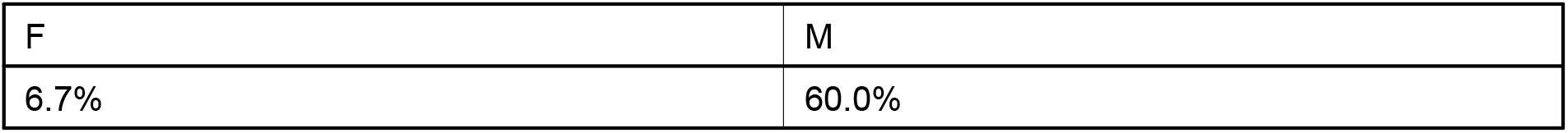

###### Grouping by age_group

<u>Worst ratio</u>: 73.3% for category 75-85 with the biggest delta 0.053 on Corrected event-based AUPRC.

<u>Worst delta</u>: 0.056 on Event-based recall for category <50.

**Table 2.1.2.b.**

| <50 | 50-65 | 65-75 | 75-85 | >85 |
| --- | --- | --- | --- | --- |
| 20.0% | 33.3% | 33.3% | 73.3% | 20.0% |

###### Grouping by APACHE_group

<u>Worst ratio</u>: 46.7% for category Cardiovascular with the biggest delta 0.104 on Corrected precision.

<u>Worst delta</u>: 0.346 on Event-based recall for category Neurological.

**Table 2.1.2.c.**

| Cardiovascular | Neurological | Gastrointestinal | Respiratory | Other | Trauma | Metabolic |
| --- | --- | --- | --- | --- | --- | --- |
| 46.7% | 46.7% | 40.0% | 40.0% | 33.3% | 6.7% | 26.7% |

###### Grouping by surgical_status

<u>Worst ratio</u>: 40.0% for category Surgical with the biggest delta 0.031 on Corrected event-based AUPRC.

<u>Worst delta</u> is the same as above.

**Table 2.1.2.d.**

| Surgical | Non-surgical |
| --- | --- |
| 40.0% | 13.3% |

##### 2.1.3. Top 3 cohorts with the biggest performance metric discrepancies

In the following table, we show for each performance metric the 3 cohorts with the biggest delta that are significantly worse off than the rest of the patients. If some cells are empty, this means that there are less than 3 cohorts, possibly none, that are significantly worse than the rest of the patients for this particular metric.

**Table 2.1.3.a.**

| Metric | Cohort 1 ($\Delta$) | Cohort 2 ($\Delta$) | Cohort 3 ($\Delta$) |
| --- | --- | --- | --- |
| Recall ↑ | Neurological (0.156) | Respiratory (0.047) | Metabolic (0.038) |
| Precision ↑ | Neurological (0.176) | 50-65 (0.048) | 75-85 (0.03) |
| NPV ↑ | Cardiovascular (0.028) | Gastrointestinal (0.016) | Other (0.014) |
| FPR ↓ | Gastrointestinal (0.008) | Cardiovascular (0.007) | 75-85 (0.004) |
| Corrected precision ↑ | Cardiovascular (0.104) | Gastrointestinal (0.067) | 75-85 (0.04) |
| Corrected NPV ↑ | - | - | - |
| Event-based recall ↑ | Neurological (0.346) | Metabolic (0.088) | <50 (0.056) |
| Calibration error ↓ | Neurological (0.096) | Metabolic (0.09) | >85 (0.056) |
| Avg. score on positive class | Neurological (0.133) | Respiratory (0.026) | Non-surgical (0.02) |
| Avg. score on negative class | Cardiovascular (0.031) | Gastrointestinal (0.013) | Surgical (0.01) |
| AUROC ↑ | Cardiovascular (0.045) | 75-85 (0.034) | Respiratory (0.027) |
| AUPRC ↑ | Neurological (0.233) | 75-85 (0.023) | 50-65 (0.016) |
| Corrected AUPRC ↑ | Cardiovascular (0.084) | M (0.042) | 75-85 (0.035) |
| Event-based AUPRC ↑ | Neurological (0.281) | 75-85 (0.046) | M (0.041) |
| Corrected event-based AUPRC ↑ | Cardiovascular (0.079) | M (0.068) | 75-85 (0.053) |

#### 2.2. Grouping by

For each grouping, we display box plots that show the performance metrics’ distributions for the different categories of patients. For each metric, we emphasize with a black star the cohorts that are significantly worse off compared to the rest of the patients and with a red star the cohorts that appear in the table **Top 3 cohorts with the biggest performance metric discrepancies**.

For each grouping, we propose a table that presents the results of the statistical analysis: comparing the different performance metrics for a cohort against the rest of the patients. P-values are obtained by running the Mann-Whitney U test with Bonferroni correction. We display only metrics and cohorts with a significant p-value (smaller than 0.001/number of comparisons) and whose delta is bigger than 0. For binary grouping, we display the category with the worst distribution for each metric. While for multicategorical grouping, we display whether the distribution for the category is better or worse than for the rest of patients

We also display the calibration curve for each grouping’s categories as well as the curves corresponding to each score-based metrics.

##### 2.2.1. … sex

**Figure 2.2.1.a.**
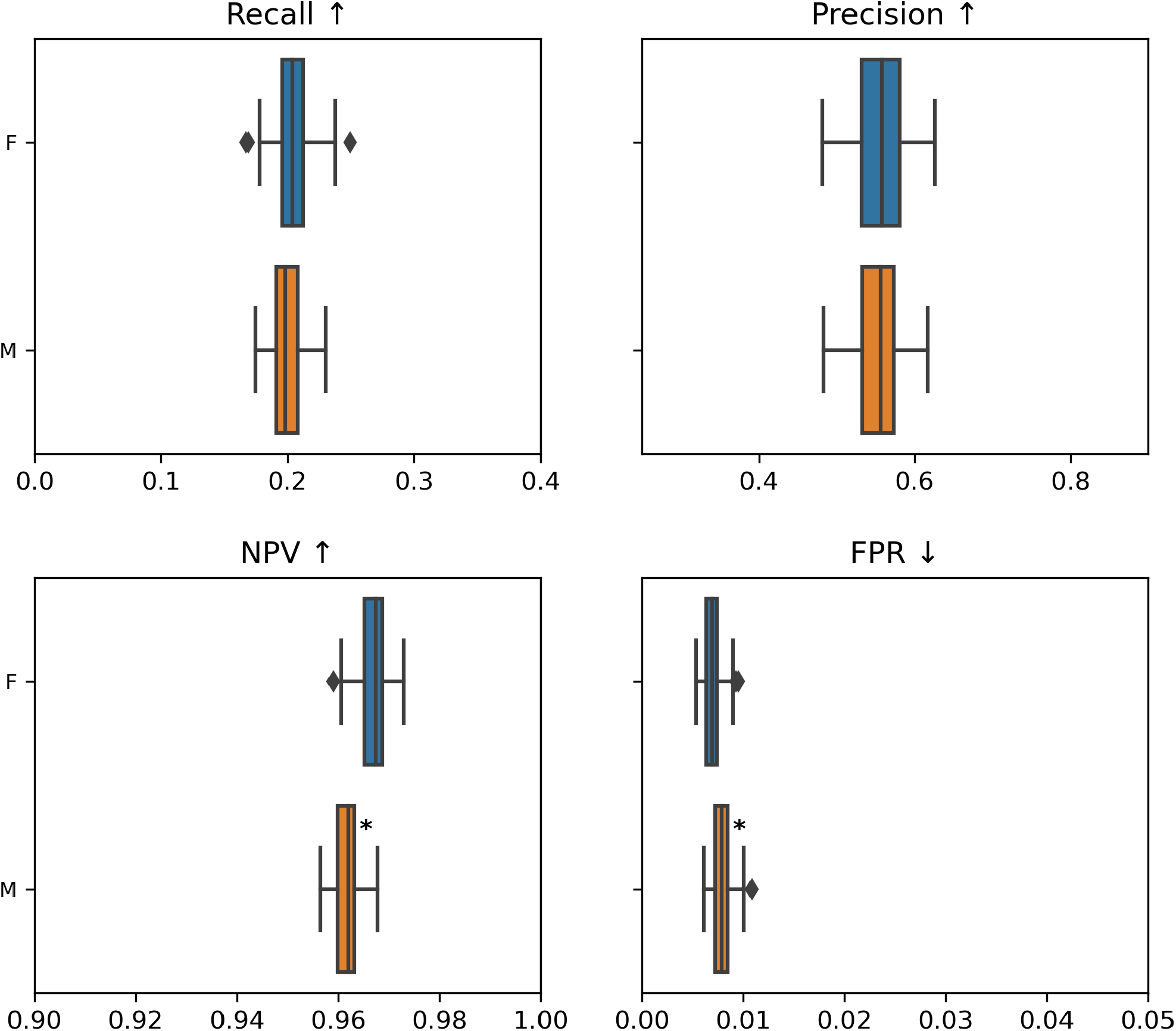
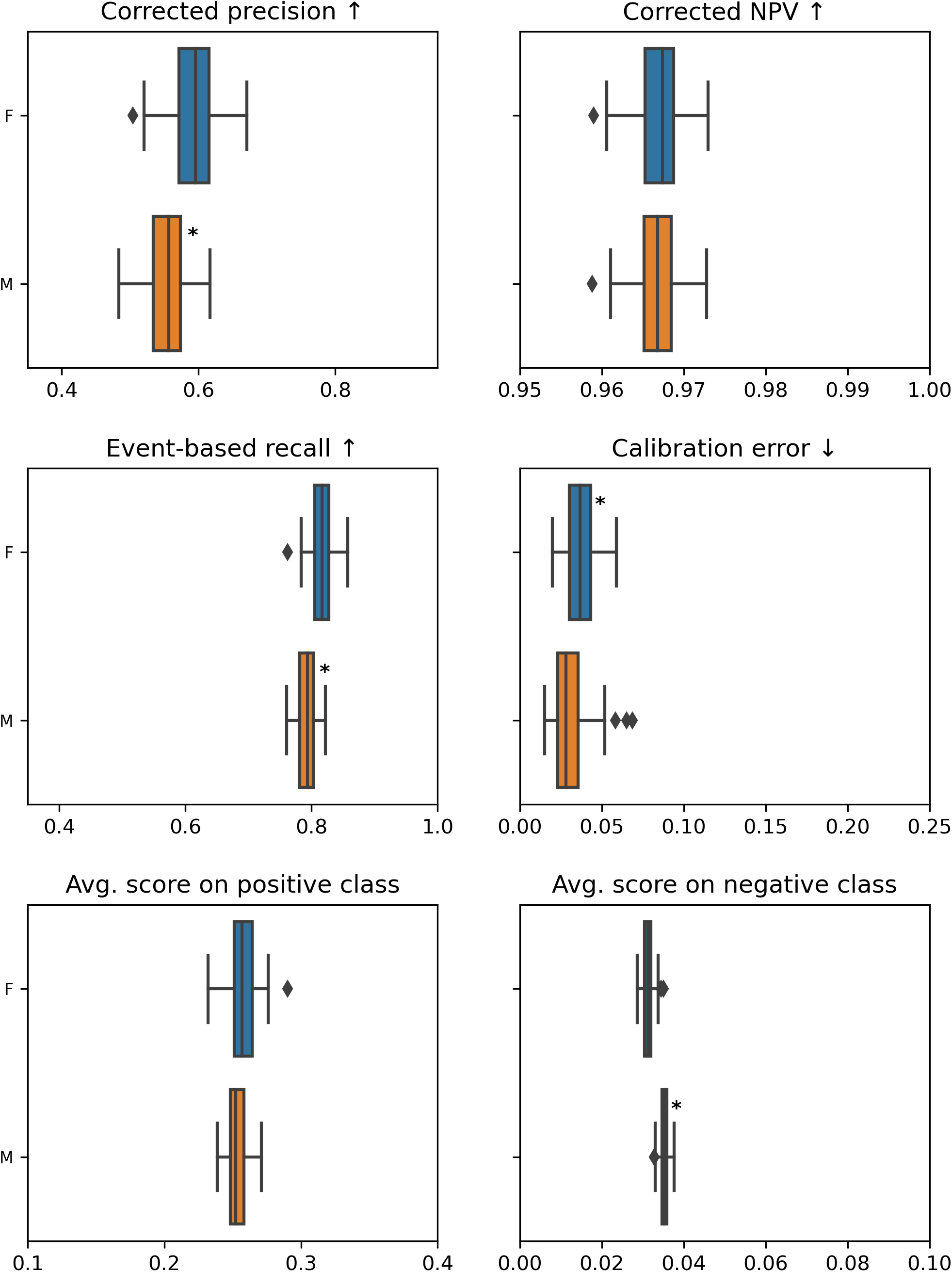
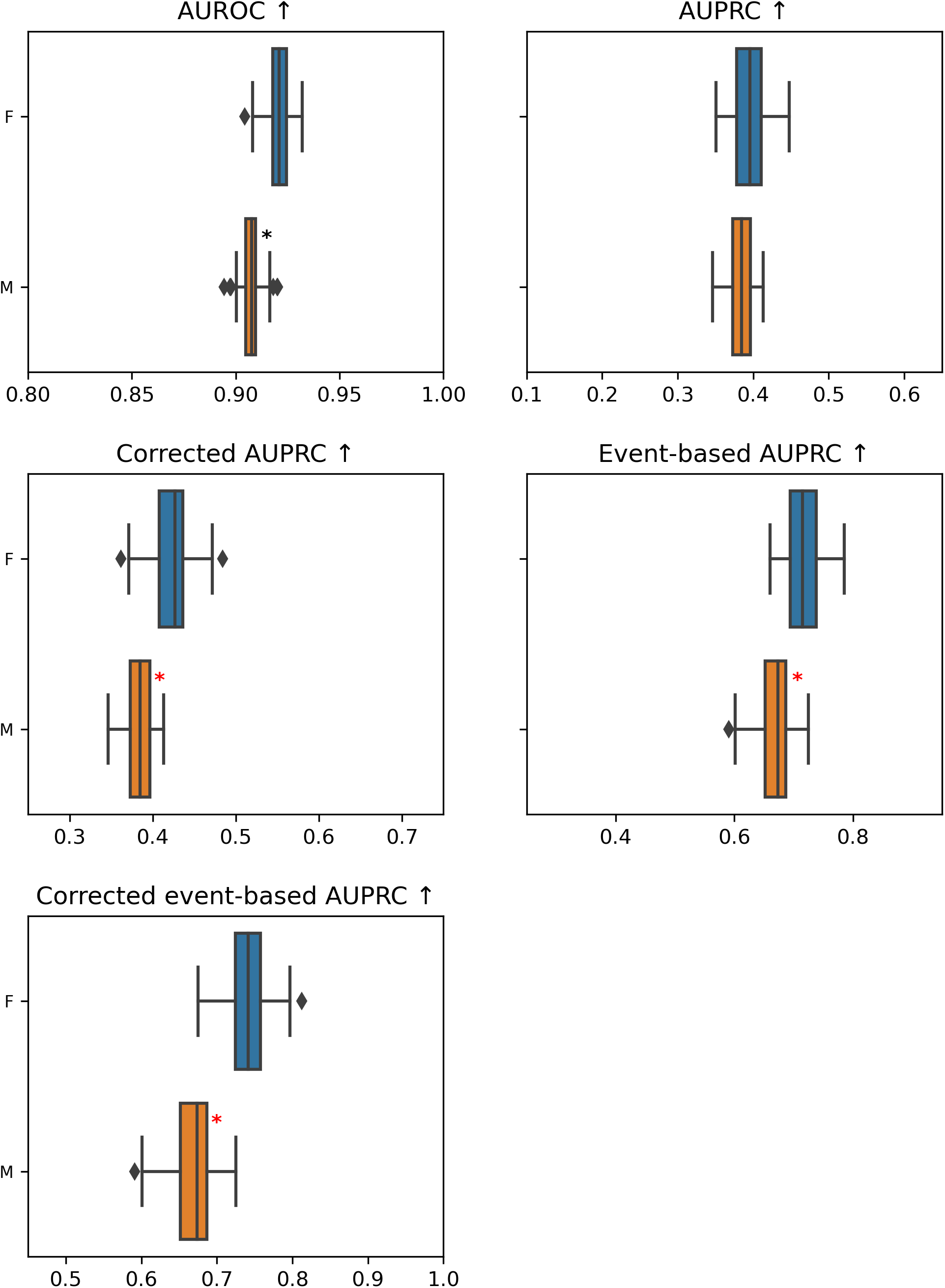

**Table 2.2.1.a.**

| Metric | Cohort with the worst metric | P-value | Delta |
| --- | --- | --- | --- |
| NPV ↑ | M | 1.30e-25 | 0.005 |
| FPR ↓ | M | 1.89e-12 | 0.001 |
| Corrected precision ↑ | M | 1.96e-14 | 0.039 |
| Event-based recall ↑ | M | 6.82e-20 | 0.023 |
| Calibration error ↓ | F | 5.68e-09 | 0.008 |
| Avg. score on negative class | M | 1.92e-33 | 0.004 |
| AUROC ↑ | M | 3.57e-30 | 0.013 |
| Corrected AUPRC ↑ | M | 9.56e-26 | 0.042 |
| Event-based AUPRC ↑ | M | 1.13e-18 | 0.041 |
| Corrected event-based AUPRC ↑ | M | 7.40e-31 | 0.068 |

**Figure 2.2.1.b.**
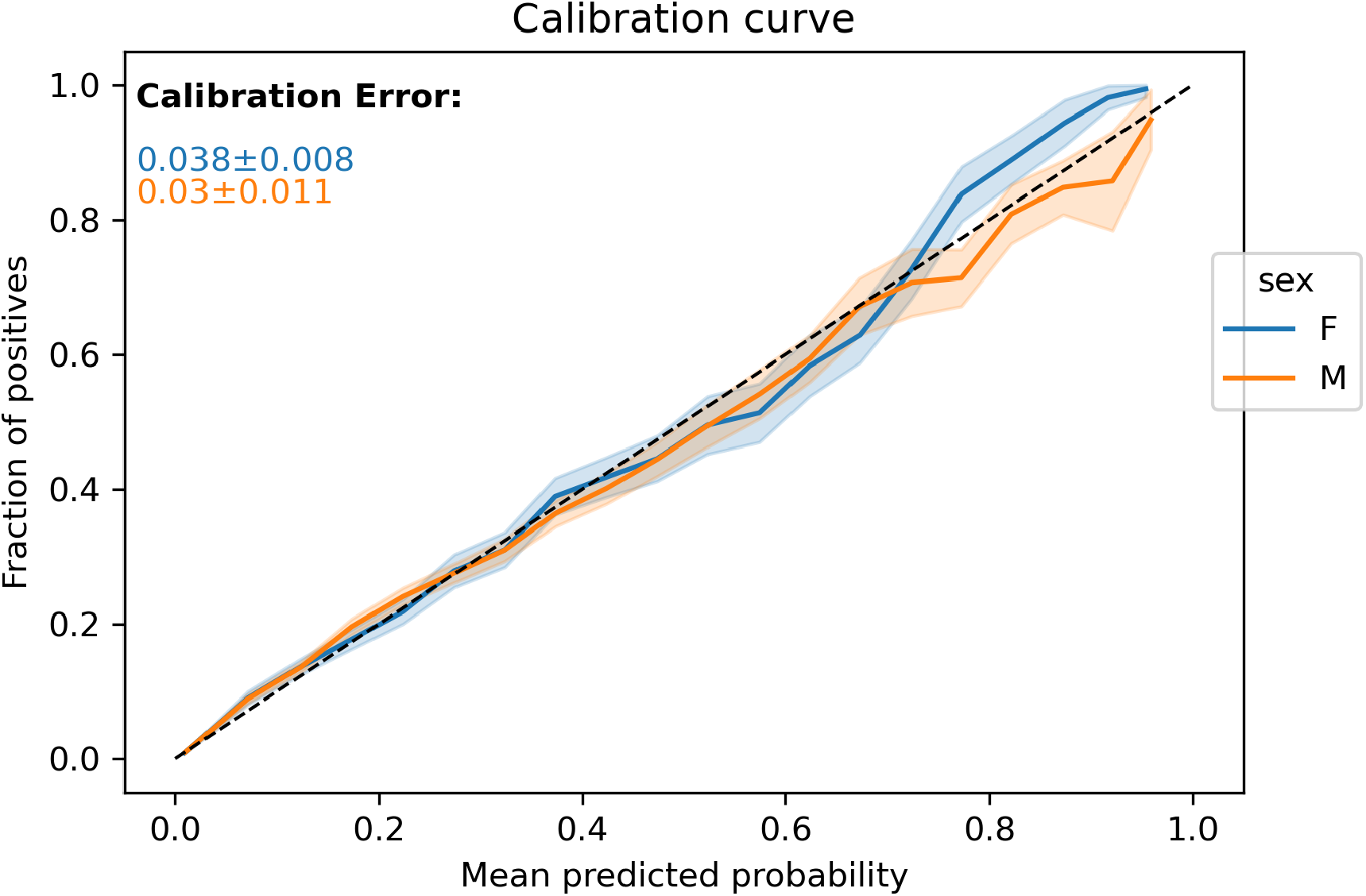

**Figure 2.2.1.c.**
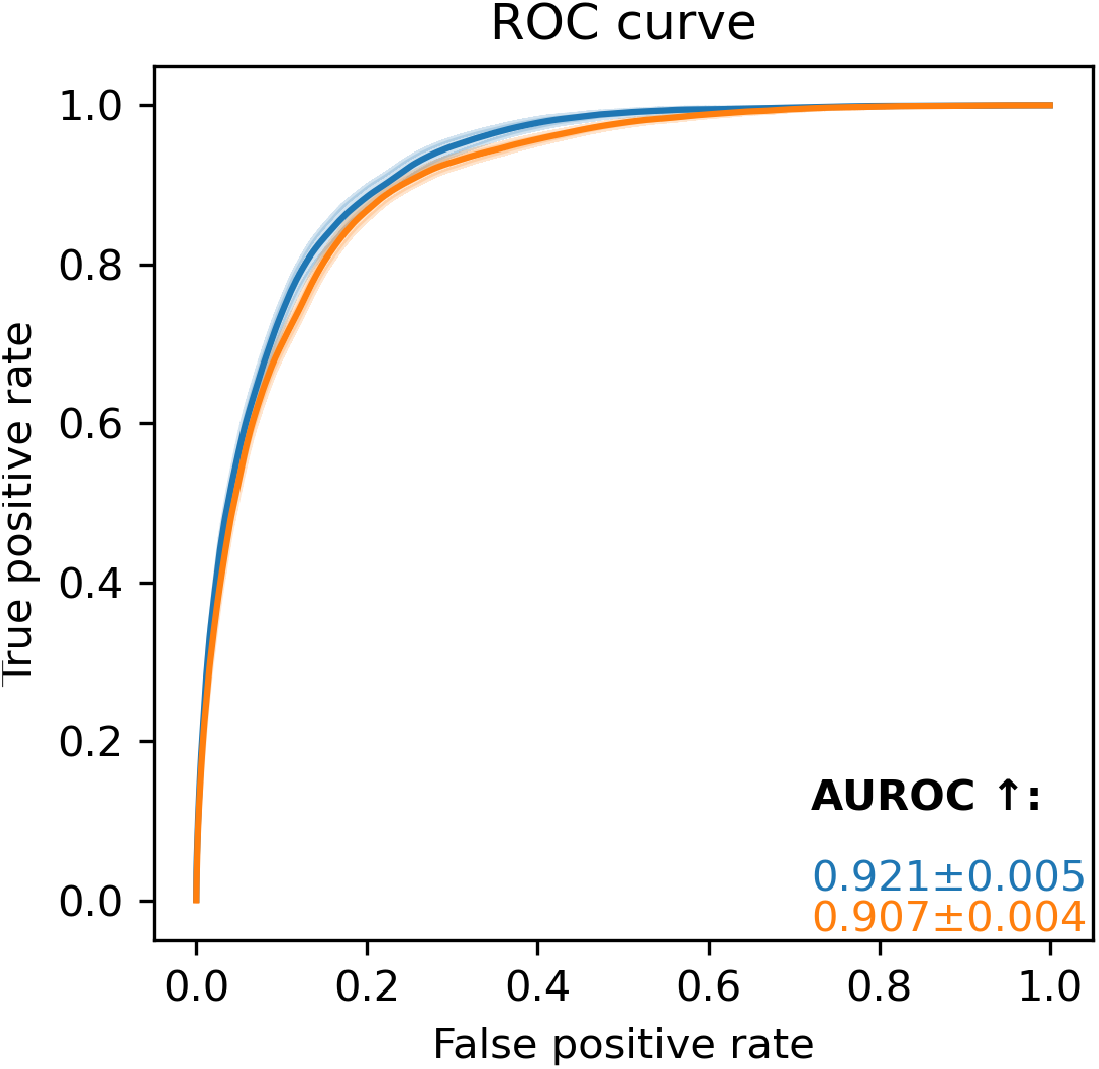

**Figure 2.2.1.d.**
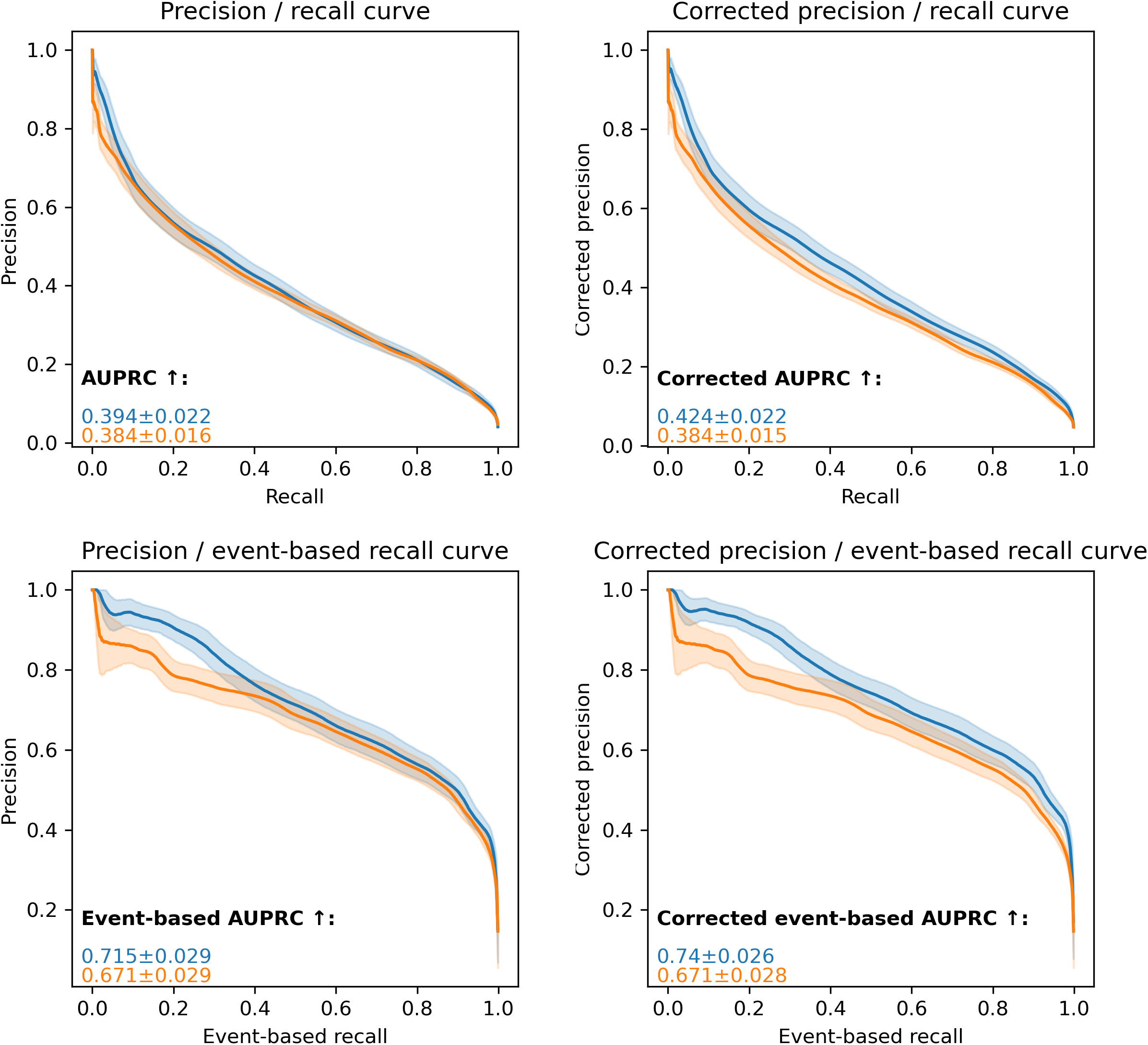

##### 2.2.2. … age_group

**Figure 2.2.2.a.**
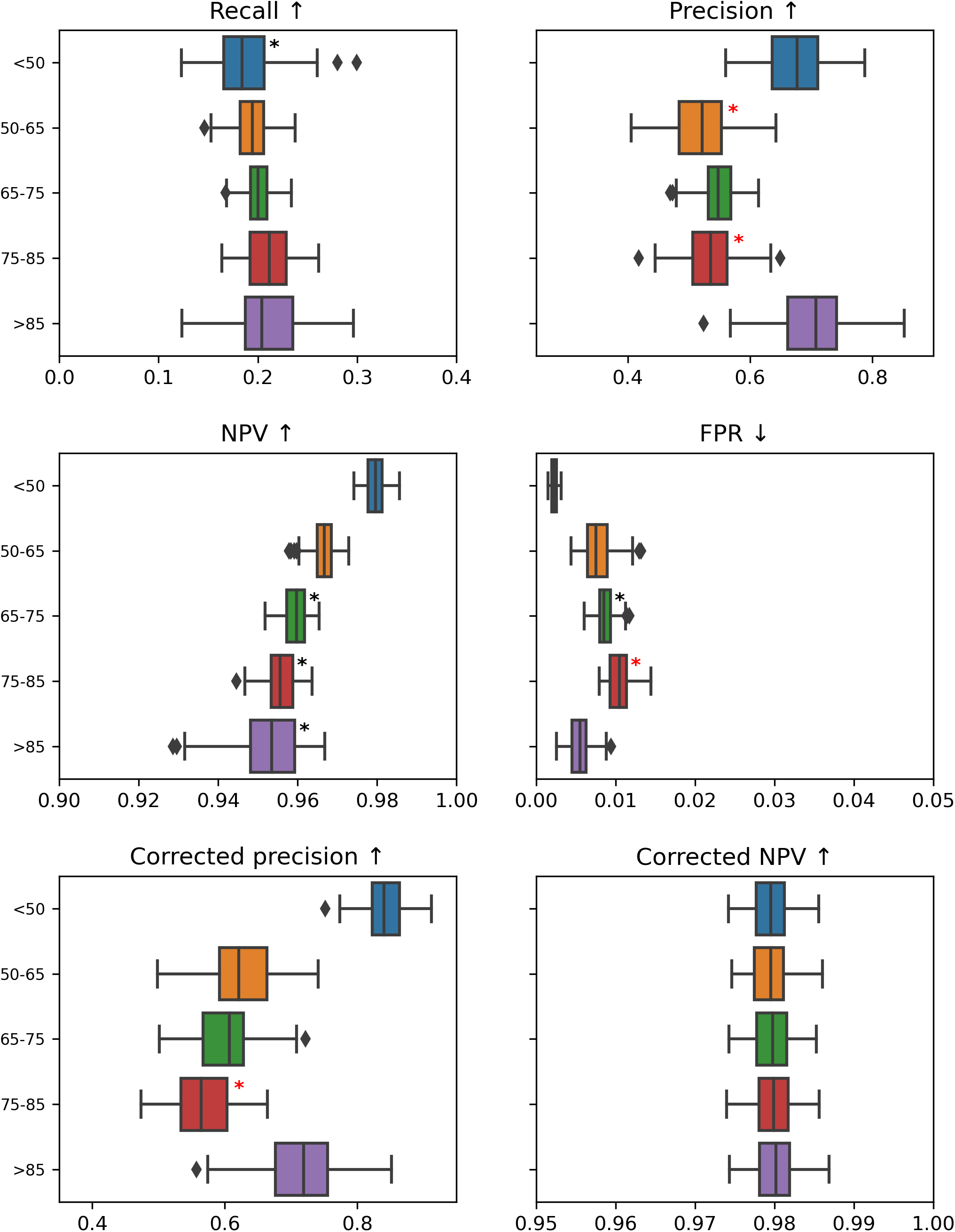
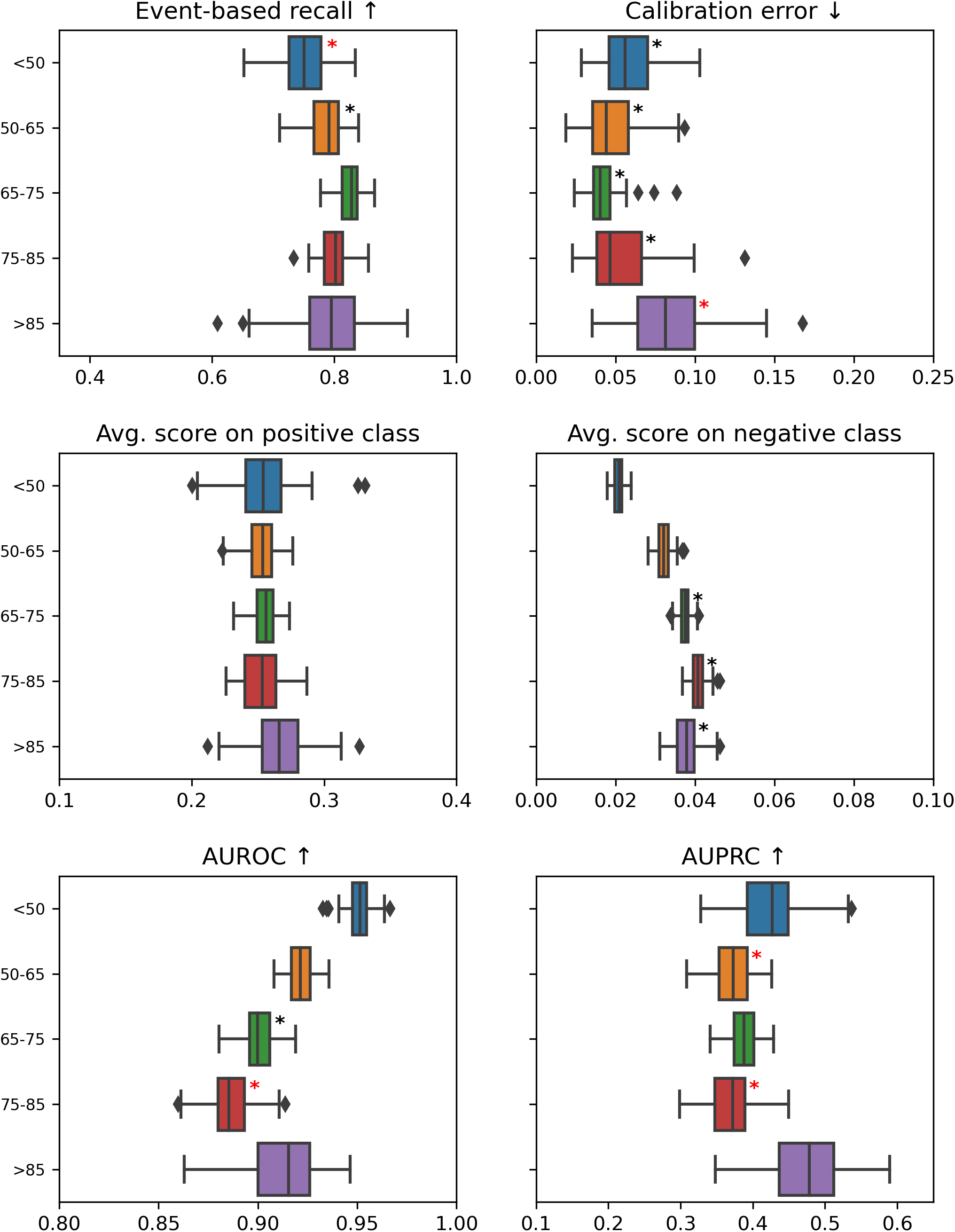
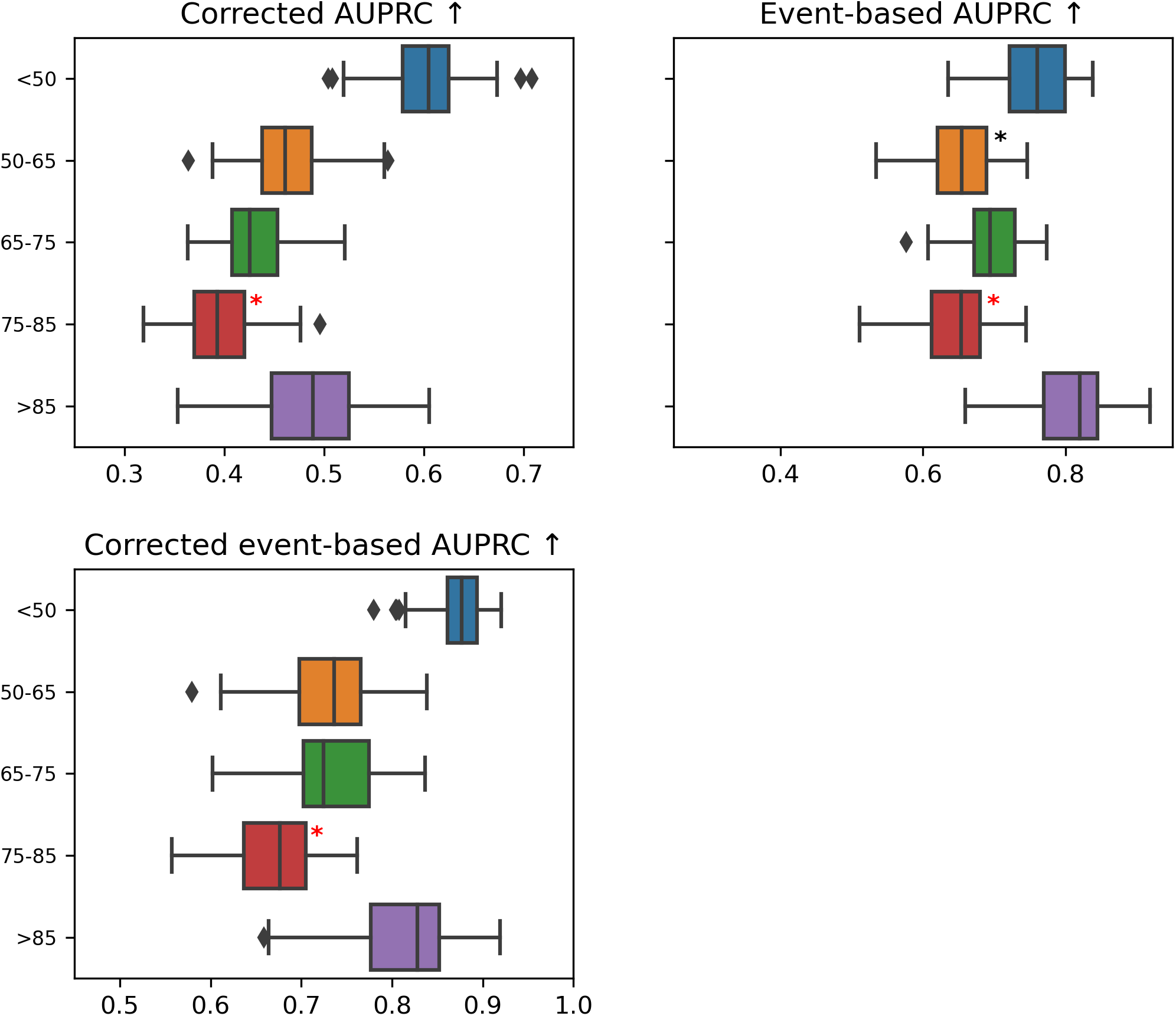

**Table 2.2.2.a.**

| Metric | Category | Cohort vs. rest | P-value | Delta |
| --- | --- | --- | --- | --- |
| Recall ↑ | <50 | worse | 9.18e-07 | 0.018 |
| Recall ↑ | 75-85 | better | 8.03e-07 | 0.013 |
| Precision ↑ | <50 | better | 4.50e-34 | 0.13 |
| Precision ↑ | 50-65 | worse | 7.48e-14 | 0.048 |
| Precision ↑ | 75-85 | worse | 6.77e-07 | 0.03 |
| Precision ↑ | >85 | better | 6.61e-33 | 0.162 |
| NPV ↑ | <50 | better | 1.28e-34 | 0.019 |
| NPV ↑ | 50-65 | better | 3.88e-19 | 0.004 |
| NPV ↑ | 65-75 | worse | 1.93e-28 | 0.006 |
| NPV ↑ | 75-85 | worse | 2.48e-34 | 0.011 |
| NPV ↑ | >85 | worse | 1.70e-27 | 0.011 |
| FPR ↓ | <50 | better | 1.28e-34 | 0.006 |
| FPR ↓ | 65-75 | worse | 2.98e-19 | 0.001 |
| FPR ↓ | 75-85 | worse | 4.50e-34 | 0.004 |
| FPR ↓ | >85 | better | 2.66e-25 | 0.002 |
| Corrected precision ↑ | <50 | better | 1.28e-34 | 0.294 |
| Corrected precision ↑ | 50-65 | better | 5.62e-11 | 0.04 |
| Corrected precision ↑ | 75-85 | worse | 2.27e-09 | 0.04 |
| Corrected precision ↑ | >85 | better | 5.35e-32 | 0.146 |
| Corrected NPV ↑ | <50 | better | 1.28e-34 | 0.013 |
| Corrected NPV ↑ | 50-65 | better | 1.28e-34 | 0.013 |
| Corrected NPV ↑ | 65-75 | better | 1.28e-34 | 0.013 |
| Corrected NPV ↑ | 75-85 | better | 1.28e-34 | 0.014 |
| Corrected NPV ↑ | >85 | better | 1.28e-34 | 0.014 |
| Event-based recall ↑ | <50 | worse | 6.44e-24 | 0.056 |
| Event-based recall ↑ | 50-65 | worse | 2.22e-06 | 0.013 |
| Event-based recall ↑ | 65-75 | better | 6.23e-27 | 0.041 |
| Calibration error ↓ | <50 | worse | 2.23e-29 | 0.029 |
| Calibration error ↓ | 50-65 | worse | 1.03e-25 | 0.021 |
| Calibration error ↓ | 65-75 | worse | 1.01e-20 | 0.014 |
| Calibration error ↓ | 75-85 | worse | 7.78e-26 | 0.019 |
| Calibration error ↓ | >85 | worse | 8.92e-34 | 0.056 |
| Avg. score on positive class | >85 | better | 3.18e-08 | 0.012 |
| Avg. score on negative class | <50 | better | 1.28e-34 | 0.016 |
| Avg. score on negative class | 50-65 | better | 1.82e-20 | 0.002 |
| Avg. score on negative class | 65-75 | worse | 2.13e-34 | 0.005 |
| Avg. score on negative class | 75-85 | worse | 1.28e-34 | 0.009 |
| Avg. score on negative class | >85 | worse | 1.51e-21 | 0.004 |
| AUROC ↑ | <50 | better | 1.28e-34 | 0.048 |
| AUROC ↑ | 50-65 | better | 5.09e-29 | 0.013 |
| AUROC ↑ | 65-75 | worse | 1.59e-30 | 0.017 |
| AUROC ↑ | 75-85 | worse | 1.49e-34 | 0.034 |
| AUPRC ↑ | <50 | better | 6.73e-13 | 0.044 |
| AUPRC ↑ | 50-65 | worse | 2.03e-07 | 0.016 |
| AUPRC ↑ | 75-85 | worse | 1.33e-09 | 0.023 |
| AUPRC ↑ | >85 | better | 1.44e-29 | 0.097 |
| Corrected AUPRC ↑ | <50 | better | 1.28e-34 | 0.222 |
| Corrected AUPRC ↑ | 50-65 | better | 4.89e-26 | 0.061 |
| Corrected AUPRC ↑ | 75-85 | worse | 7.34e-14 | 0.035 |
| Corrected AUPRC ↑ | >85 | better | 3.98e-27 | 0.086 |
| Event-based AUPRC ↑ | <50 | better | 7.33e-25 | 0.083 |
| Event-based AUPRC ↑ | 50-65 | worse | 8.e-11 | 0.041 |
| Event-based AUPRC ↑ | 75-85 | worse | 4.99e-15 | 0.046 |
| Event-based AUPRC ↑ | >85 | better | 1.13e-30 | 0.145 |
| Corrected event-based AUPRC ↑ | <50 | better | 1.28e-34 | 0.199 |
| Corrected event-based AUPRC ↑ | 50-65 | better | 2.24e-06 | 0.031 |
| Corrected event-based AUPRC ↑ | 75-85 | worse | 1.37e-18 | 0.053 |
| Corrected event-based AUPRC ↑ | >85 | better | 5.53e-29 | 0.132 |

**Figure 2.2.2.b.**
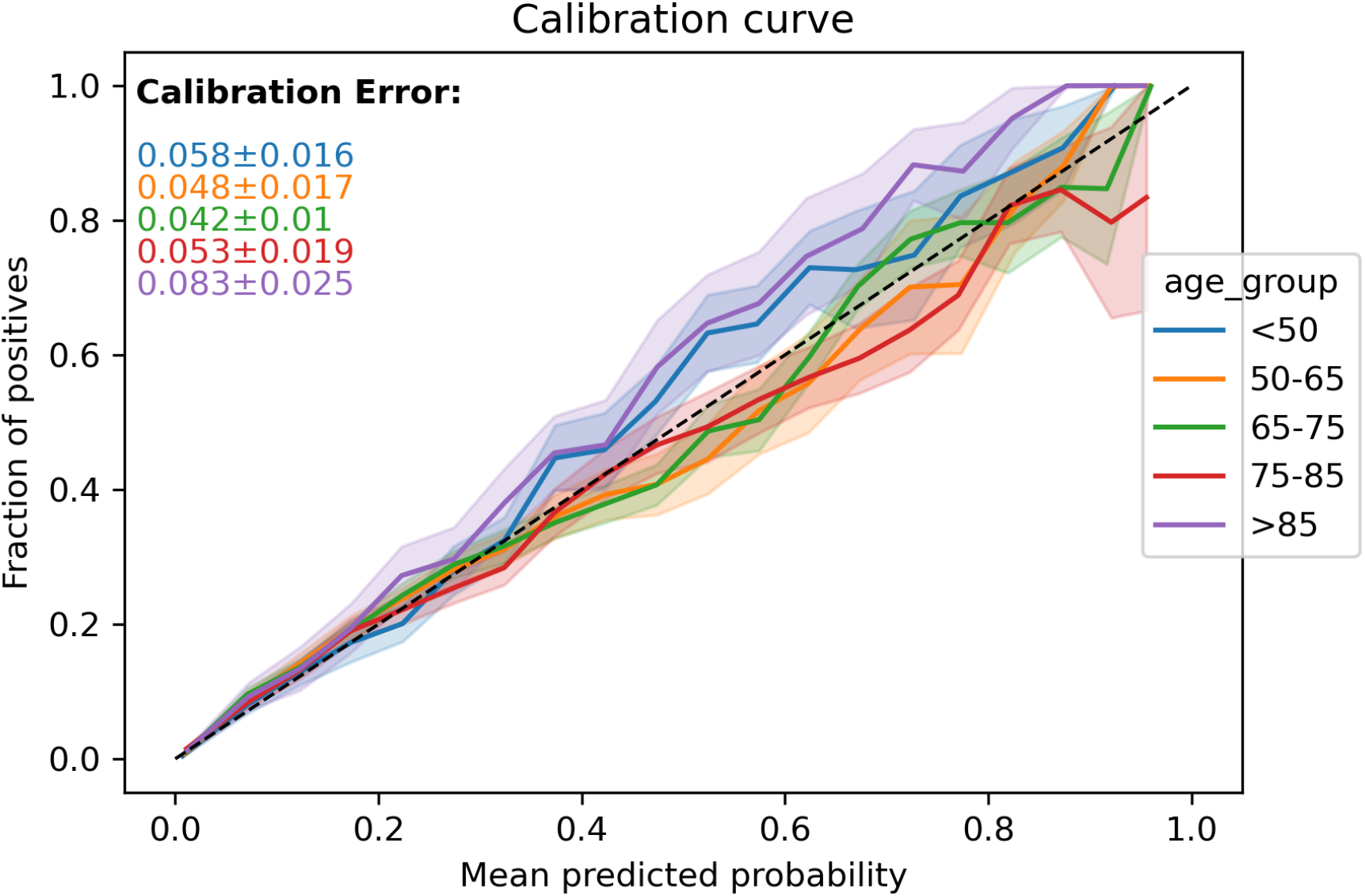

**Figure 2.2.2.c.**
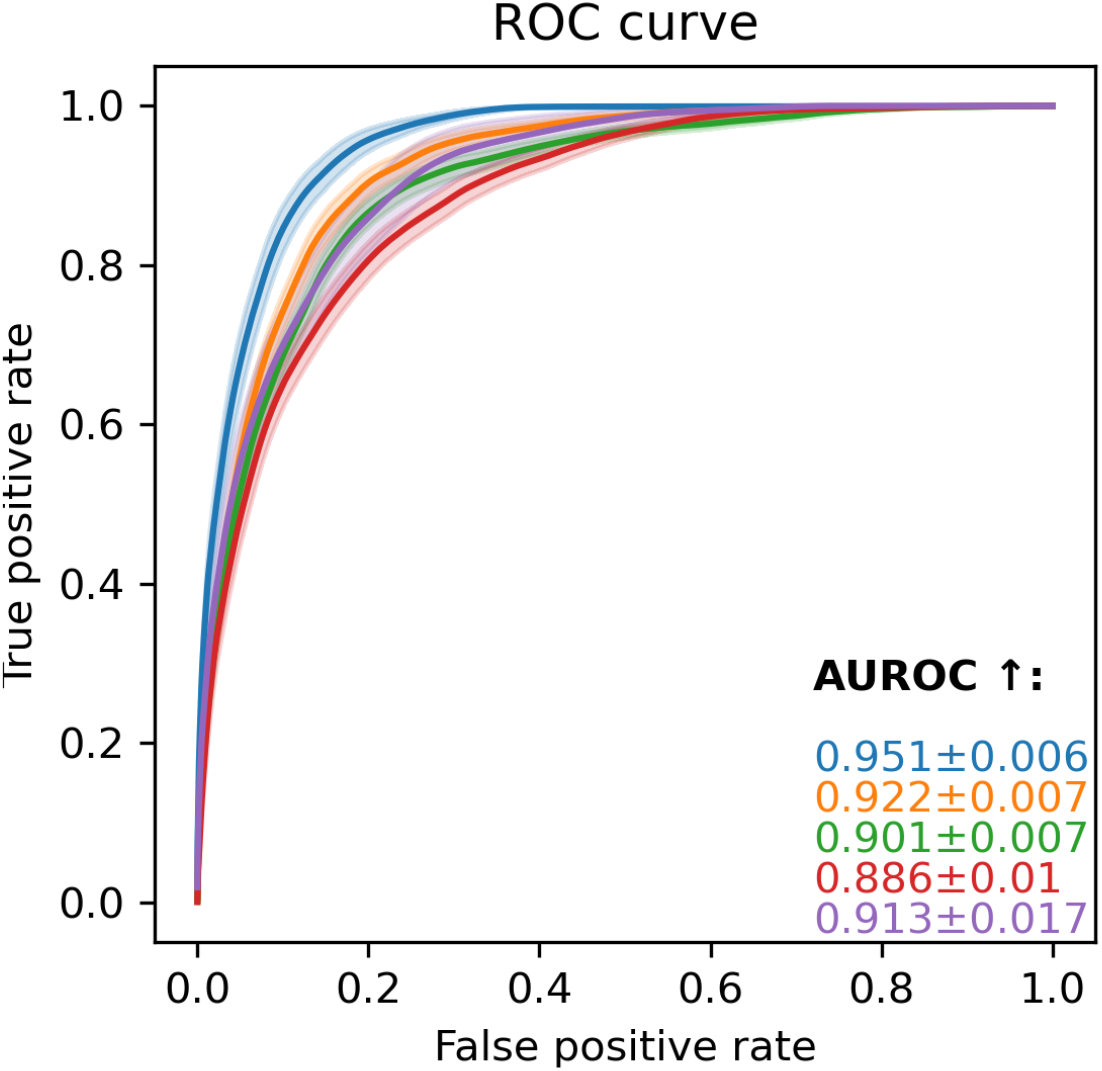

**Figure 2.2.2.d.**
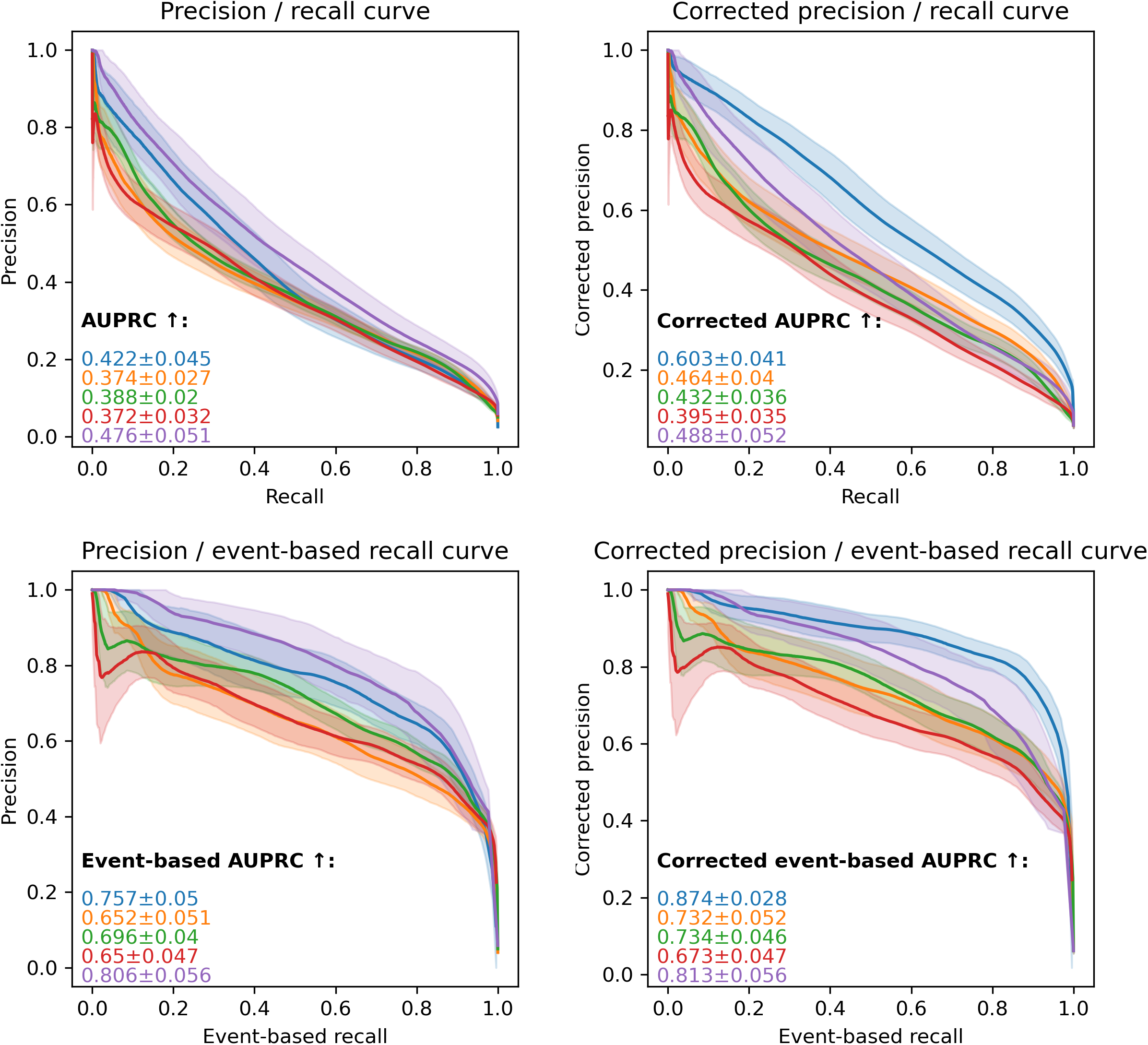

##### 2.2.3. … APACHE_group

**Figure 2.2.3.a.**
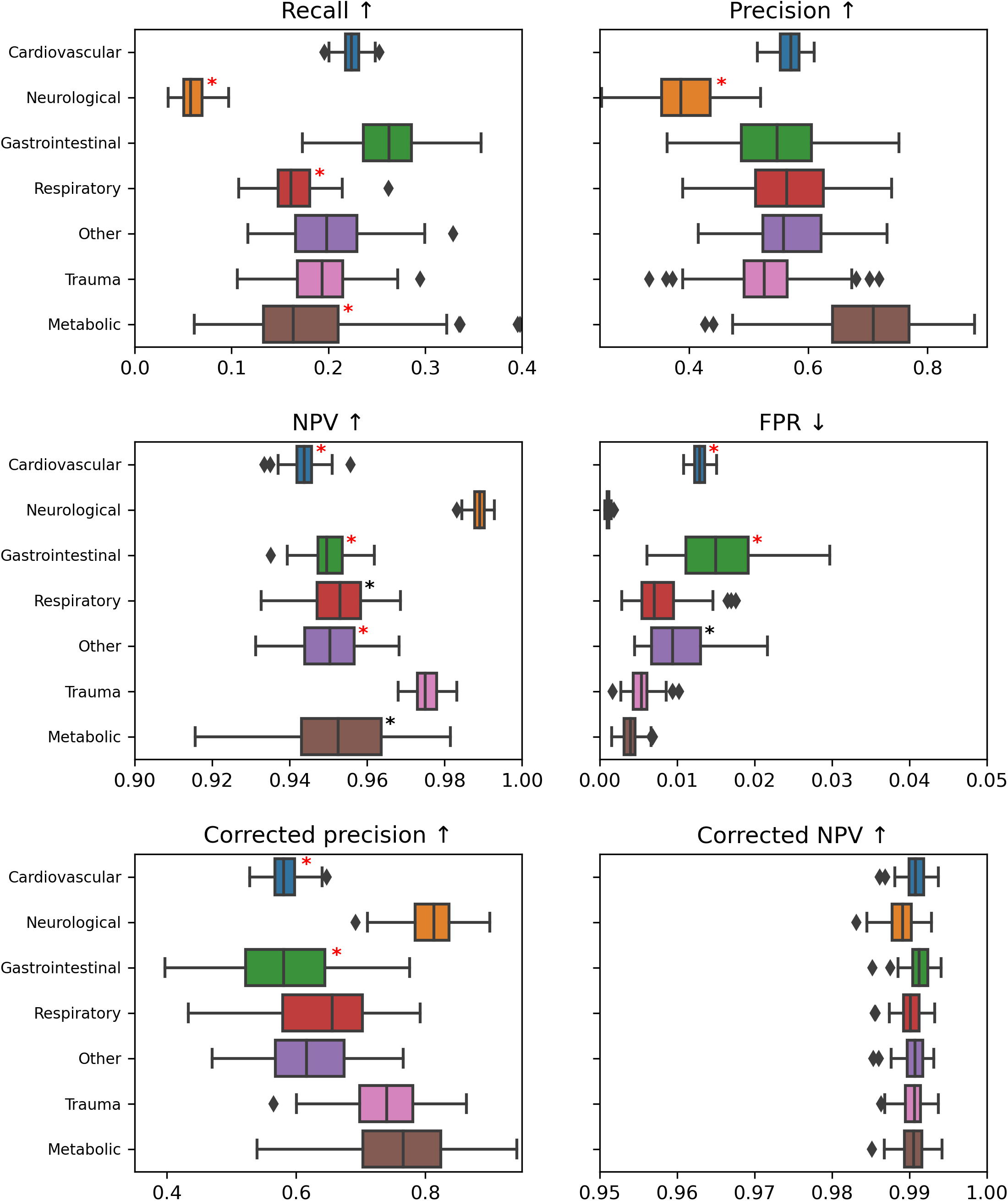
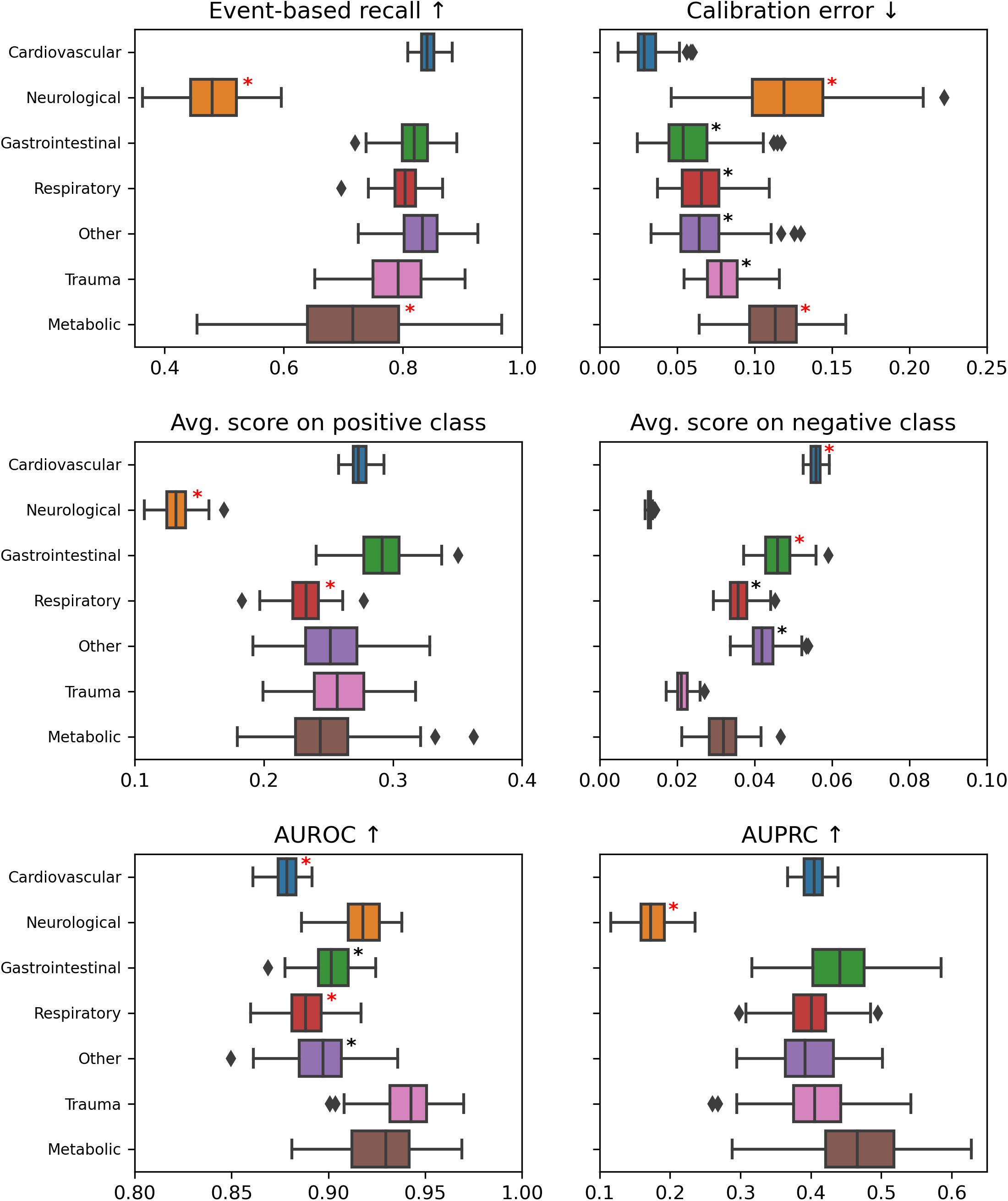
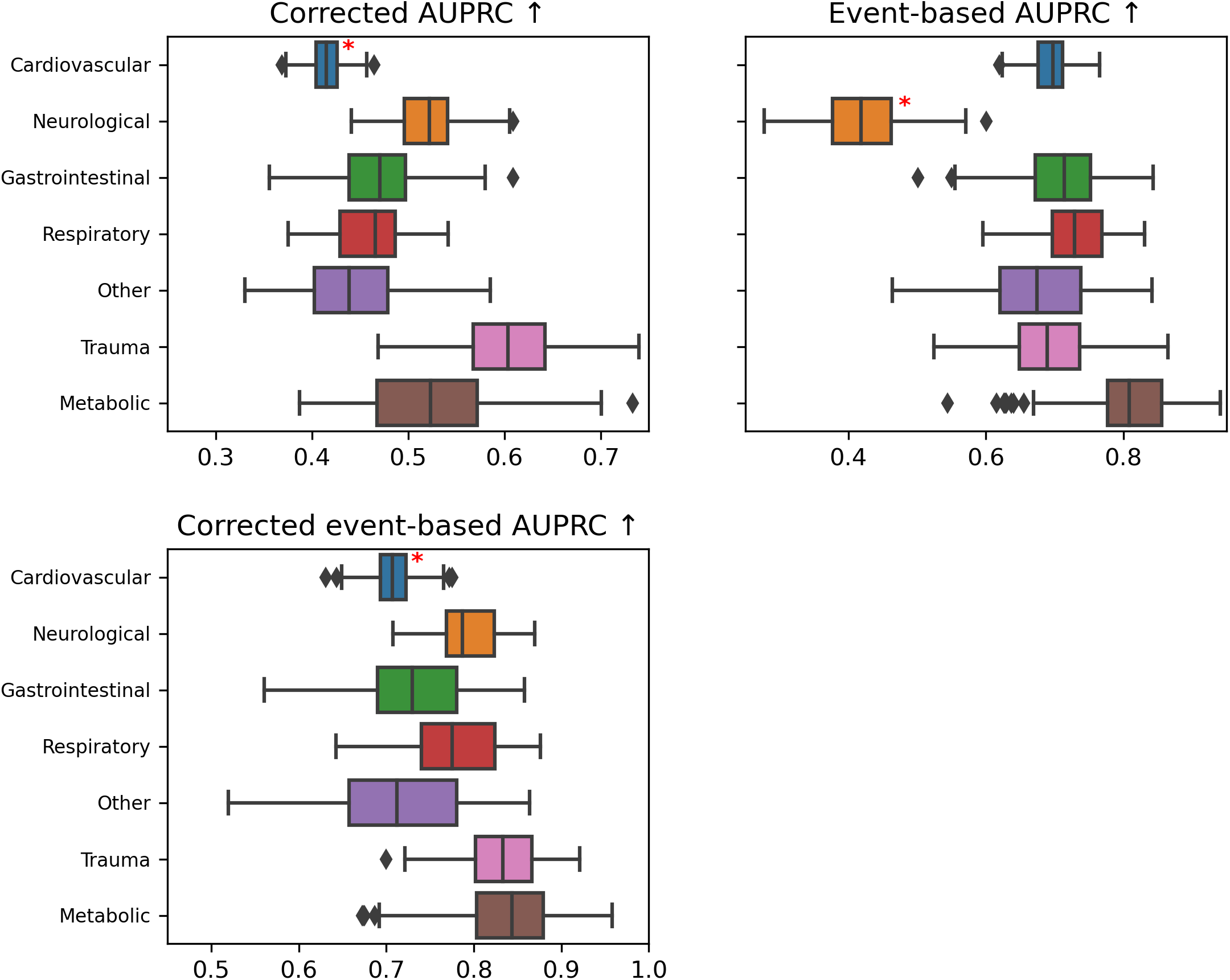

**Table 2.2.3.a.**

| Metric | Category | Cohort vs. rest | P-value | Delta |
| --- | --- | --- | --- | --- |
| Recall ↑ | Cardiovascular | better | 1.38e-31 | 0.044 |
| Recall ↑ | Neurological | worse | 1.28e-34 | 0.156 |
| Recall ↑ | Gastrointestinal | better | 7.78e-32 | 0.071 |
| Recall ↑ | Respiratory | worse | 7.05e-28 | 0.047 |
| Recall ↑ | Metabolic | worse | 1.05e-07 | 0.038 |
| Precision ↑ | Cardiovascular | better | 2.35e-08 | 0.027 |
| Precision ↑ | Neurological | worse | 1.44e-34 | 0.176 |
| Precision ↑ | Metabolic | better | 1.64e-22 | 0.155 |
| NPV ↑ | Cardiovascular | worse | 1.28e-34 | 0.028 |
| NPV ↑ | Neurological | better | 1.28e-34 | 0.038 |
| NPV ↑ | Gastrointestinal | worse | 1.63e-34 | 0.016 |
| NPV ↑ | Respiratory | worse | 2.16e-30 | 0.012 |
| NPV ↑ | Other | worse | 5.91e-30 | 0.014 |
| NPV ↑ | Trauma | better | 1.28e-34 | 0.012 |
| NPV ↑ | Metabolic | worse | 4.35e-10 | 0.011 |
| FPR ↓ | Cardiovascular | worse | 1.28e-34 | 0.007 |
| FPR ↓ | Neurological | better | 1.28e-34 | 0.01 |
| FPR ↓ | Gastrointestinal | worse | 4.52e-33 | 0.008 |
| FPR ↓ | Other | worse | 2.04e-06 | 0.002 |
| FPR ↓ | Trauma | better | 9.45e-22 | 0.002 |
| FPR ↓ | Metabolic | better | 1.13e-33 | 0.004 |
| Corrected precision ↑ | Cardiovascular | worse | 5.07e-34 | 0.104 |
| Corrected precision ↑ | Neurological | better | 1.28e-34 | 0.252 |
| Corrected precision ↑ | Gastrointestinal | worse | 2.1e-10 | 0.067 |
| Corrected precision ↑ | Trauma | better | 1.84e-27 | 0.114 |
| Corrected precision ↑ | Metabolic | better | 1.03e-25 | 0.134 |
| Corrected NPV ↑ | Cardiovascular | better | 1.28e-34 | 0.019 |
| Corrected NPV ↑ | Neurological | better | 1.28e-34 | 0.016 |
| Corrected NPV ↑ | Gastrointestinal | better | 1.28e-34 | 0.019 |
| Corrected NPV ↑ | Respiratory | better | 1.28e-34 | 0.018 |
| Corrected NPV ↑ | Other | better | 1.28e-34 | 0.018 |
| Corrected NPV ↑ | Trauma | better | 1.28e-34 | 0.018 |
| Corrected NPV ↑ | Metabolic | better | 1.28e-34 | 0.018 |
| Event-based recall ↑ | Cardiovascular | better | 1.32e-34 | 0.081 |
| Event-based recall ↑ | Neurological | worse | 1.28e-34 | 0.346 |
| Event-based recall ↑ | Gastrointestinal | better | 5.90e-10 | 0.02 |
| Event-based recall ↑ | Other | better | 3.13e-13 | 0.035 |
| Event-based recall ↑ | Metabolic | worse | 1.55e-13 | 0.088 |
| Calibration error ↓ | Cardiovascular | better | 2.11e-11 | 0.01 |
| Calibration error ↓ | Neurological | worse | 1.28e-34 | 0.096 |
| Calibration error ↓ | Gastrointestinal | worse | 3.38e-30 | 0.032 |
| Calibration error ↓ | Respiratory | worse | 5.88e-33 | 0.041 |
| Calibration error ↓ | Other | worse | 4.93e-33 | 0.04 |
| Calibration error ↓ | Trauma | worse | 1.28e-34 | 0.056 |
| Calibration error ↓ | Metabolic | worse | 1.28e-34 | 0.09 |
| Avg. score on positive class | Cardiovascular | better | 1.58e-34 | 0.035 |
| Avg. score on positive class | Neurological | worse | 1.28e-34 | 0.133 |
| Avg. score on positive class | Gastrointestinal | better | 1.94e-32 | 0.043 |
| Avg. score on positive class | Respiratory | worse | 1.57e-27 | 0.026 |
| Avg. score on negative class | Cardiovascular | worse | 1.28e-34 | 0.031 |
| Avg. score on negative class | Neurological | better | 1.28e-34 | 0.032 |
| Avg. score on negative class | Gastrointestinal | worse | 1.28e-34 | 0.013 |
| Avg. score on negative class | Respiratory | worse | 1.71e-10 | 0.002 |
| Avg. score on negative class | Other | worse | 3.15e-34 | 0.009 |
| Avg. score on negative class | Trauma | better | 1.28e-34 | 0.014 |
| AUROC ↑ | Cardiovascular | worse | 1.28e-34 | 0.045 |
| AUROC ↑ | Neurological | better | 2.e-29 | 0.023 |
| AUROC ↑ | Gastrointestinal | worse | 1.18e-12 | 0.011 |
| AUROC ↑ | Respiratory | worse | 2.99e-33 | 0.027 |
| AUROC ↑ | Other | worse | 1.03e-18 | 0.016 |
| AUROC ↑ | Trauma | better | 1.34e-30 | 0.033 |
| AUROC ↑ | Metabolic | better | 1.05e-10 | 0.019 |
| AUPRC ↑ | Cardiovascular | better | 8.15e-19 | 0.028 |
| AUPRC ↑ | Neurological | worse | 1.28e-34 | 0.233 |
| AUPRC ↑ | Gastrointestinal | better | 1.40e-18 | 0.062 |
| AUPRC ↑ | Metabolic | better | 4.27e-17 | 0.081 |
| Corrected AUPRC ↑ | Cardiovascular | worse | 1.53e-34 | 0.084 |
| Corrected AUPRC ↑ | Neurological | better | 1.28e-34 | 0.117 |
| Corrected AUPRC ↑ | Trauma | better | 1.63e-34 | 0.158 |
| Corrected AUPRC ↑ | Metabolic | better | 7.75e-17 | 0.072 |
| Event-based AUPRC ↑ | Neurological | worse | 1.28e-34 | 0.281 |
| Event-based AUPRC ↑ | Respiratory | better | 4.99e-15 | 0.047 |
| Event-based AUPRC ↑ | Metabolic | better | 1.99e-23 | 0.126 |
| Corrected event-based AUPRC ↑ | Cardiovascular | worse | 8.48e-32 | 0.079 |
| Corrected event-based AUPRC ↑ | Neurological | better | 8.35e-33 | 0.088 |
| Corrected event-based AUPRC ↑ | Respiratory | better | 5.11e-08 | 0.03 |
| Corrected event-based AUPRC ↑ | Trauma | better | 3.23e-26 | 0.095 |
| Corrected event-based AUPRC ↑ | Metabolic | better | 8.35e-23 | 0.101 |

**Figure 2.2.3.b.**
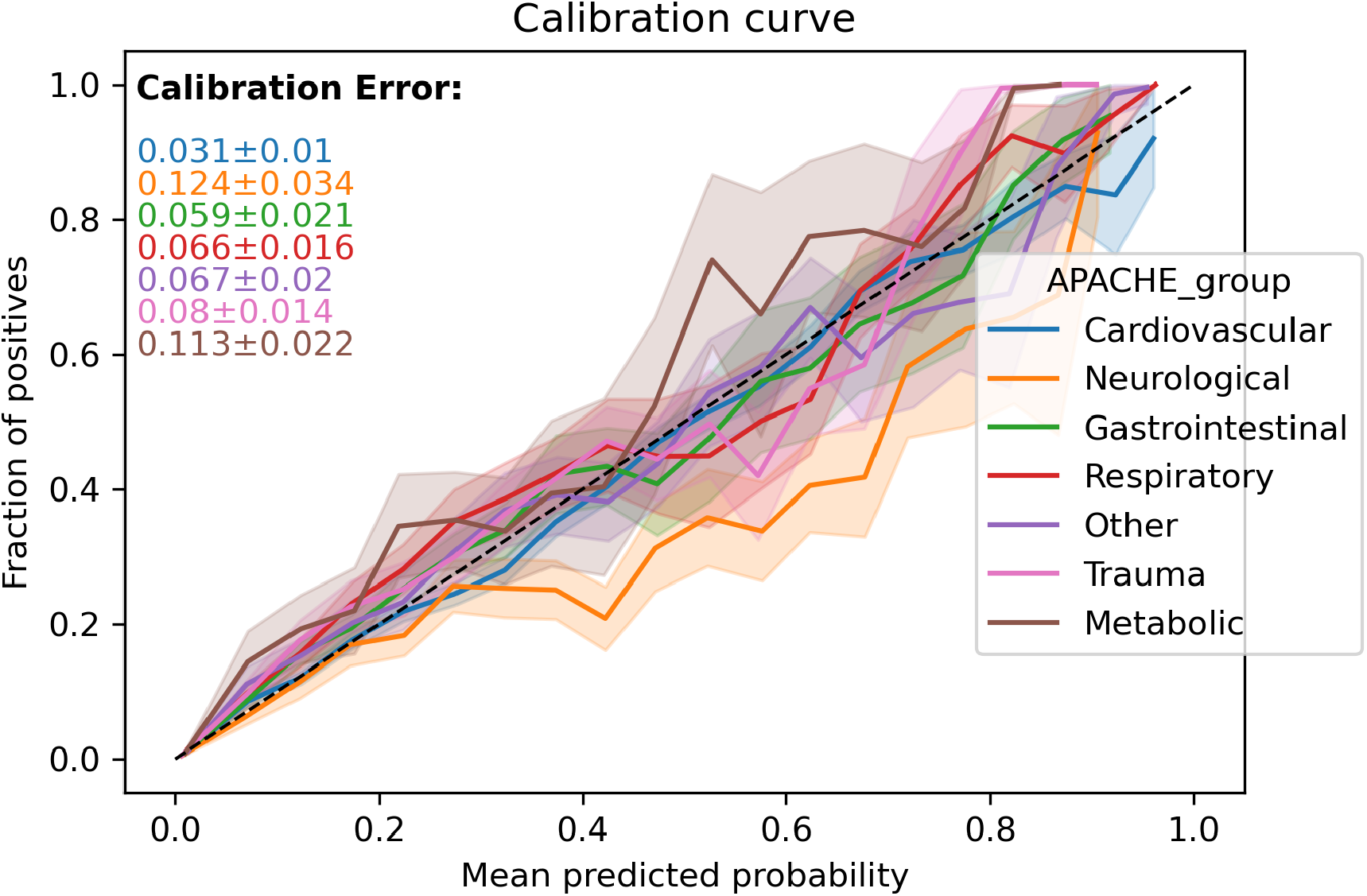

**Figure 2.2.3.c.**
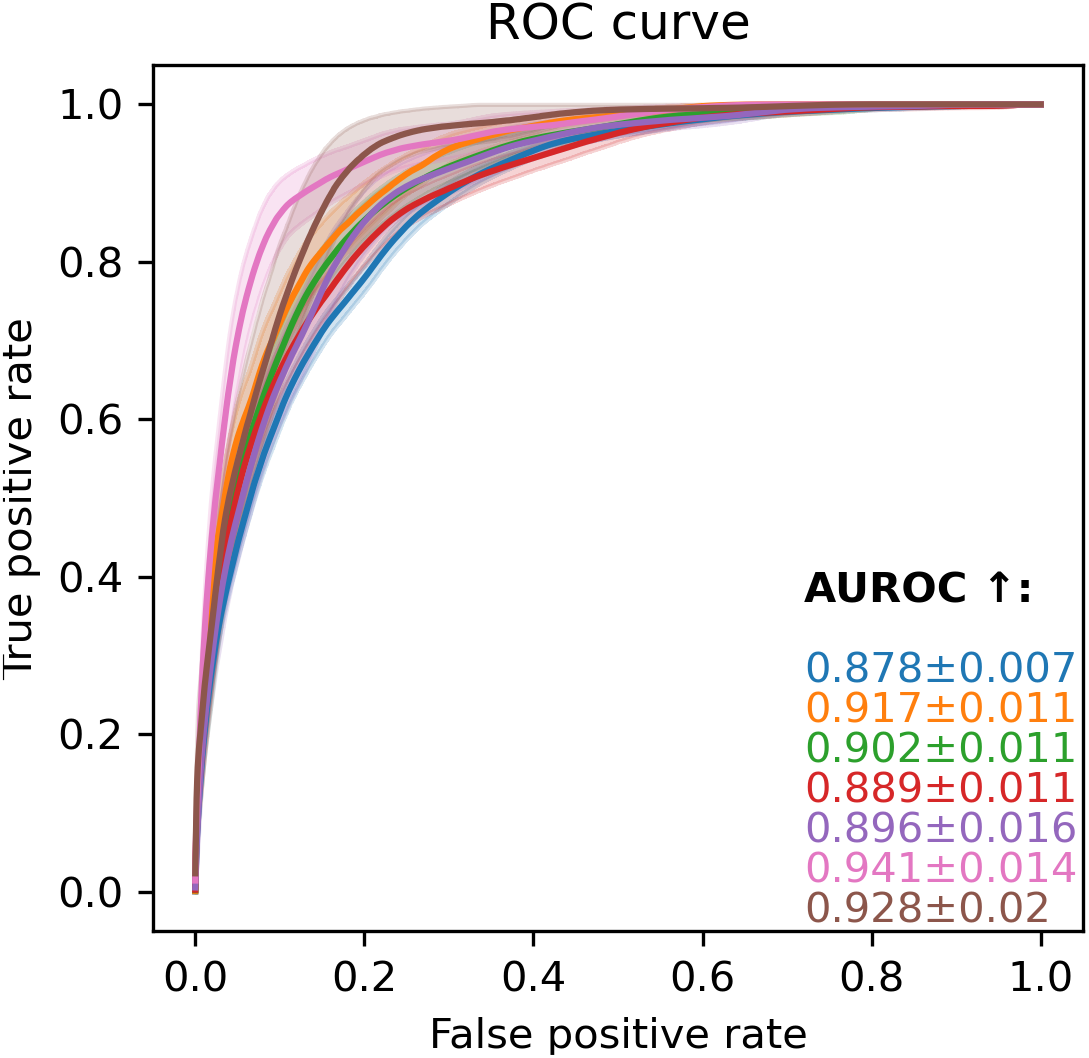

**Figure 2.2.3.d.**
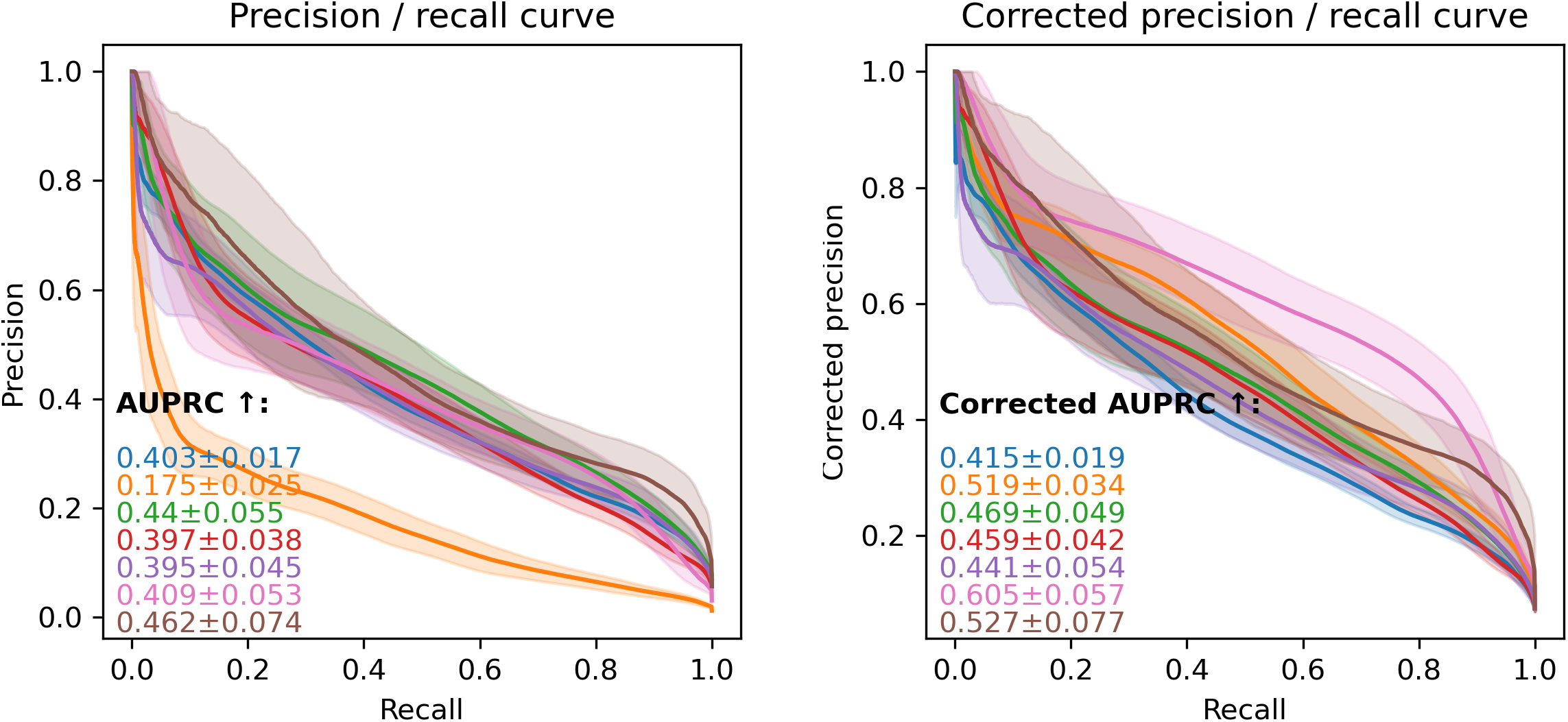
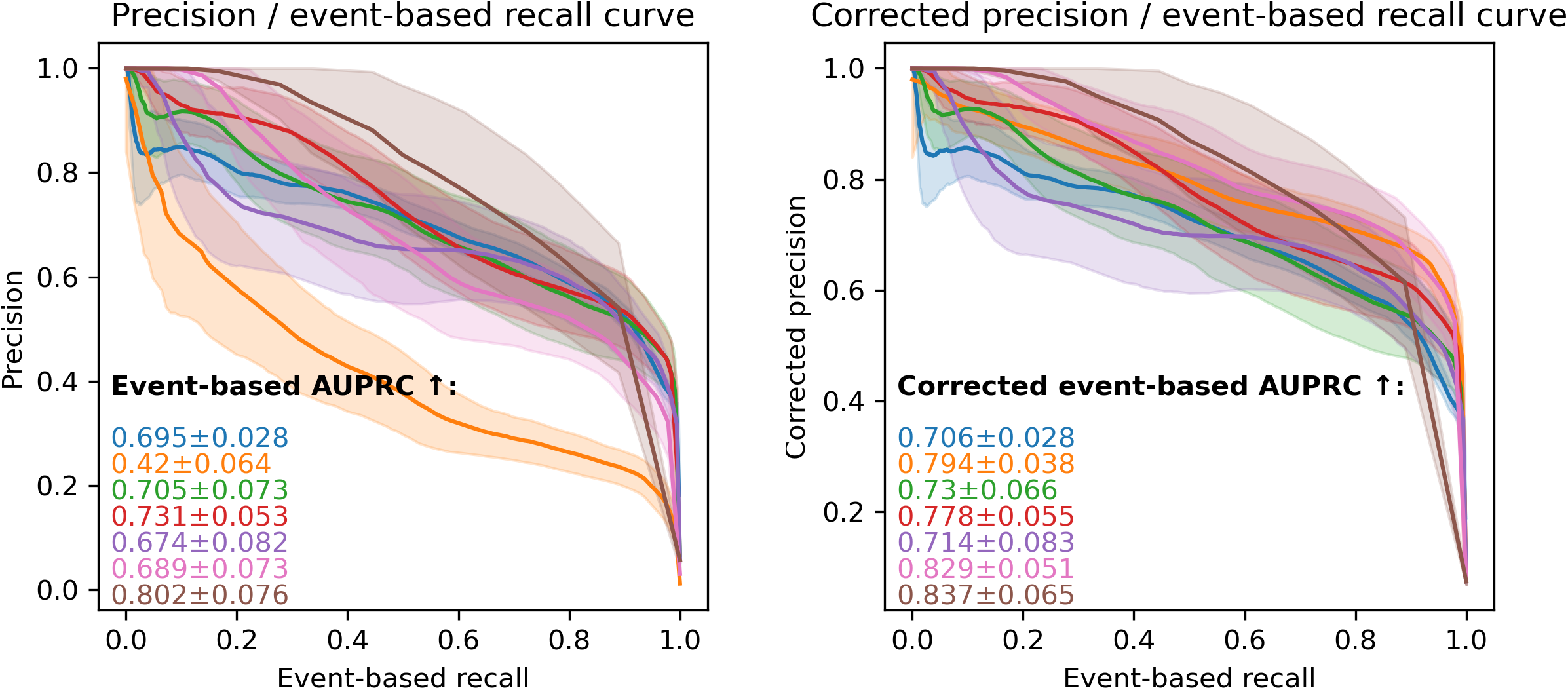

##### 2.2.4. … surgical_status

**Figure 2.2.4.a.**
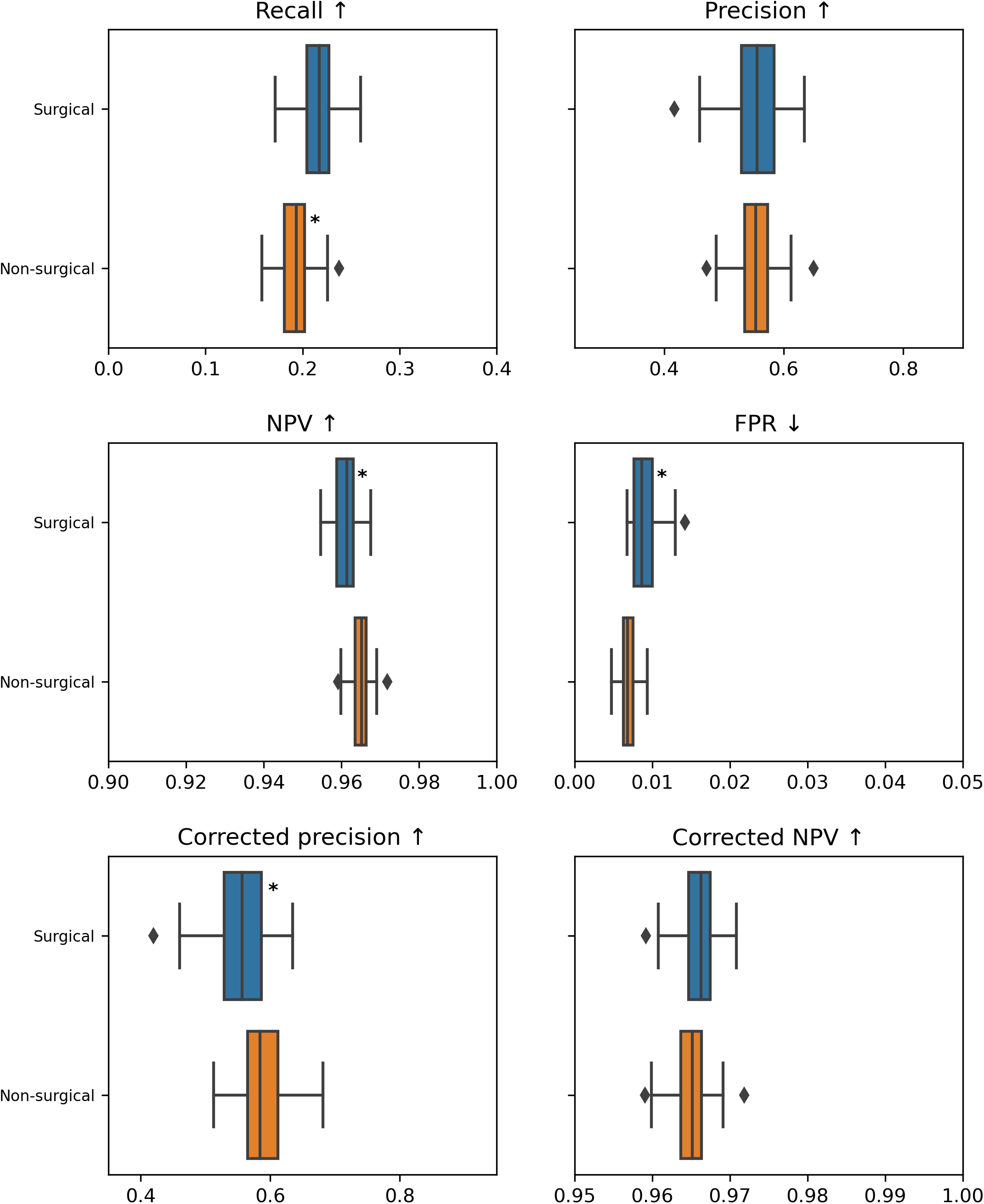
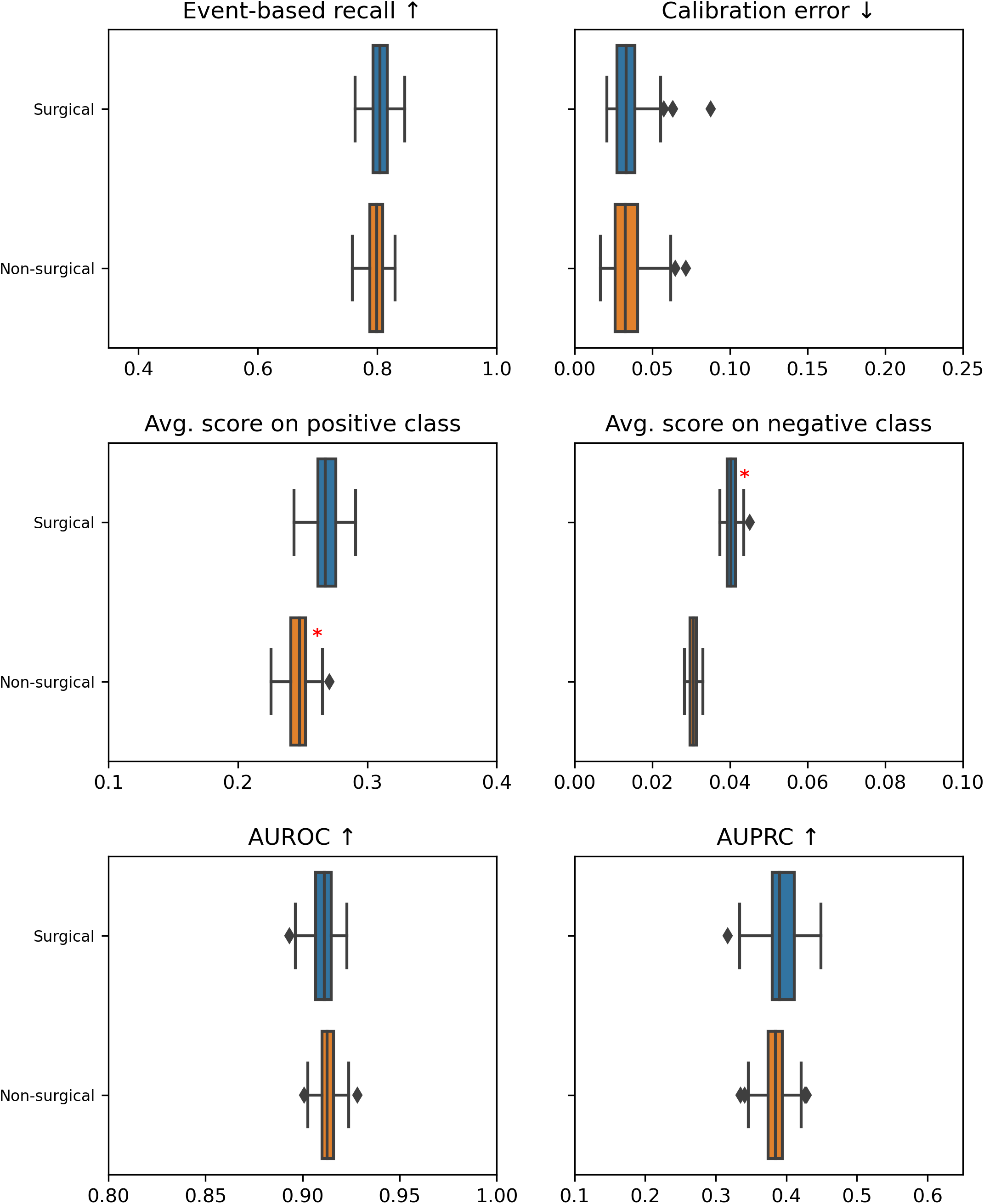
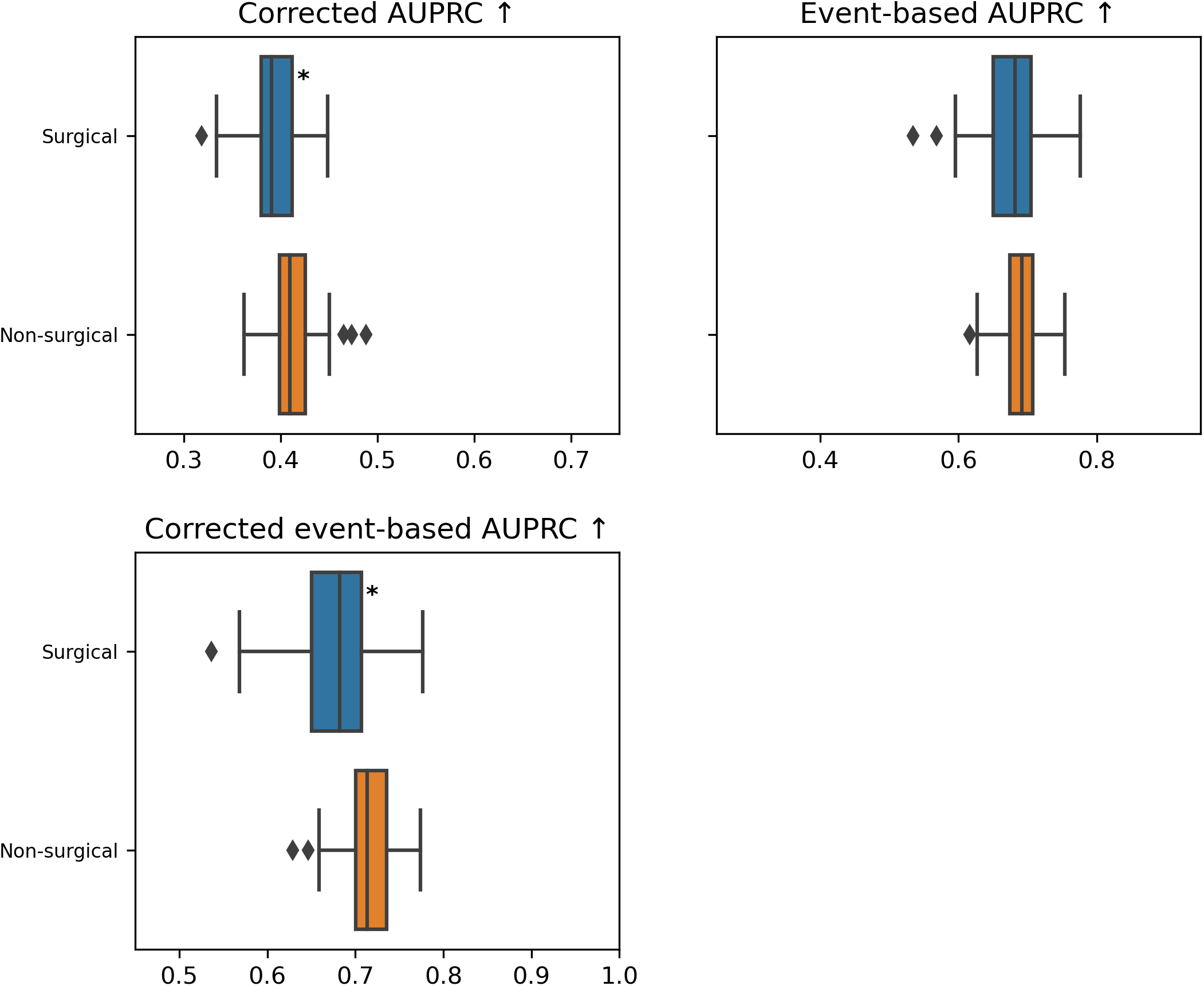

**Table 2.2.4.a.**

| Metric | Cohort with the worst metric | P-value | Delta |
| --- | --- | --- | --- |
| Recall ↑ | Non-surgical | 2.1e-19 | 0.024 |
| NPV ↑ | Surgical | 8.23e-18 | 0.004 |
| FPR ↓ | Surgical | 1.62e-21 | 0.002 |
| Corrected precision ↑ | Surgical | 1.57e-08 | 0.027 |
| Avg. score on positive class | Non-surgical | 3.41e-28 | 0.02 |
| Avg. score on negative class | Surgical | 1.28e-34 | 0.01 |
| Corrected AUPRC ↑ | Surgical | 6.08e-08 | 0.019 |
| Corrected event-based AUPRC ↑ | Surgical | 1.08e-09 | 0.031 |

**Figure 2.2.4.b.**
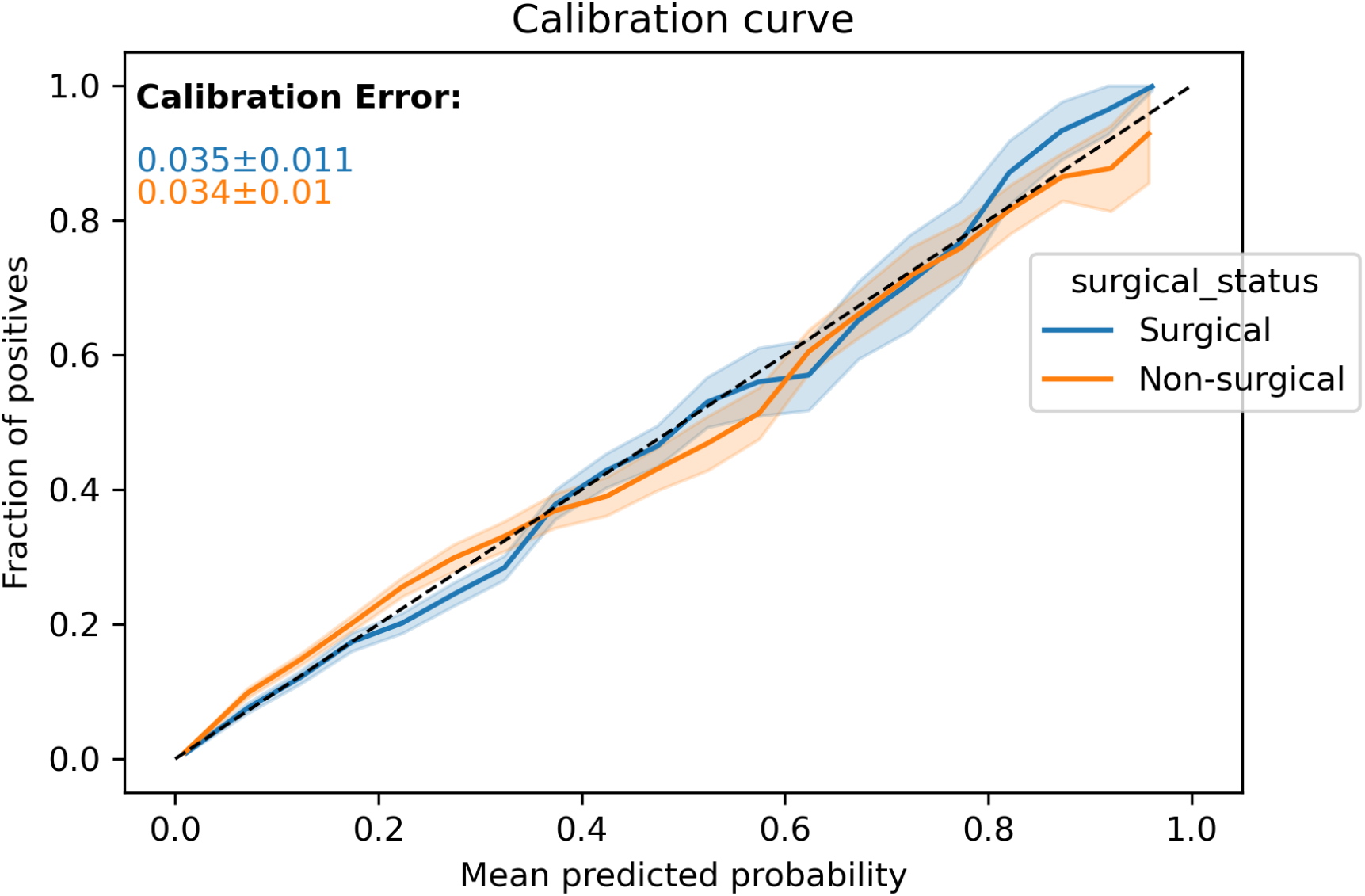

**Figure 2.2.4.c.**
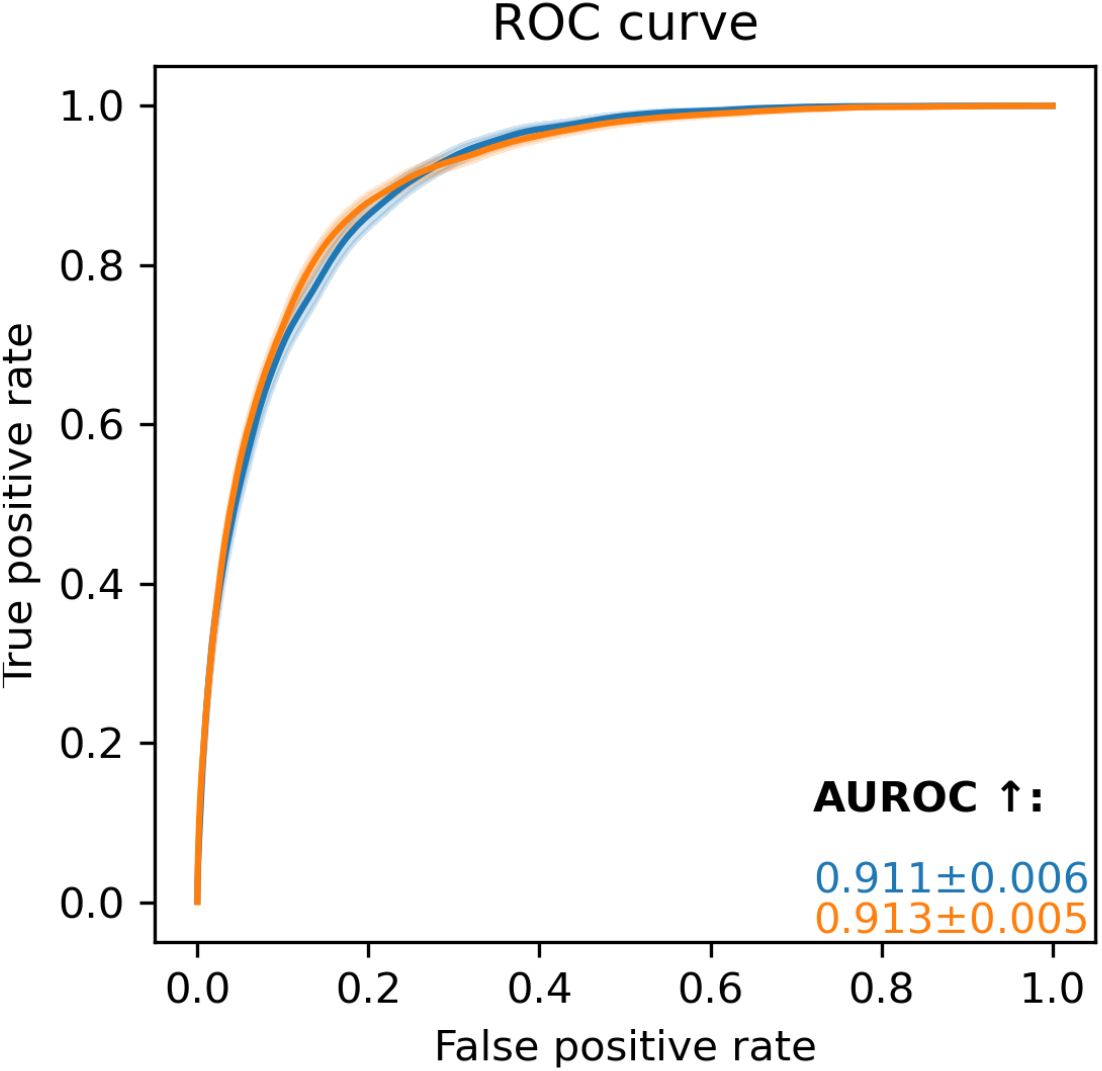

**Figure 2.2.4.d.**
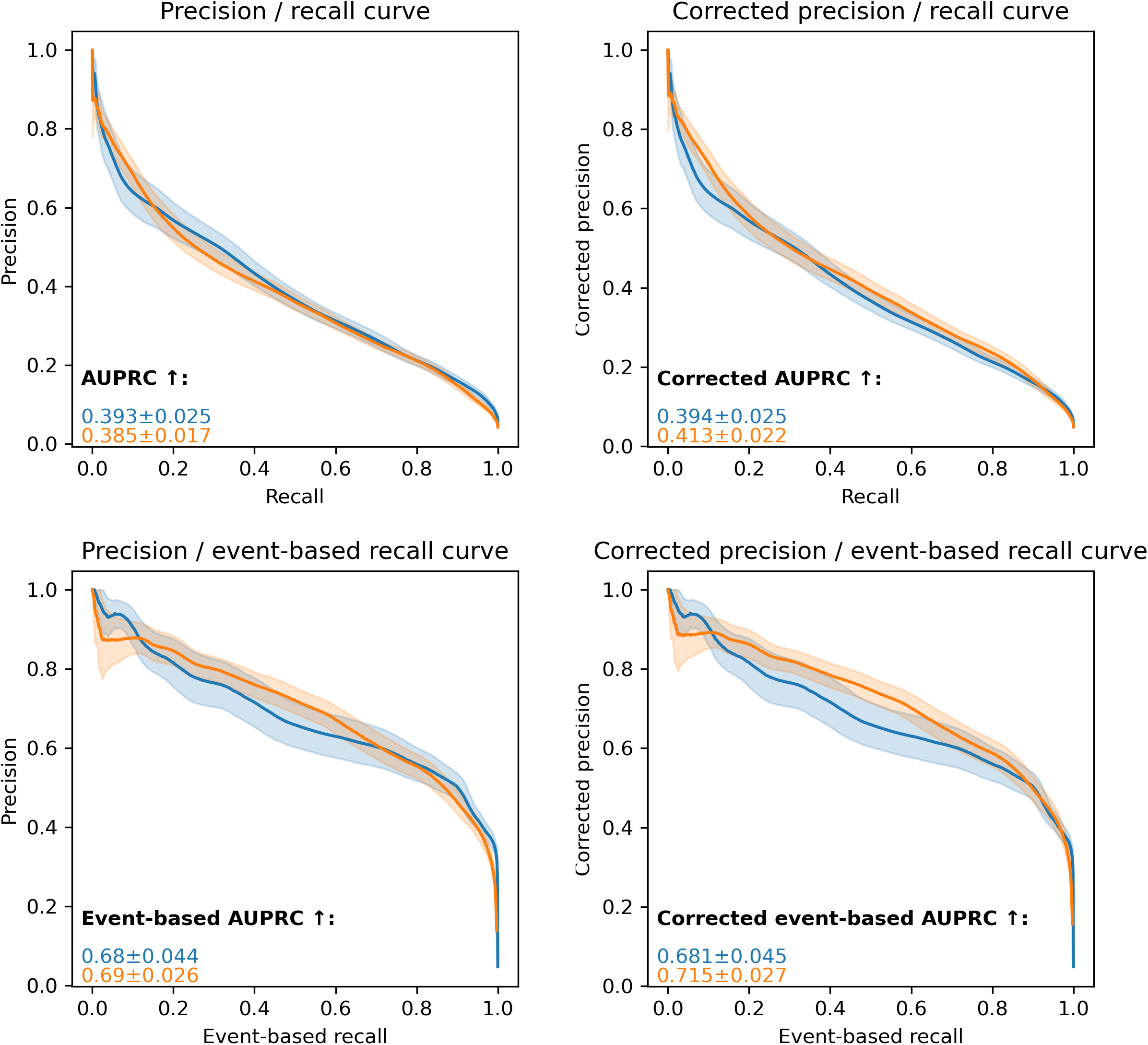

### 3. Time Gap Analysis

***Goal: Checking whether the time gap between the first correct alarm and the start of the corresponding event are similar across cohorts of patients***

#### 3.1. Aggregated views

##### 3.1.1. Summary statistics of median time gap per grouping

For event starting in the window 0-3h, the overall macro-averaged median time gap is 48.4 (in minutes).

For event starting in the window 3-6h, the overall macro-averaged median time gap is 218.2 (in minutes).

For event starting in the window 6-12h, the overall macro-averaged median time gap is 394.4 (in minutes).

For event starting in the window >12h, the overall macro-averaged median time gap is 66.6 (in minutes).

###### Grouping by sex

**Table 3.1.1.a.**

| Start event | Macro-average (in minutes) | Minimum (category) | For minority category |
| --- | --- | --- | --- |
| 0-3h | 47.5 | 45.0 (M) | 50.0 |
| 3-6h | 217.5 | 215.0 (F) | 215.0 |
| 6-12h | 405.0 | 395.0 (M) | 415.0 |
| >12h | 30.625 | 26.25 (F) | 26.25 |

###### Grouping by age_group

**Table 3.1.1.b.**

| Start event | Macro-average (in minutes) | Minimum (category) | For minority category |
| --- | --- | --- | --- |
| 0-3h | 43.0 | 25.0 (>85) | 25.0 |
| 3-6h | 216.25 | 210.0 (50-65) | 215.0 |
| 6-12h | 403.0 | 375.0 (<50) | 432.5 |
| >12h | 38.5 | 20.0 (75-85) | 55.0 |

###### Grouping by APACHE_group

**Table 3.1.1.c.**

| Start event | Macro-average (in minutes) | Minimum (category) | For minority category |
| --- | --- | --- | --- |
| 0-3h | 55.357 | 35.0 (Neurological) | 90.0 |
| 3-6h | 220.893 | 190.0 (Respiratory) | 255.0 |
| 6-12h | 367.143 | 157.5 (Neurological) | 370.0 |
| >12h | 170.357 | 20.0 (Cardiovascular) | 140.0 |

###### Grouping by surgical_status

**Table 3.1.1.d.**

| Start event | Macro-average (in minutes) | Minimum (category) | For minority category |
| --- | --- | --- | --- |
| 0-3h | 47.5 | 45.0 (Surgical) | 45.0 |
| 3-6h | 217.5 | 215.0 (Surgical) | 215.0 |
| 6-12h | 405.0 | 400.0 (Surgical) | 400.0 |
| >12h | 32.5 | 30.0 (Non-surgical) | 35.0 |

##### 3.1.2. Top 3 cohorts with the biggest time gap discrepancies

In the following table, we show for each start of the event window the 3 cohorts with the biggest delta that are significantly worse off than the rest of the patients. If some cells are empty, this means that there are fewer than 3 cohorts, possibly none, that are significantly worse than the rest of the patients for this particular start of the event window.

**Table 3.1.2.a.**

| Start event | Cohort 1 ($\Delta$ in minutes) | Cohort 2 ($\Delta$ in minutes) | Cohort 3 ($\Delta$ in minutes) |
| --- | --- | --- | --- |
| 0-3h | >85 (25.0) | Neurological (15.0) | Cardiovascular (5.0) |
| 3-6h | Respiratory (30.0) | Other (15.0) | 50-65 (10.0) |
| 6-12h | Neurological (247.5) | Trauma (40.0) | <50 (30.0) |
| >12h | Cardiovascular (70.0) | 75-85 (17.5) | Other (12.5) |

#### 3.2. Grouping by

For each grouping, we display box plots that show the median time gap between alarm and event for the different categories of patients depending on the period of the stay when the event began. For each start of event window, we emphasize with a black star the cohorts that are significantly worse off compared to the rest of the patients and with a red star the cohorts that appear in the table **Top 3 cohorts with the biggest time gap discrepancies**.

For each grouping, we propose a table that presents the results of the statistical analysis: comparing the time gap from alarm to event for one cohort against the rest of the patients. P-values are obtained by running the Mann-Whitney U test with Bonferroni correction. We display only start of event windows and cohorts with a significant p-value (smaller than 0.001/number of comparisons) and whose delta is bigger than 0. For binary grouping, we display the category with the worst time gap distribution for each start of event window. While for multicategorical grouping we display whether the distribution for the category is better or worse than for the rest of patients

##### 3.2.1. … sex

**Figure 3.2.1.a.**
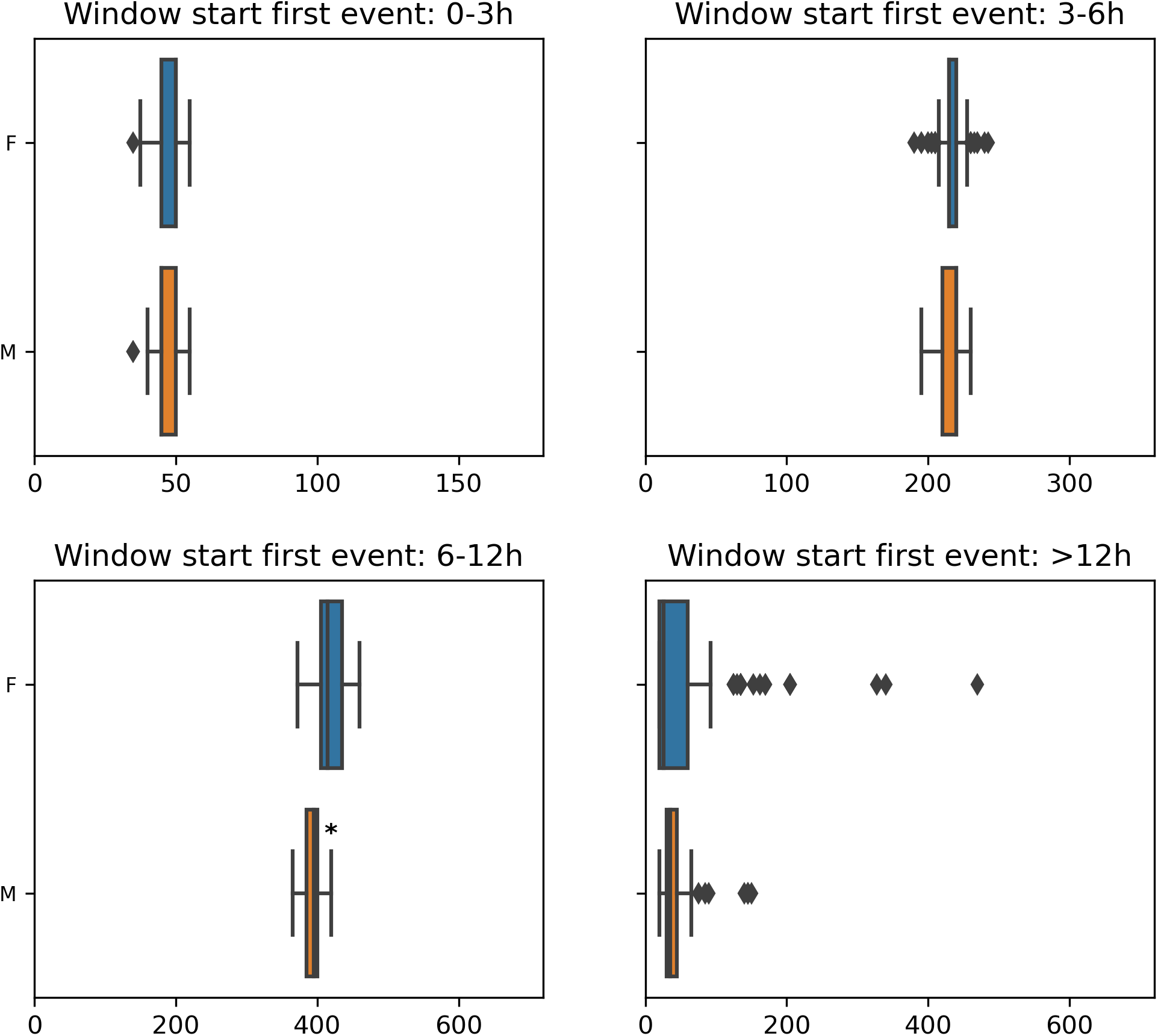

**Table 3.2.1.a.**
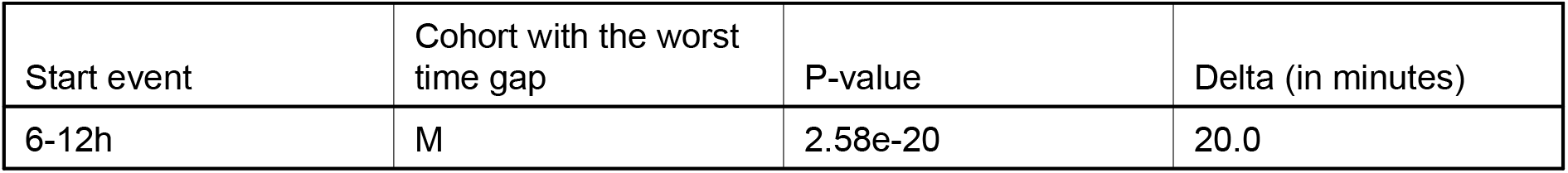

| Start event | Cohort with the worst time gap | P-value | Delta (in minutes) |
| --- | --- | --- | --- |
| 6-12h | M | 2.58e-20 | 20.0 |

##### 3.2.2. … age_group

**Figure 3.2.2.a.**
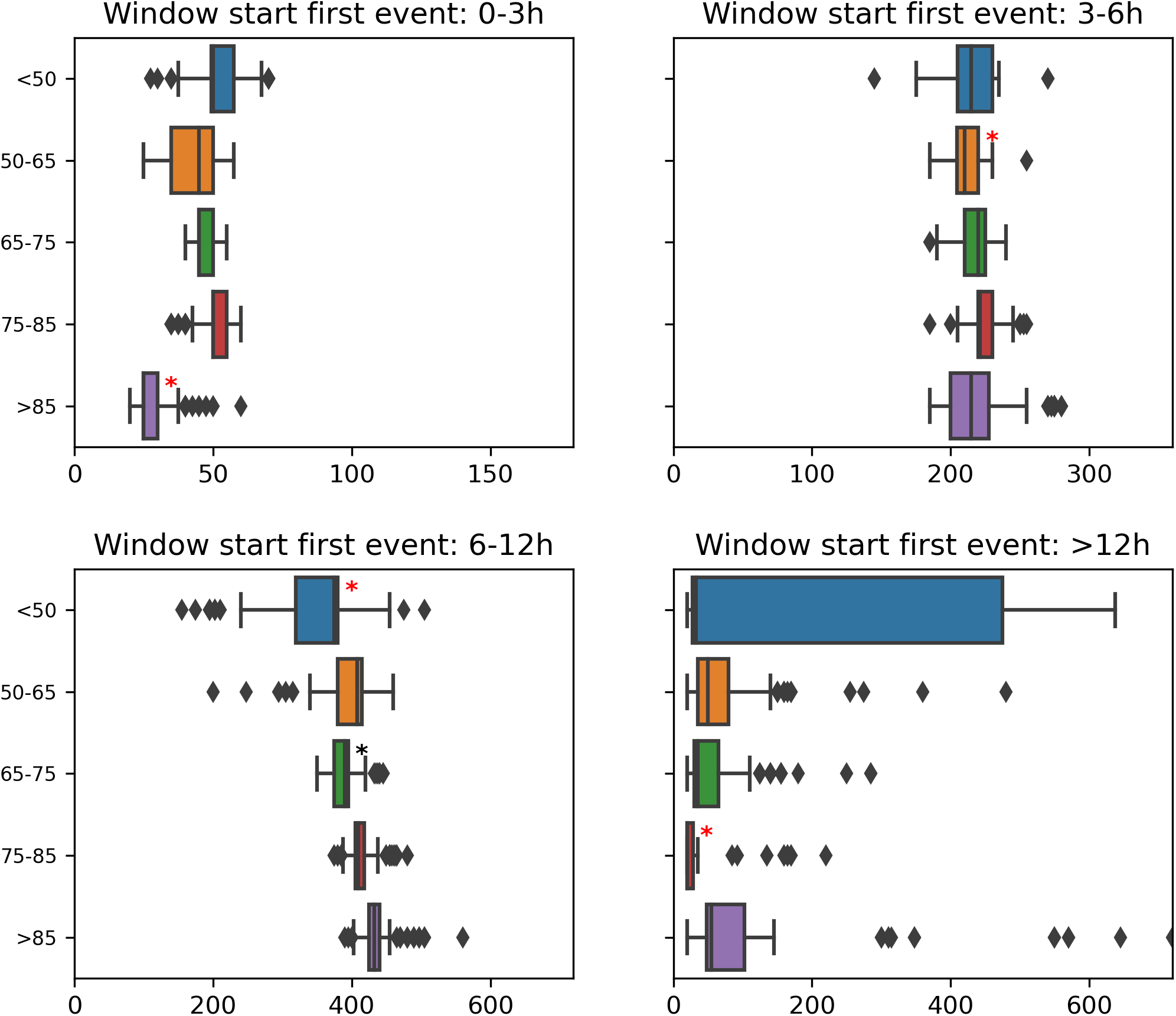

**Table 3.2.2.a.**

| Start event | Category | Cohort vs. rest | P-value | Delta (in minutes) |
| --- | --- | --- | --- | --- |
| 0-3h | <50 | better | 1.48e-12 | 5.0 |
| 0-3h | 75-85 | better | 6.25e-11 | 5.0 |
| 0-3h | >85 | worse | 3.53e-32 | 25.0 |
| 3-6h | 50-65 | worse | 6.9e-09 | 10.0 |
| 3-6h | 75-85 | better | 3.18e-11 | 6.25 |
| 6-12h | <50 | worse | 2.1e-20 | 30.0 |
| 6-12h | 65-75 | worse | 1.1e-16 | 20.0 |
| 6-12h | 75-85 | better | 1.05e-09 | 15.0 |
| 6-12h | >85 | better | 2.75e-29 | 37.5 |
| >12h | 50-65 | better | 3.62e-13 | 20.0 |
| >12h | 75-85 | worse | 4.78e-18 | 17.5 |
| >12h | >85 | better | 7.04e-16 | 25.0 |

##### 3.2.3. … APACHE_group

**Figure 3.2.3.a.**
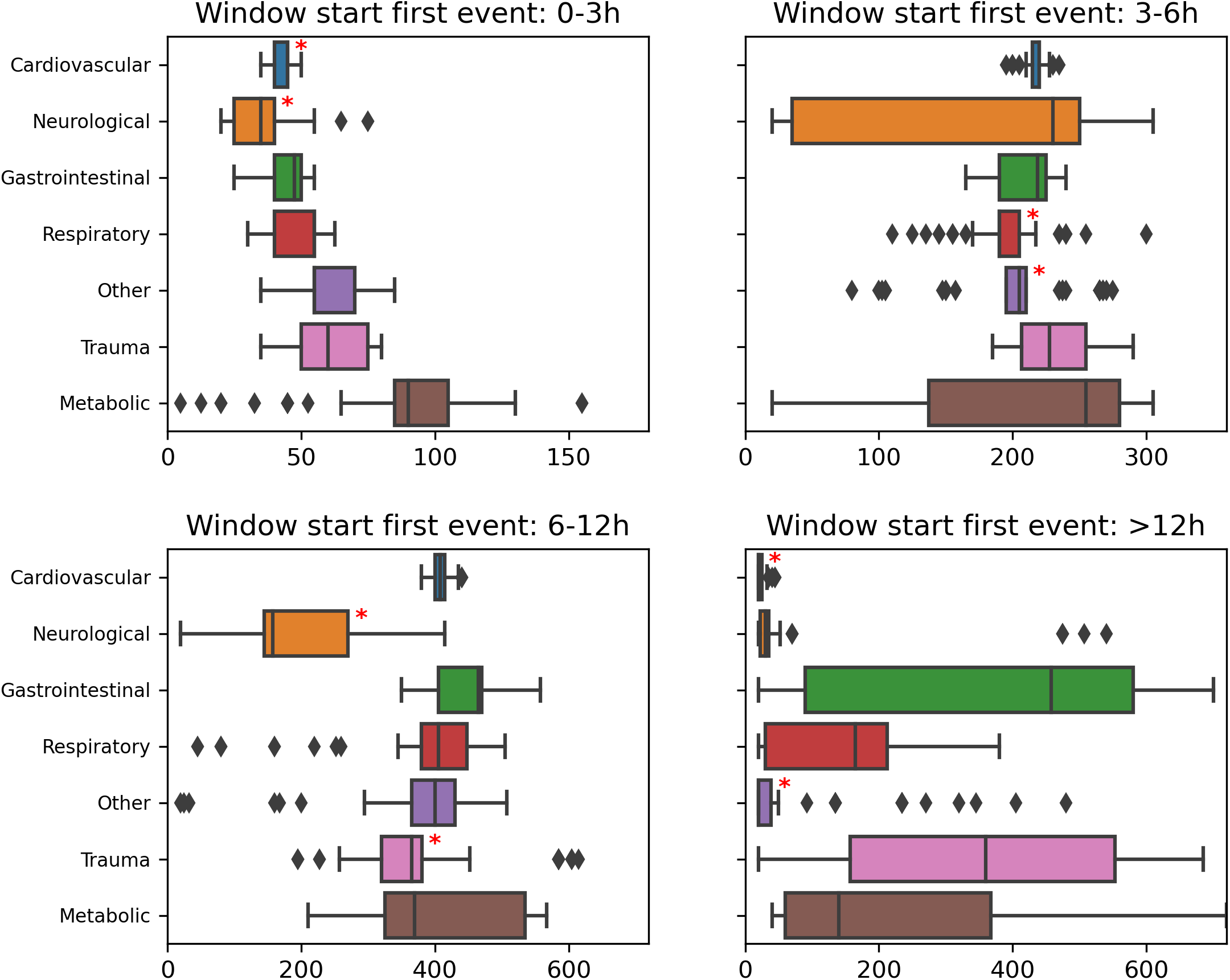

**Table 3.2.3.a.**

| Start event | Category | Cohort vs. rest | P-value | Delta (in minutes) |
| --- | --- | --- | --- | --- |
| 0-3h | Cardiovascular | worse | 1.71e-14 | 5.0 |
| 0-3h | Neurological | worse | 4.61e-13 | 15.0 |
| 0-3h | Respiratory | better | 7.03e-06 | 10.0 |
| 0-3h | Other | better | 3.51e-18 | 10.0 |
| 0-3h | Trauma | better | 1.58e-14 | 15.0 |
| 0-3h | Metabolic | better | 1.69e-21 | 45.0 |
| 3-6h | Cardiovascular | better | 1.99e-06 | 10.0 |
| 3-6h | Respiratory | worse | 5.34e-24 | 30.0 |
| 3-6h | Other | worse | 3.59e-11 | 15.0 |
| 6-12h | Cardiovascular | better | 1.19e-09 | 16.25 |
| 6-12h | Neurological | worse | 1.28e-25 | 247.5 |
| 6-12h | Gastrointestinal | better | 1.68e-12 | 65.0 |
| 6-12h | Trauma | worse | 1.23e-18 | 40.0 |
| >12h | Cardiovascular | worse | 1.72e-33 | 70.0 |
| >12h | Gastrointestinal | better | 1.83e-26 | 427.5 |
| >12h | Respiratory | better | 1.4e-08 | 135.0 |
| >12h | Other | worse | 1.19e-08 | 12.5 |
| >12h | Trauma | better | 6.72e-31 | 330.0 |
| >12h | Metabolic | better | 9.84e-30 | 110.0 |

##### 3.2.4. … surgical_status

**Figure 3.2.4.a.**
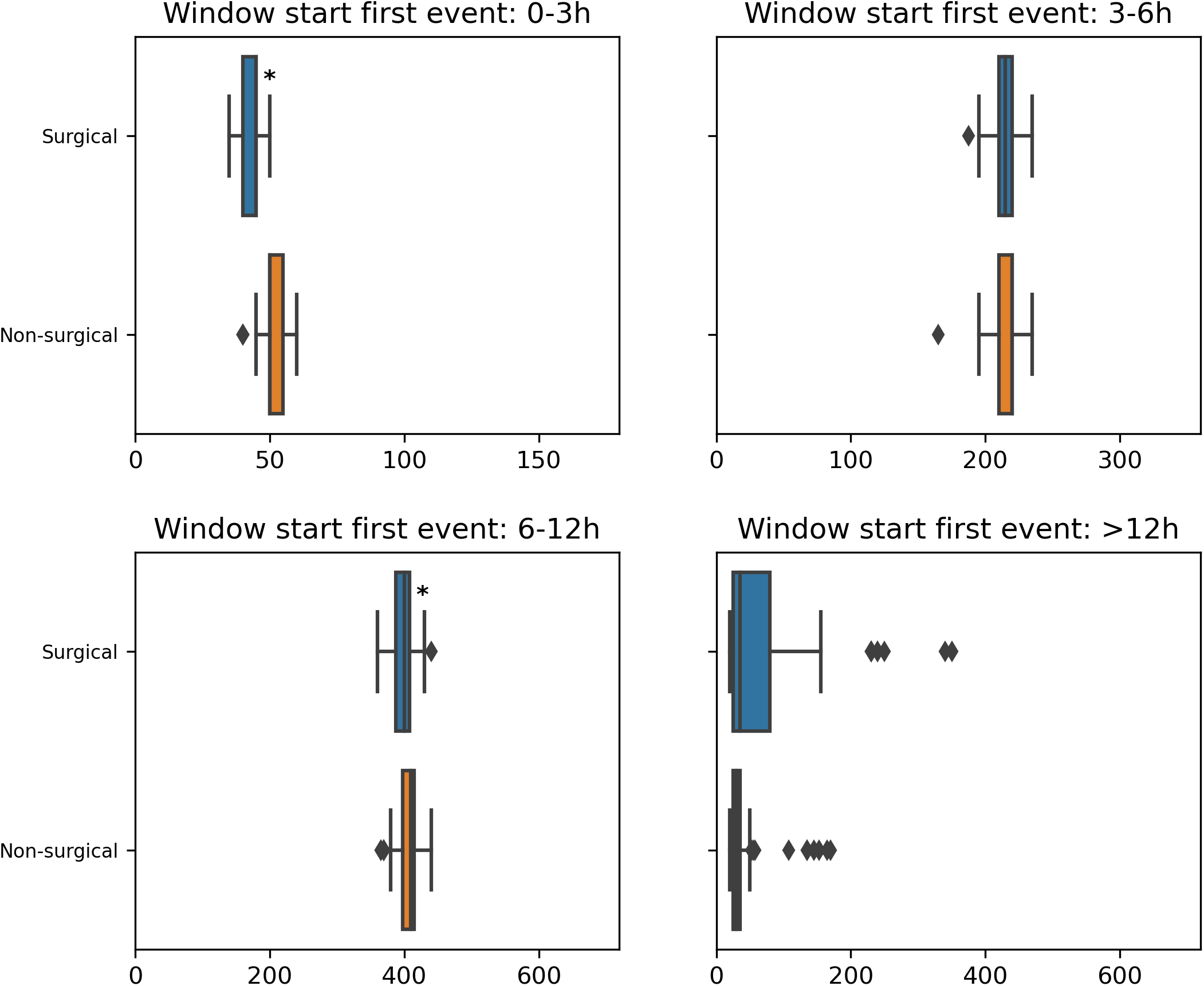

**Table 3.2.4.a.**

| Start event | Cohort with the worst time gap | P-value | Delta (in minutes) |
| --- | --- | --- | --- |
| 0-3h | Surgical | 5.81e-23 | 5.0 |
| 6-12h | Surgical | 5.46e-05 | 10.0 |

### 4. Medical Variable Analysis

***Goal: Comparing the median value of relevant medical variables across cohorts***

We check the following variables: a_Lac, ABPm

#### 4.1. Aggregated views

##### 4.1.1. Top 3 cohorts with the biggest differences in the medical variables distributions

In the following table, for each of the selected medical variables and median computation condition, we show the 3 cohorts with the biggest delta that are significantly different than the rest of the patients. If some cells are empty, that means that there are less than 3 cohorts (possibly none) that are significantly different than the rest of the patients for this particular medical variable and median computation condition.

**Table 4.1.1.a.**
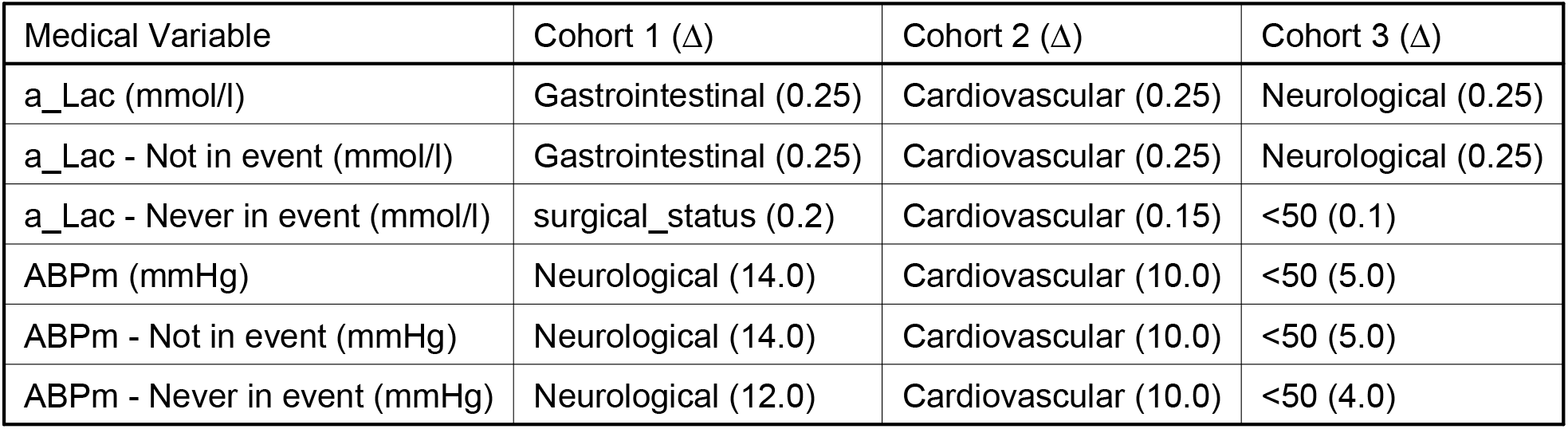

#### 4.2. Grouping by

For each grouping, we display box plots that show the median value of the selected medical variables for three conditions: all time points during the entire stay, time points while not in an event, and time points from patients not experiencing any event. For each variable and condition, we emphasize with a black star the cohorts that are significantly different compared to the rest of the patients and with a red star the cohorts that appear in the table **Top 3 cohorts with the biggest differences in the medical variables values**.

For each grouping, we propose a table that presents the results of the statistical analysis: comparing the medical variables’ median value for one cohort against the rest of the patients. P-values are obtained by running the Mann-Whitney U test with Bonferroni correction. We display only medical variables and cohorts with a significant p-value (smaller than 0.001/number of comparisons) and whose delta is bigger than 0. For binary grouping, we display the category with the greatest median value for each of the selected medical variables and median computation condition. While for multicategorical grouping we display whether the median value for the category is greater or less than for the rest of patients

##### 4.2.1. … sex

**Figure 4.2.1.a.**
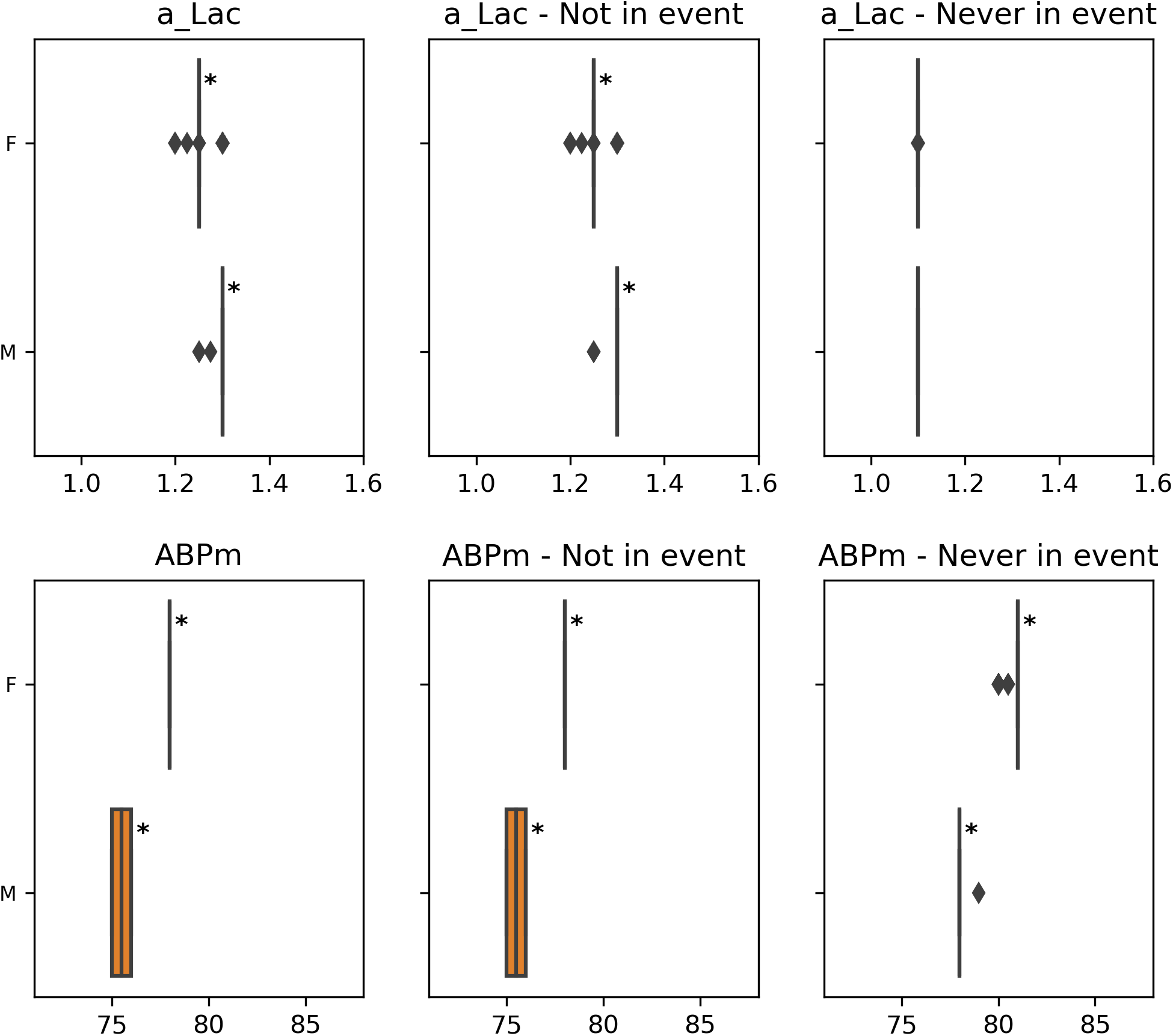

**Table 4.2.1.a.**

| Medical Variable | Cohort with greater median value | P-value | Delta |
| --- | --- | --- | --- |
| a_Lac | M | 4.79e-30 | 0.05 |
| a_Lac - Not in event | M | 4.95e-30 | 0.05 |
| ABPm | F | 3.98e-40 | 2.5 |
| ABPm - Not in event | F | 3.77e-40 | 2.5 |
| ABPm - Never in event | F | 2.97e-43 | 3.0 |

##### 4.2.2. … age_group

**Figure 4.2.2.a.**
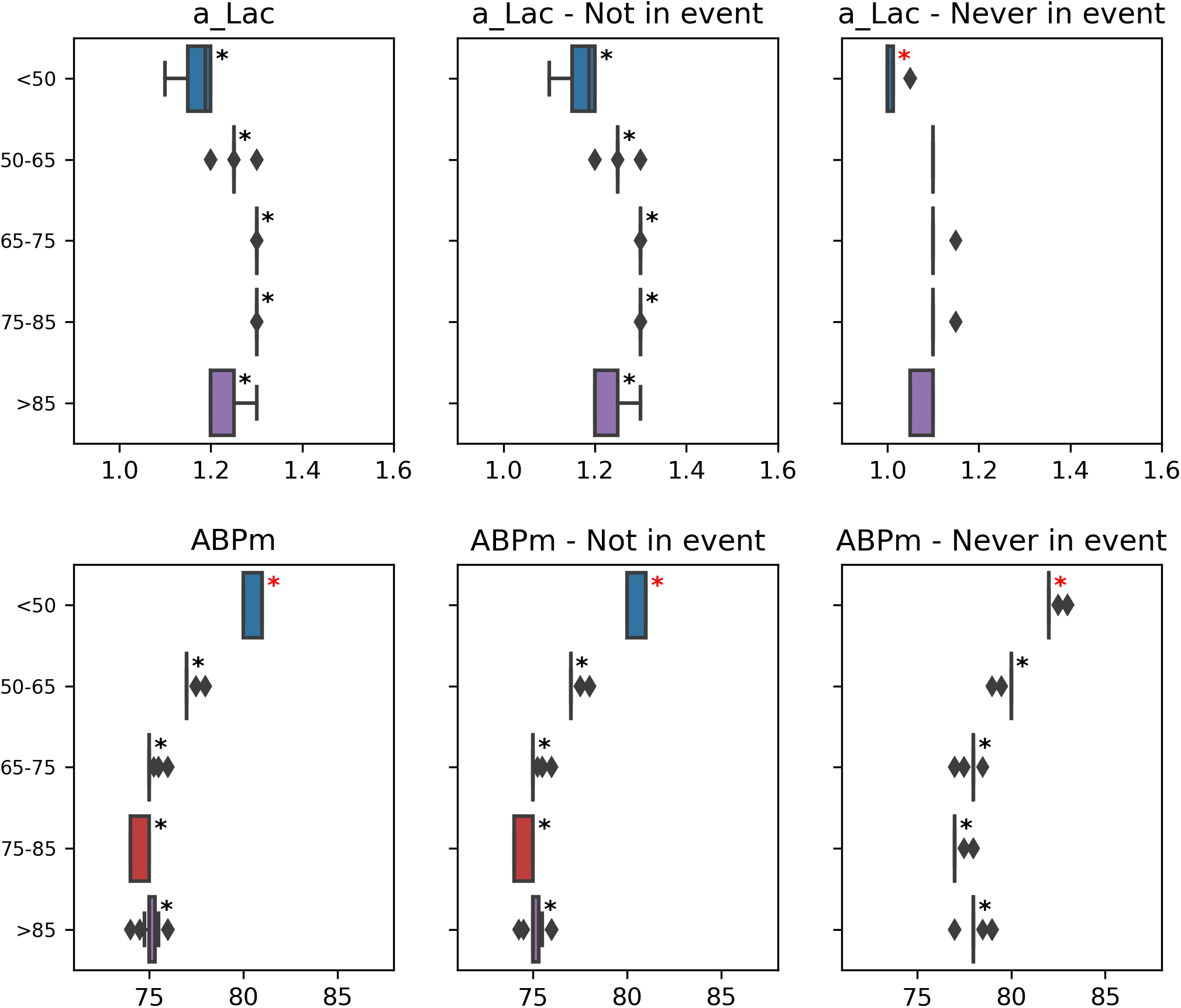

**Table 4.2.2.a.**
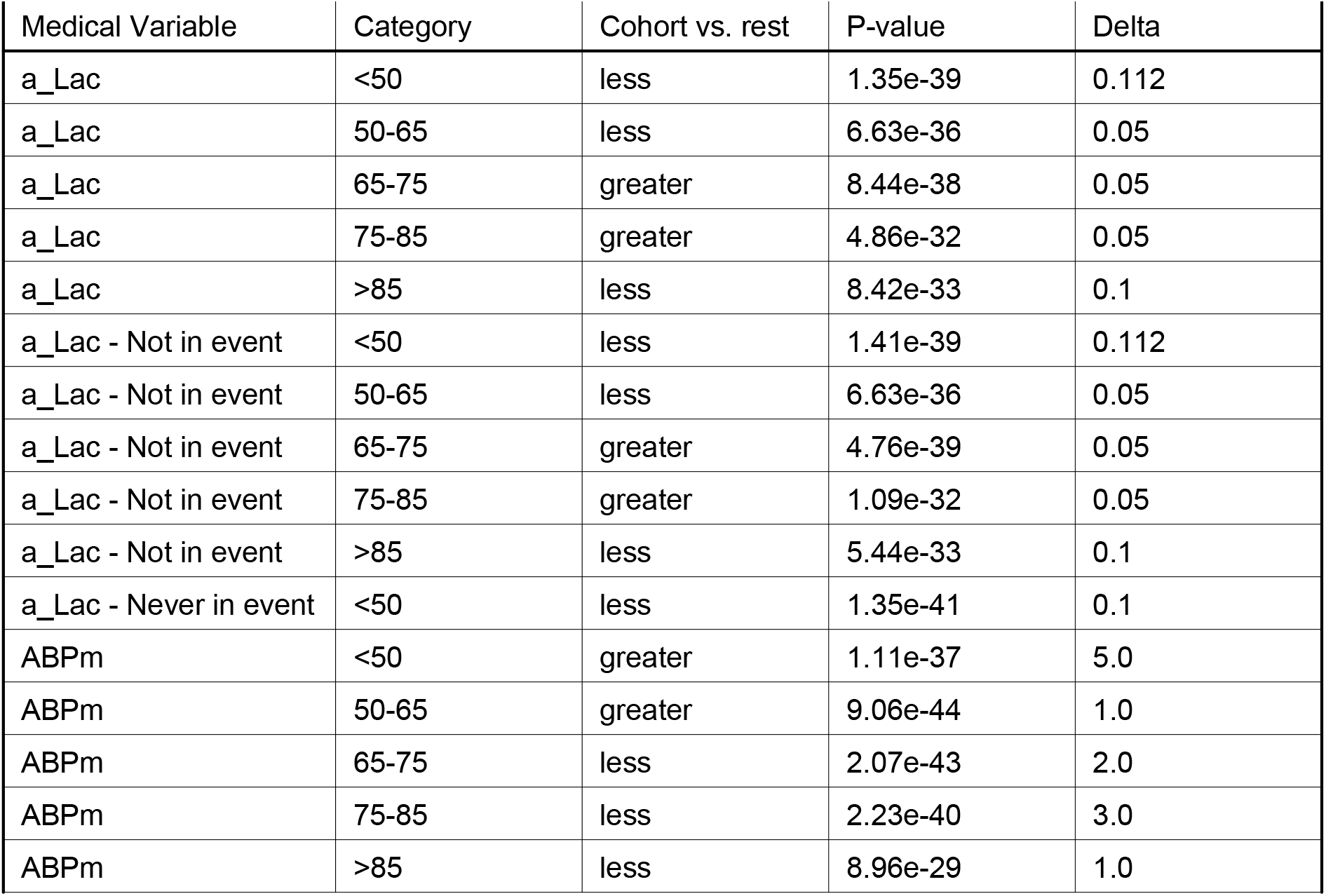
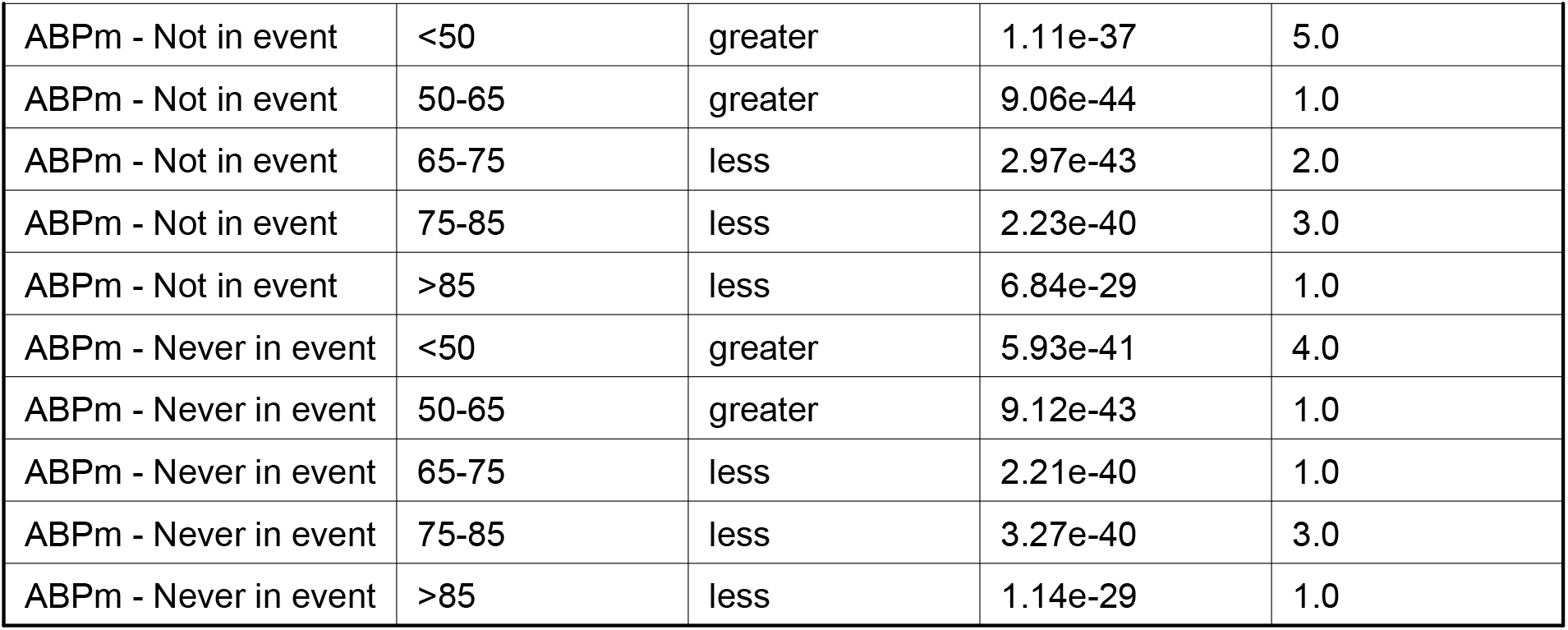

| Medical Variable | Category | Cohort vs. rest | P-value | Delta |
| --- | --- | --- | --- | --- |
| a_Lac | <50 | less | 1.35e-39 | 0.112 |
| a_Lac | 50-65 | less | 6.63e-36 | 0.05 |
| a_Lac | 65-75 | greater | 8.44e-38 | 0.05 |
| a_Lac | 75-85 | greater | 4.86e-32 | 0.05 |
| a_Lac | >85 | less | 8.42e-33 | 0.1 |
| a_Lac - Not in event | <50 | less | 1.41e-39 | 0.112 |
| a_Lac - Not in event | 50-65 | less | 6.63e-36 | 0.05 |
| a_Lac - Not in event | 65-75 | greater | 4.76e-39 | 0.05 |
| a_Lac - Not in event | 75-85 | greater | 1.09e-32 | 0.05 |
| a_Lac - Not in event | >85 | less | 5.44e-33 | 0.1 |
| a_Lac - Never in event | <50 | less | 1.35e-41 | 0.1 |
| ABPm | <50 | greater | 1.11e-37 | 5.0 |
| ABPm | 50-65 | greater | 9.06e-44 | 1.0 |
| ABPm | 65-75 | less | 2.07e-43 | 2.0 |
| ABPm | 75-85 | less | 2.23e-40 | 3.0 |
| ABPm | >85 | less | 8.96e-29 | 1.0 |
| ABPm - Not in event | <50 | greater | 1.11e-37 | 5.0 |
| ABPm - Not in event | 50-65 | greater | 9.06e-44 | 1.0 |
| ABPm - Not in event | 65-75 | less | 2.97e-43 | 2.0 |
| ABPm - Not in event | 75-85 | less | 2.23e-40 | 3.0 |
| ABPm - Not in event | >85 | less | 6.84e-29 | 1.0 |
| ABPm - Never in event | <50 | greater | 5.93e-41 | 4.0 |
| ABPm - Never in event | 50-65 | greater | 9.12e-43 | 1.0 |
| ABPm - Never in event | 65-75 | less | 2.21e-40 | 1.0 |
| ABPm - Never in event | 75-85 | less | 3.27e-40 | 3.0 |
| ABPm - Never in event | >85 | less | 1.14e-29 | 1.0 |

##### 4.2.3. … APACHE_group

**Figure 4.2.3.a.**
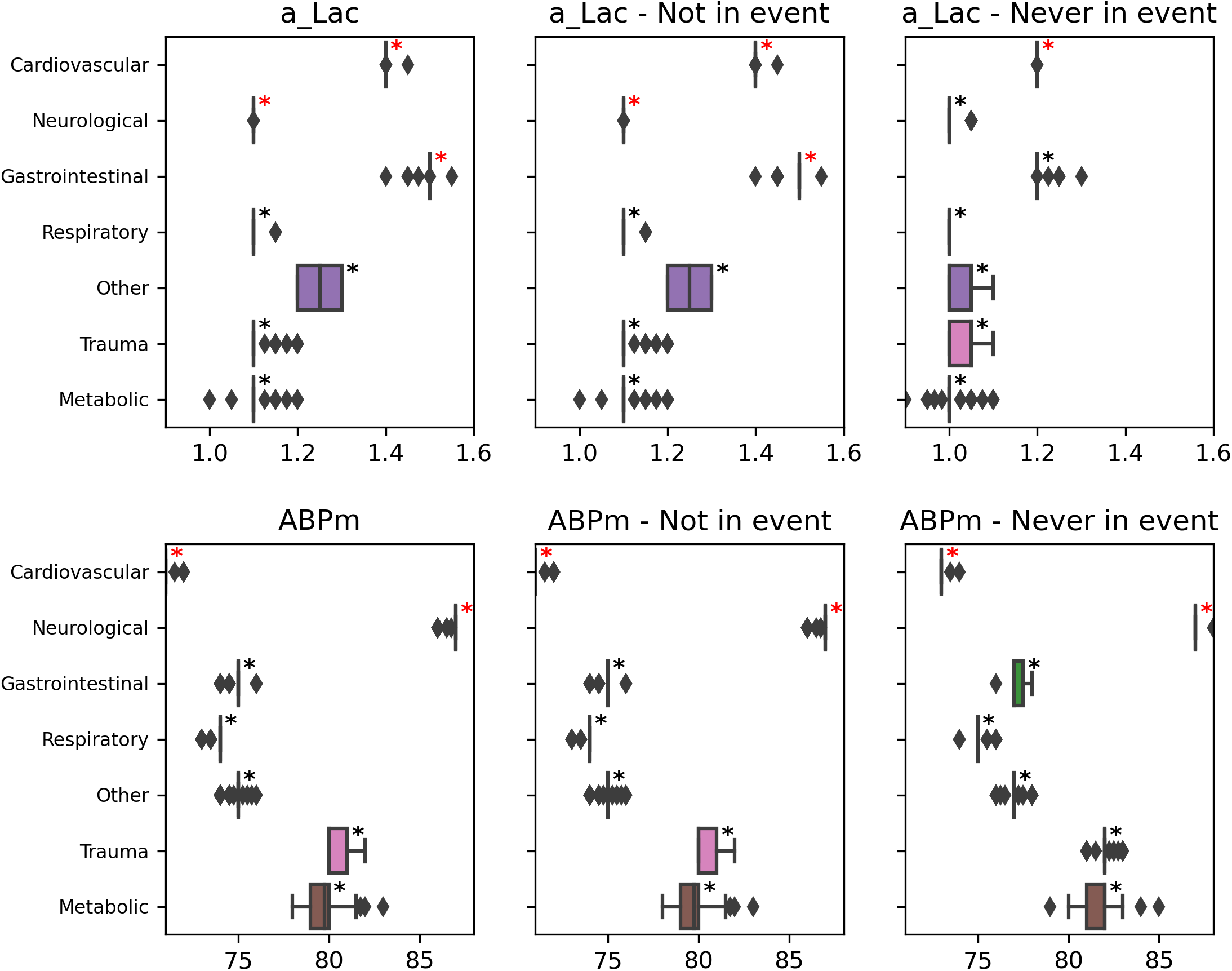

**Table 4.2.3.a.**

| Medical Variable | Category | Cohort vs. rest | P-value | Delta |
| --- | --- | --- | --- | --- |
| a_Lac | Cardiovascular | greater | 5.77e-40 | 0.25 |
| a_Lac | Neurological | less | 8.43e-41 | 0.25 |
| a_Lac | Gastrointestinal | greater | 2.23e-43 | 0.25 |
| a_Lac | Respiratory | less | 4.14e-44 | 0.2 |
| a_Lac | Other | less | 4.51e-21 | 0.05 |
| a_Lac | Trauma | less | 1.37e-41 | 0.2 |
| a_Lac | Metabolic | less | 4.28e-42 | 0.2 |
| a_Lac - Not in event | Cardiovascular | greater | 5.77e-40 | 0.25 |
| a_Lac - Not in event | Neurological | less | 9.91e-41 | 0.25 |
| a_Lac - Not in event | Gastrointestinal | greater | 3.22e-43 | 0.25 |
| a_Lac - Not in event | Respiratory | less | 4.14e-44 | 0.2 |
| a_Lac - Not in event | Other | less | 4.51e-21 | 0.05 |
| a_Lac - Not in event | Trauma | less | 1.37e-41 | 0.2 |
| a_Lac - Not in event | Metabolic | less | 4.28e-42 | 0.2 |
| a_Lac - Never in event | Cardiovascular | greater | 1.90e-42 | 0.15 |
| a_Lac - Never in event | Neurological | less | 3.20e-41 | 0.1 |
| a_Lac - Never in event | Gastrointestinal | greater | 1.32e-43 | 0.1 |
| a_Lac - Never in event | Respiratory | less | 1.76e-45 | 0.1 |
| a_Lac - Never in event | Other | less | 8.31e-37 | 0.1 |
| a_Lac - Never in event | Trauma | less | 2.33e-35 | 0.05 |
| a_Lac - Never in event | Metabolic | less | 7.89e-39 | 0.1 |
| ABPm | Cardiovascular | less | 6.14e-44 | 10.0 |
| ABPm | Neurological | greater | 1.76e-42 | 14.0 |
| ABPm | Gastrointestinal | less | 2.10e-37 | 2.0 |
| ABPm | Respiratory | less | 2.95e-41 | 3.0 |
| ABPm | Other | less | 5.26e-36 | 1.0 |
| ABPm | Trauma | greater | 4.33e-41 | 4.0 |
| ABPm | Metabolic | greater | 6.85e-39 | 3.75 |
| ABPm - Not in event | Cardiovascular | less | 9.05e-44 | 10.0 |
| ABPm - Not in event | Neurological | greater | 1.76e-42 | 14.0 |
| ABPm - Not in event | Gastrointestinal | less | 2.10e-37 | 2.0 |
| ABPm - Not in event | Respiratory | less | 2.95e-41 | 3.0 |
| ABPm - Not in event | Other | less | 5.26e-36 | 1.0 |
| ABPm - Not in event | Trauma | greater | 4.33e-41 | 4.0 |
| ABPm - Not in event | Metabolic | greater | 6.85e-39 | 3.75 |
| ABPm - Never in event | Cardiovascular | less | 4.65e-45 | 10.0 |
| ABPm - Never in event | Neurological | greater | 7.39e-45 | 12.0 |
| ABPm - Never in event | Gastrointestinal | less | 2.35e-41 | 2.0 |
| ABPm - Never in event | Respiratory | less | 2.8e-38 | 4.0 |
| ABPm - Never in event | Other | less | 6.96e-42 | 2.0 |
| ABPm - Never in event | Trauma | greater | 2.6e-41 | 3.0 |
| ABPm - Never in event | Metabolic | greater | 3.47e-39 | 3.0 |

##### 4.2.4. … surgical_status

**Figure 4.2.4.a.**
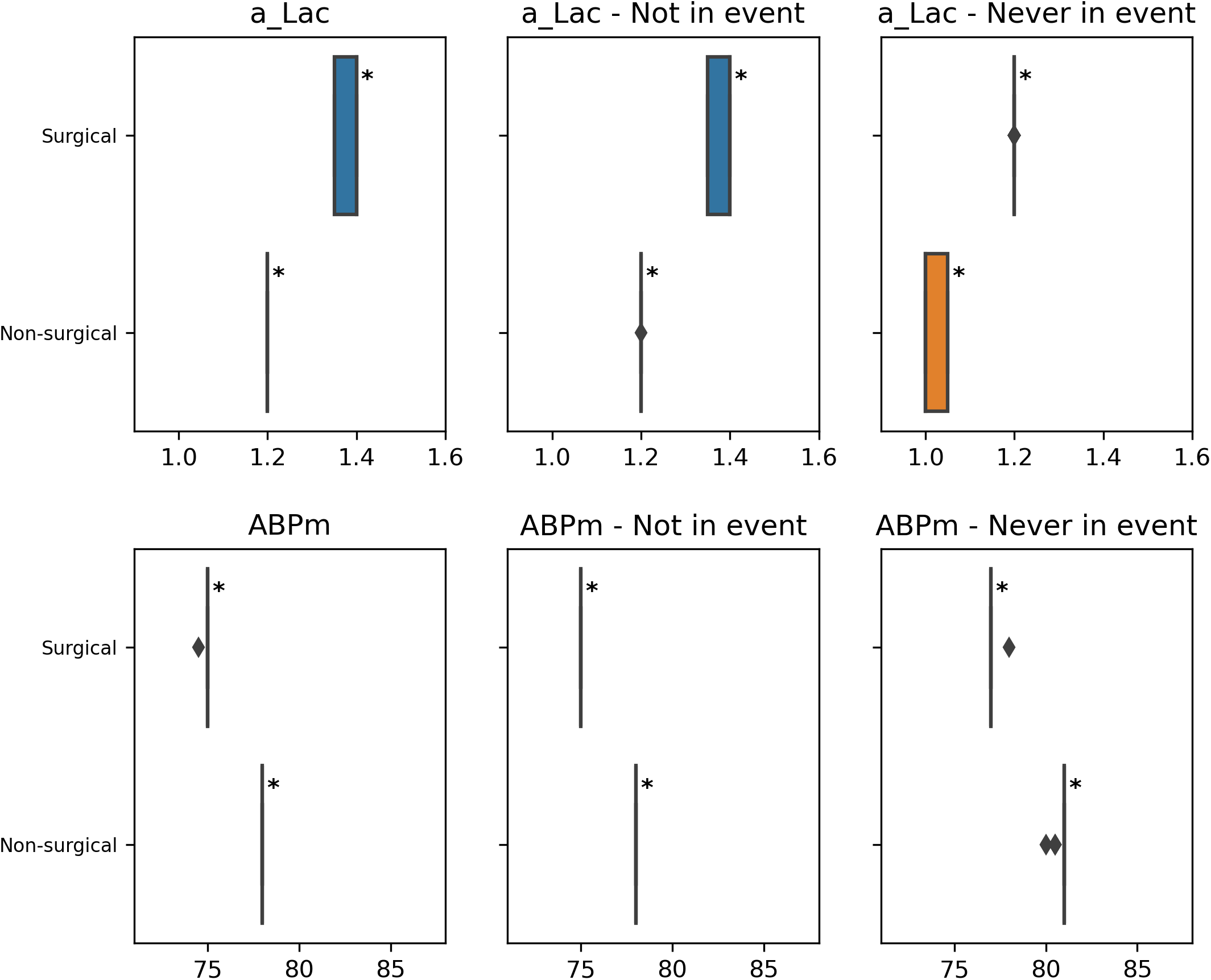

**Table 4.2.4.a.**
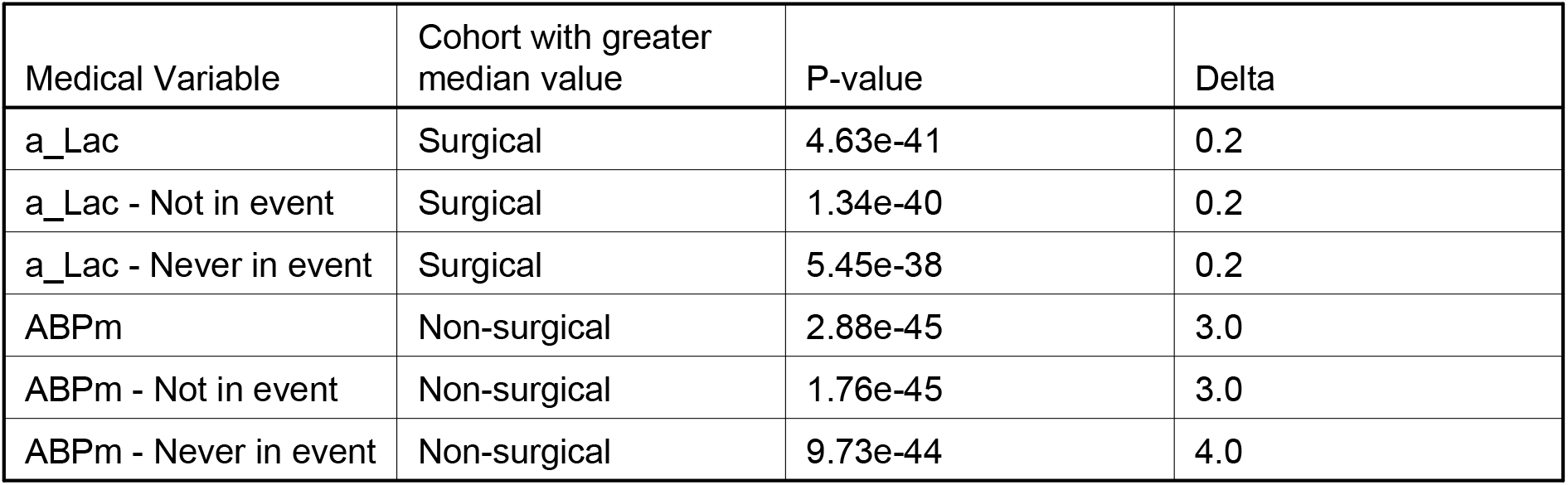

| Medical Variable | Cohort with greater median value | P-value | Delta |
| --- | --- | --- | --- |
| a_Lac | Surgical | 4.63e-41 | 0.2 |
| a_Lac - Not in event | Surgical | 1.34e-40 | 0.2 |
| a_Lac - Never in event | Surgical | 5.45e-38 | 0.2 |
| ABPm | Non-surgical | 2.88e-45 | 3.0 |
| ABPm - Not in event | Non-surgical | 1.76e-45 | 3.0 |
| ABPm - Never in event | Non-surgical | 9.73e-44 | 4.0 |

### 5. Feature importance Analysis

***Goal: Comparing the top 15 most important features across cohorts***

#### 5.1. Aggregated views

##### 5.1.1 Similarity of feature ranking per groupig

The following table displays the RBO (similarity measure) between the feature ranking for a patients’ cohort and the general feature ranking. We consider the feature ranking for a specific cohort to be significantly different when its RBO is smaller than 0.627 (colored in red in the table).

**Table 5.1.1.a.**

| Grouping | Category | RBO |
| --- | --- | --- |
| sex | F | 0.635 |
| sex | M | 0.627 |
| age_group | <50 | 0.574 |
| age_group | 50-65 | 0.618 |
| age_group | 65-75 | 0.617 |
| age_group | 75-85 | 0.633 |
| age_group | >85 | 0.627 |
| APACHE_group | Cardiovascular | 0.611 |
| APACHE_group | Neurological | 0.618 |
| APACHE_group | Gastrointestinal | 0.622 |
| APACHE_group | Respiratory | 0.615 |
| APACHE_group | Other | 0.619 |
| APACHE_group | Trauma | 0.607 |
| APACHE_group | Metabolic | 0.597 |
| surgical_status | Surgical | 0.627 |
| surgical_status | Non-surgical | 0.635 |

#### 5.2. Grouping by

We will now display for each grouping, the top 15 most important features. When the feature’s rank changes compared to the general ranking, we put the rank difference in parentheses.

We color in red the features that aren’t in the general top 15 features and in blue the ones that change place within the top 15, when their delta of inverse rank is significantly large.

##### 5.2.1. … sex

**Table 5.2.1.a.**

| Top 15 | Top 15 F | Top 15 M |
| --- | --- | --- |
| a_Lac | a_Lac | a_Lac |
| datetime | datetime | datetime |
| ABPm | ABPm | ABPm |
| ABPs | ABPs | ABPs |
| HR | HR | HR |
| ABPd | ABPd | Spitzendruck (↑ 1) |
| Spitzendruck | Spitzendruck | ABPd (↓ 1) |
| RASS | RASS | RASS |
| age | age | age |
| a-BE | a-BE | a-BE |
| creatinine | creatinine | creatinine |
| norepinephrine | norepinephrine | norepinephrine |
| ETCO2 | ETCO2 | ETCO2 |
| glucose | glucose | glucose |
| INR | INR | INR |

##### 5.2.2. … age_group

**Table 5.2.2.a.**
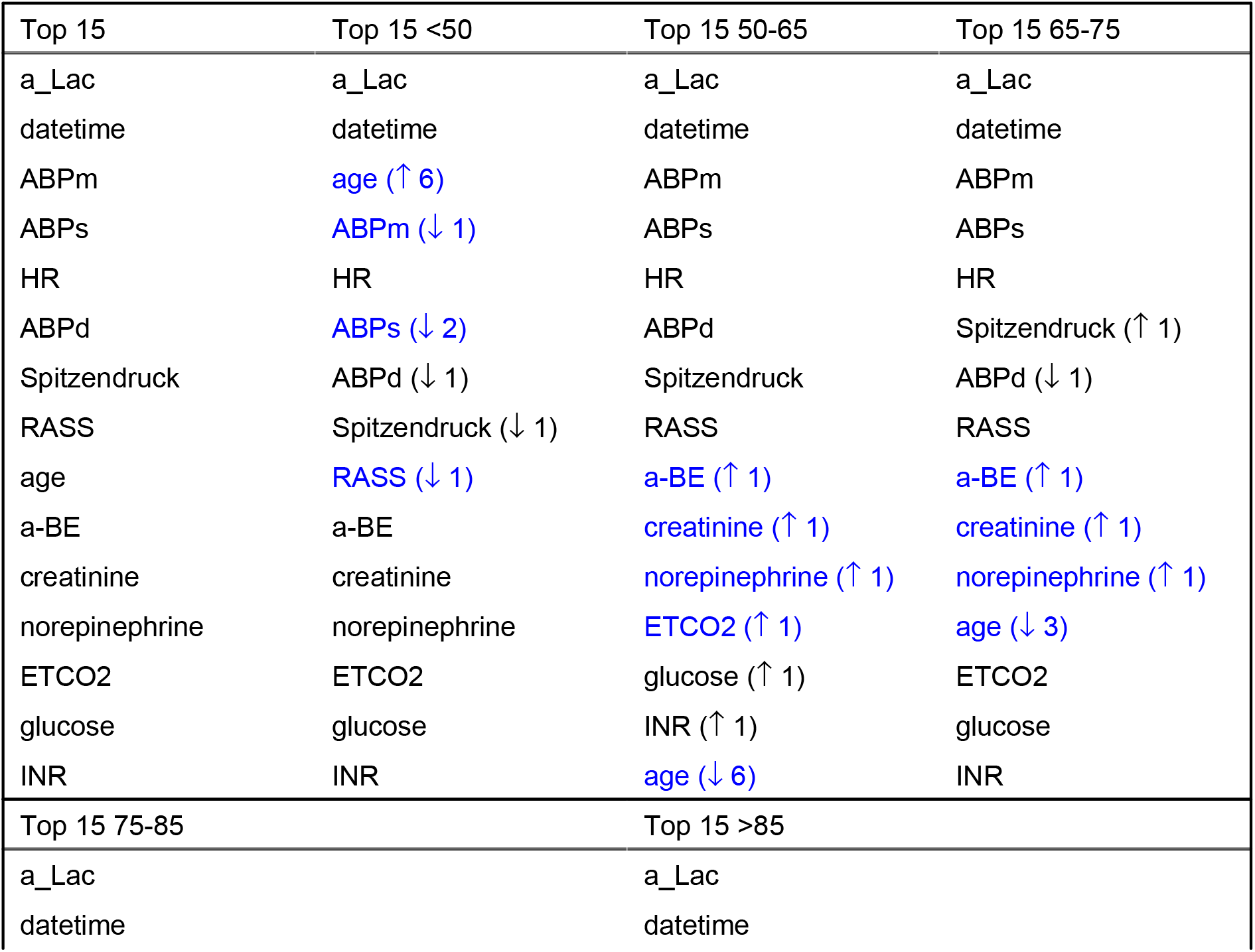
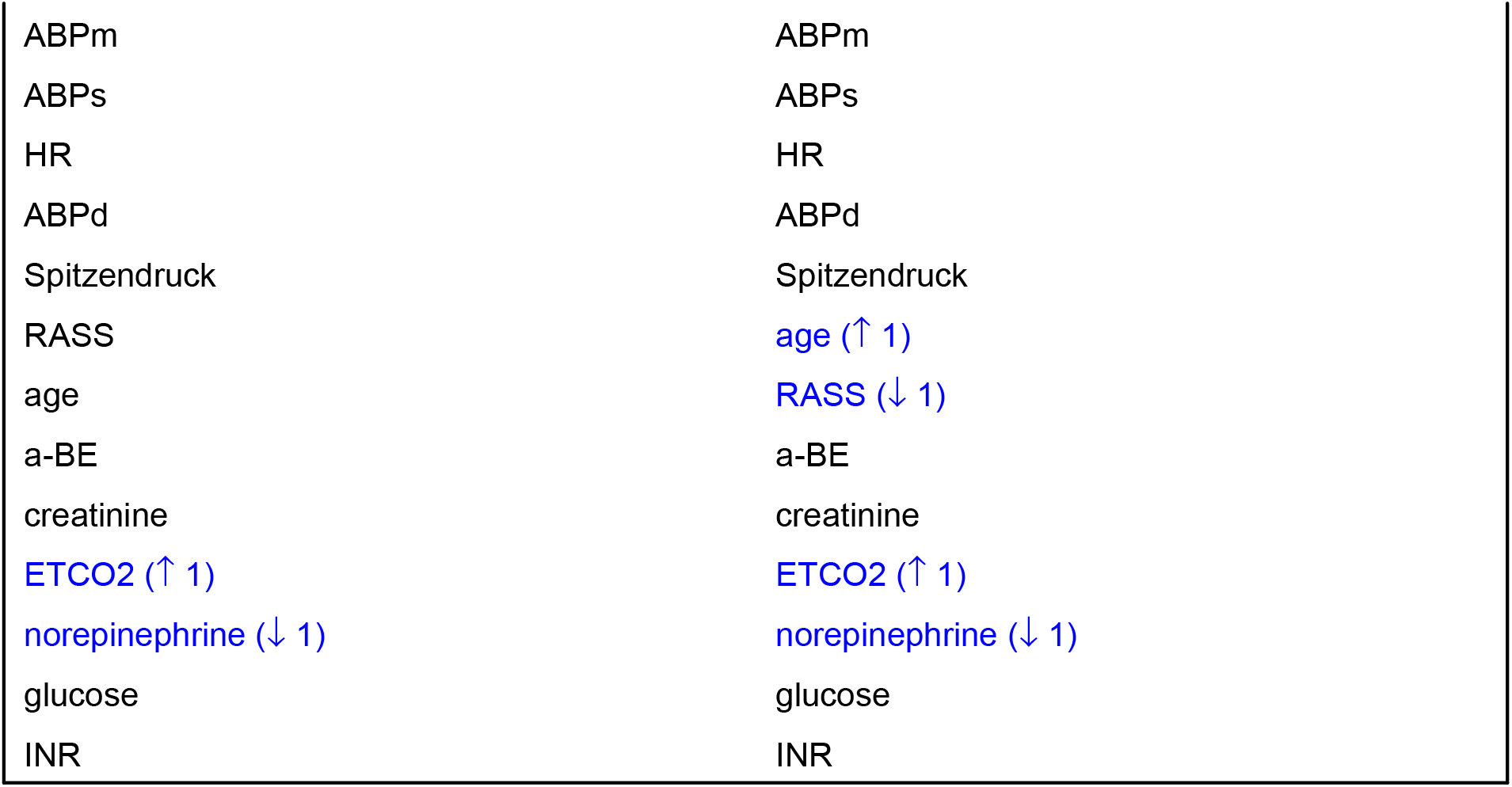

| Top 15 | Top 15 <50 | Top 15 50-65 | Top 15 65-75 |
| --- | --- | --- | --- |
| a_Lac | a_Lac | a_Lac | a_Lac |
| datetime | datetime | datetime | datetime |
| ABPm | age (↑ 6) | ABPm | ABPm |
| ABPs | ABPm (↓ 1) | ABPs | ABPs |
| HR | HR | HR | HR |
| ABPd | ABPs (↓ 2) | ABPd | Spitzendruck (↑ 1) |
| Spitzendruck | ABPd (↓ 1) | Spitzendruck | ABPd (↓ 1) |
| RASS | Spitzendruck (↓ 1) | RASS | RASS |
| age | RASS (↓ 1) | a-BE (↑ 1) | a-BE (↑ 1) |
| a-BE | a-BE | creatinine (↑ 1) | creatinine (↑ 1) |
| creatinine | creatinine | norepinephrine (↑ 1) | norepinephrine (↑ 1) |
| norepinephrine | norepinephrine | ETCO2 (↑ 1) | age (↓ 3) |
| ETCO2 | ETCO2 | glucose (↑ 1) | ETCO2 |
| glucose | glucose | INR (↑ 1) | glucose |
| INR | INR | age (↓ 6) | INR |
| Top 15 75-85 |  | Top 15 >85 |  |
| a_Lac |  | a_Lac |  |
| datetime |  | datetime |  |

|  |  |
| --- | --- |
| ABPm | ABPm |
| ABPs | ABPs |
| HR | HR |
| ABPd | ABPd |
| Spitzendruck | Spitzendruck |
| RASS | age (↑ 1) |
| age | RASS (↓ 1) |
| a-BE | a-BE |
| creatinine | creatinine |
| ETCO2 (↑ 1) | ETCO2 (↑ 1) |
| norepinephrine (↓ 1) | norepinephrine (↓ 1) |
| glucose | glucose |
| INR | INR |

##### 5.2.3. … APACHE_group

**Table 5.2.3.a.**

| Top 15 | Top 15 Cardiovascular | Top 15 Neurological | Top 15 Gastrointestinal |
| --- | --- | --- | --- |
| a_Lac | a_Lac | a_Lac | a_Lac |
| datetime | datetime | datetime | datetime |
| ABPm | ABPm | ABPm | ABPm |
| ABPs | ABPs | ABPs | ABPs |
| HR | Spitzendruck (↑ 2) | HR | HR |
| ABPd | HR (↓ 1) | ABPd | ABPd |
| Spitzendruck | ABPd (↓ 1) | RASS (↑ 1) | Spitzendruck |
| RASS | RASS | Spitzendruck (↓ 1) | a-BE (↑ 2) |
| age | age | age | age |
| a-BE | a-BE | norepinephrine (↑ 2) | RASS (↓ 2) |
| creatinine | creatinine | creatinine | creatinine |
| norepinephrine | ETCO2 (↑ 1) | a-BE (↓ 2) | norepinephrine |
| ETCO2 | INR (↑ 2) | glucose (↑ 1) | INR (↑ 2) |
| glucose | norepinephrine (↓ 2) | ETCO2 (↓ 1) | ETCO2 (↓ 1) |
| INR | glucose (↓ 1) | NIBPm (↑ 1) | glucose (↓ 1) |
| Top 15 Respiratory | Top 15 Other | Top 15 Trauma | Top 15 Metabolic |
| a_Lac | a_Lac | a_Lac | a_Lac |
| datetime | datetime | datetime | datetime |
| ABPm | ABPm | ABPm | ABPm |
| HR (↑ 1) | ABPs | HR (↑ 1) | ABPs |
| ABPs (↓ 1) | HR | ABPs (↓ 1) | HR |
| ABPd | ABPd | Spitzendruck (↑ 1) | ABPd |
| Spitzendruck | Spitzendruck | ABPd (↓ 1) | a-BE (↑ 3) |
| RASS | a-BE (↑ 2) | age (↑ 1) | age (↑ 1) |
| a-BE (↑ 1) | age | RASS (↓ 1) | Spitzendruck (↓ 2) |
| age (↓ 1) | RASS (↓ 2) | a-BE | glucose (↑ 4) |
| creatinine | norepinephrine (↑ 1) | creatinine | RASS (↓ 3) |
| ETCO2 (↑ 1) | creatinine (↓ 1) | norepinephrine | creatinine (↓ 1) |
| norepinephrine (↓ 1) | INR (↑ 2) | ETCO2 | NIBPm (↑ 3) |
| glucose | ETCO2 (↓ 1) | glucose | norepinephrine (↓ 2) |
| INR | glucose (↓ 1) | GCS Motorik (↑ 2) | ETCO2 (↓ 2) |

##### 5.2.4. … surgical_status

**Table 5.2.4.a.**

| Top 15 | Top 15 Surgical | Top 15 Non-surgical |
| --- | --- | --- |
| a_Lac | a_Lac | a_Lac |
| datetime | datetime | datetime |
| ABPm | ABPm | ABPm |
| ABPs | ABPs | ABPs |
| HR | HR | HR |
| ABPd | Spitzendruck (↑ 1) | ABPd |
| Spitzendruck | ABPd (↓ 1) | Spitzendruck |
| RASS | RASS | RASS |
| age | age | age |
| a-BE | a-BE | a-BE |
| creatinine | creatinine | creatinine |
| norepinephrine | norepinephrine | norepinephrine |
| ETCO2 | ETCO2 | ETCO2 |
| glucose | glucose | glucose |
| INR | INR | INR |

### 6. Missingness Analysis

***Goal: Comparing the intensity of measurements across cohorts of patients and its impact of performance***

Binary metrics computed with a threshold on score of 0.445.

#### 6.1. Aggregated views

##### 6.1.1. a_Lac

###### Groupings that are statistically dependent on the intensity of measurements

**Table 6.1.1.a.**

| Group name | Category with the biggest rate of no_msrt | Category with the biggest rate of insufficient |
| --- | --- | --- |
| sex | F | M |
| age_group | <50 | 75-85 |
| APACHE_group | Neurological | Cardiovascular |
| surgical_status | Surgical | Surgical |

###### Summary of the impact of missingness on performance

**missing_msrt**: 36.4% of metrics are worse than for with measurement time points, with the biggest delta 0.115 for metric Recall.

##### 6.1.2. Spitzendruck

###### Groupings that are statistically dependent on the intensity of measurements

**Table 6.1.2.a.**

| Group name | Category with the biggest rate of no_msrt | Category with the biggest rate of insufficient |
| --- | --- | --- |
| sex | F | M |
| age_group | <50 | 75-85 |
| APACHE_group | Metabolic | Cardiovascular |
| surgical_status | Non-surgical | Surgical |

###### Summary of the impact of missingness on performance

**no_msrt**: 45.5% of metrics are worse than for with measurement time points, with the biggest delta 0.175 for metric AUPRC.

**missing_msrt**: 45.5% of metrics are worse than for with measurement time points, with the biggest delta 0.155 for metric AUPRC.

For each grouping, we display a bar plot that shows the percentage of each intensity of measurement category within a cohort of patients. The dashed lines represent the percentage of each intensity of measurement category with respect to the entire patient population. We run the Chi-squared independence test (with significance level 0.001) to assess the depence between the intensity of measurement and the grouping.

In the impact on performance subsection, we present box plots that show the metrics’ distribution for each of the missingness categories. For each metric, we mark with a black star the missingness categories that are significantly worse compared to metrics computed on data points with present measurement.

We also propose tables presenting the results of the impact on performance statistical analysis, we display only metrics and missingness categories with a significant p-value (smaller than 0.001/number of comparisons) and whose delta is bigger than 0. We compare the metrics for missingness categories *missing_msrt* and *no_msrt* (when relevant) against the *with_msrt* category. P-values are obtained by running the Mann-Whitney U test with Bonferroni correction.

#### 6.2. Study of the variable a_Lac

##### 6.2.1. Intensity of measurement per grouping

###### Grouping by sex

**Figure 6.2.1.a.**
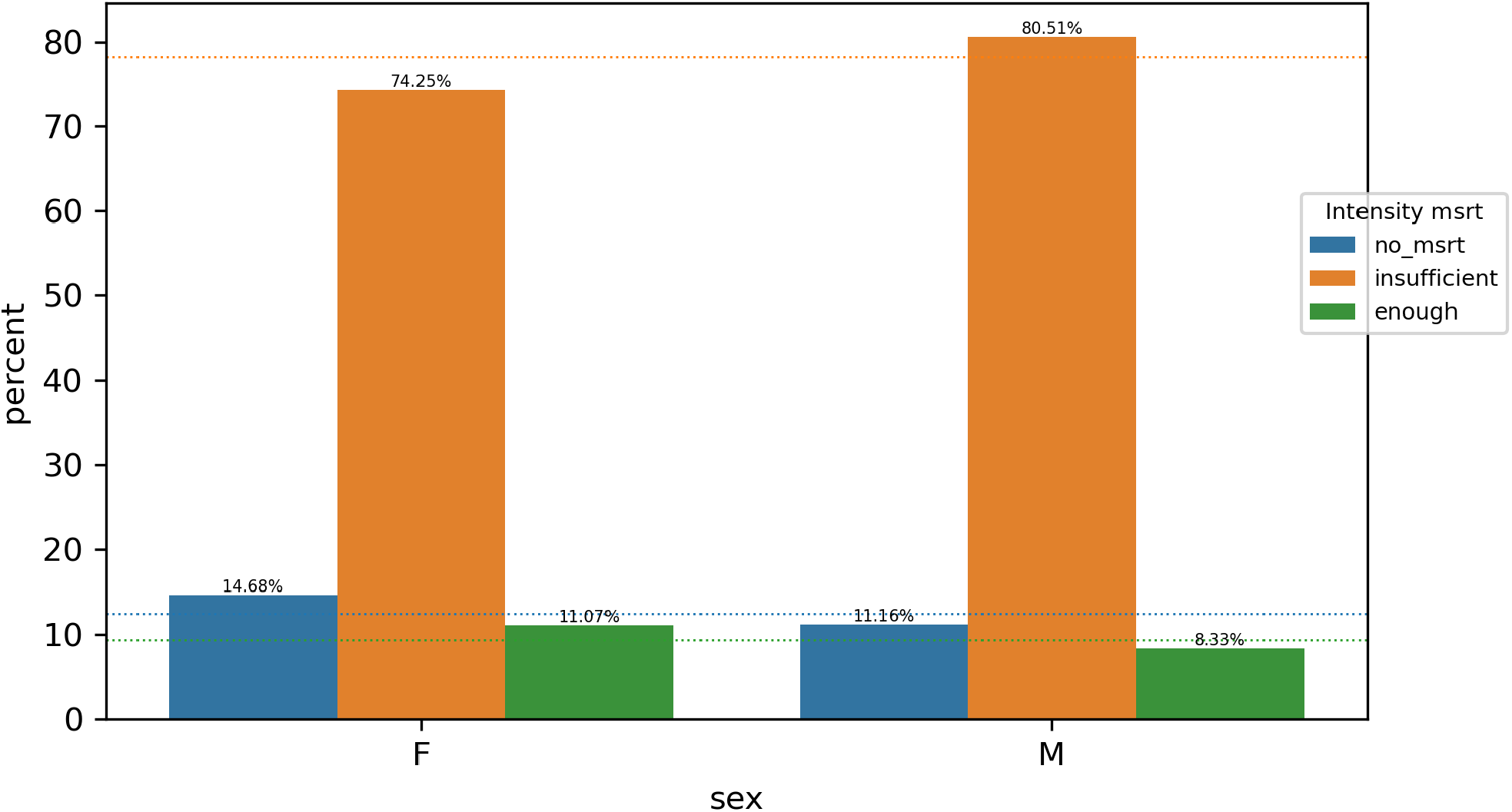

The intensity of measurements of a_Lac and sex attributes are <u>dependent</u>.

###### Grouping by age_group

**Figure 6.2.1.b.**
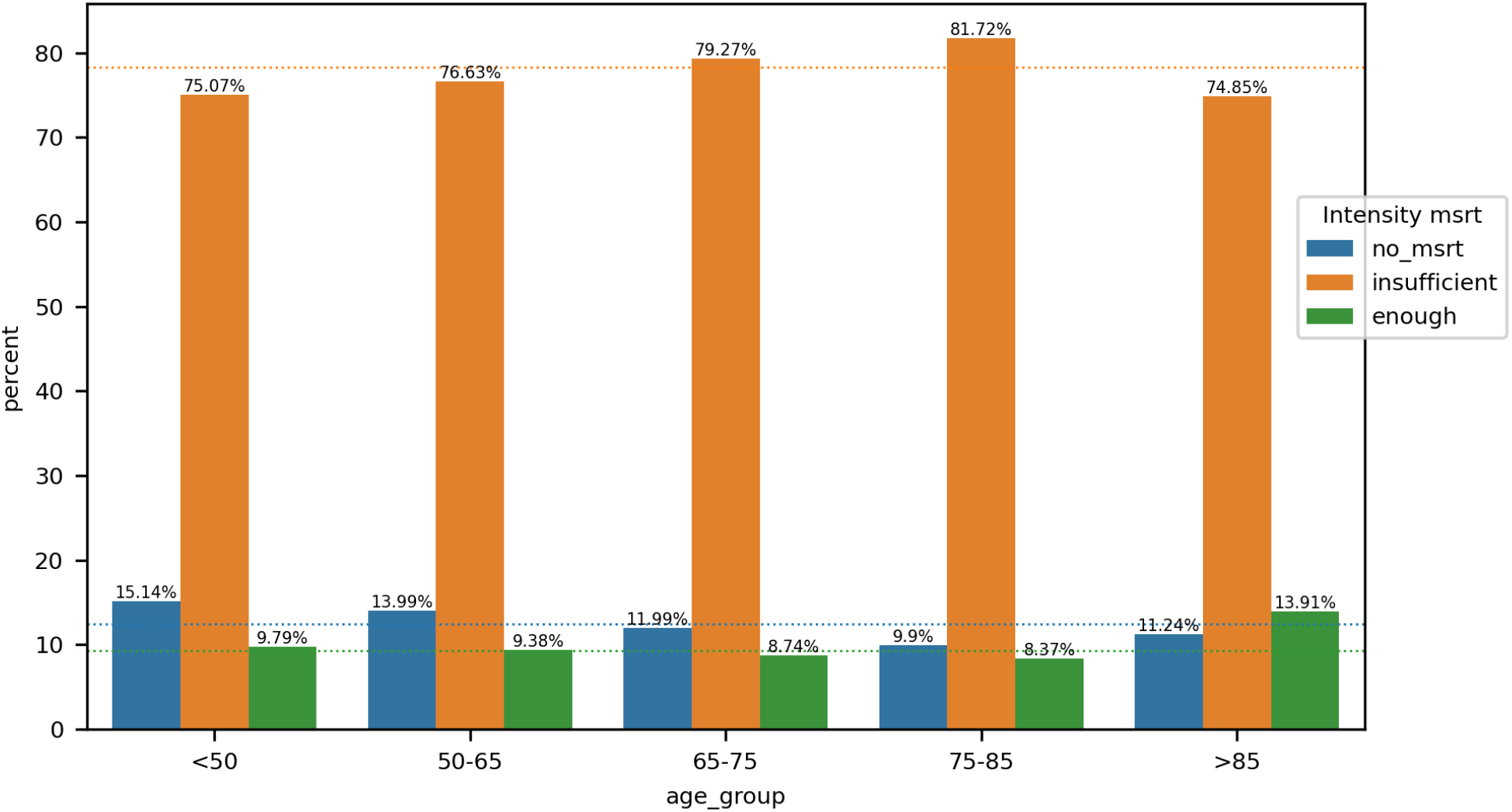

The intensity of measurements of a_Lac and age_group attributes are <u>dependent</u>.

###### Grouping by APACHE_group

**Figure 6.2.1.c.**
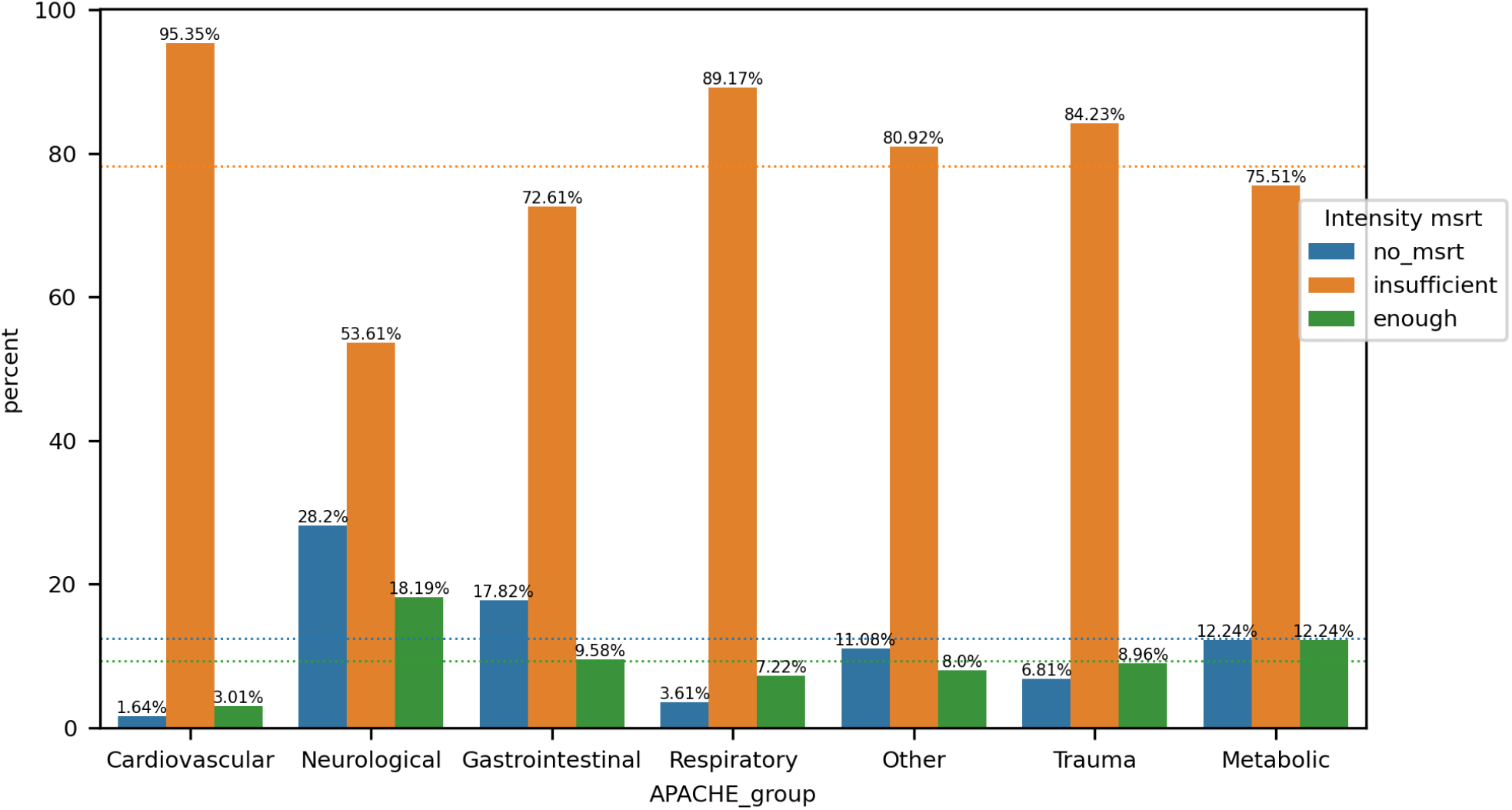

The intensity of measurements of a_Lac and APACHE_group attributes are <u>dependent</u>.

###### Grouping by surgical_status

**Figure 6.2.1.d.**
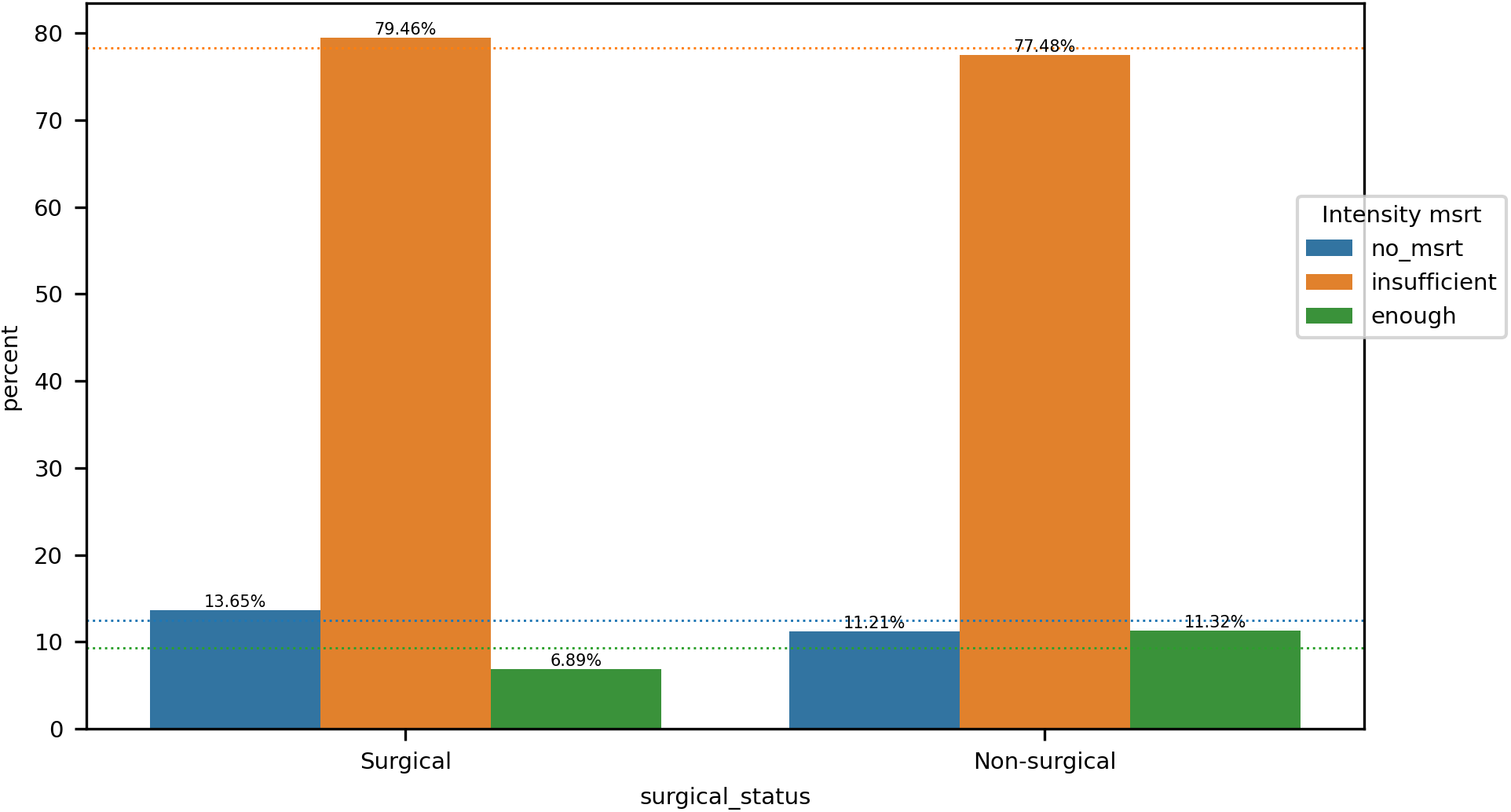

The intensity of measurements of a_Lac and surgical_status attributes are <u>dependent</u>.

##### 6.2.2. Impact on performance

**Figure 6.2.2.a.**
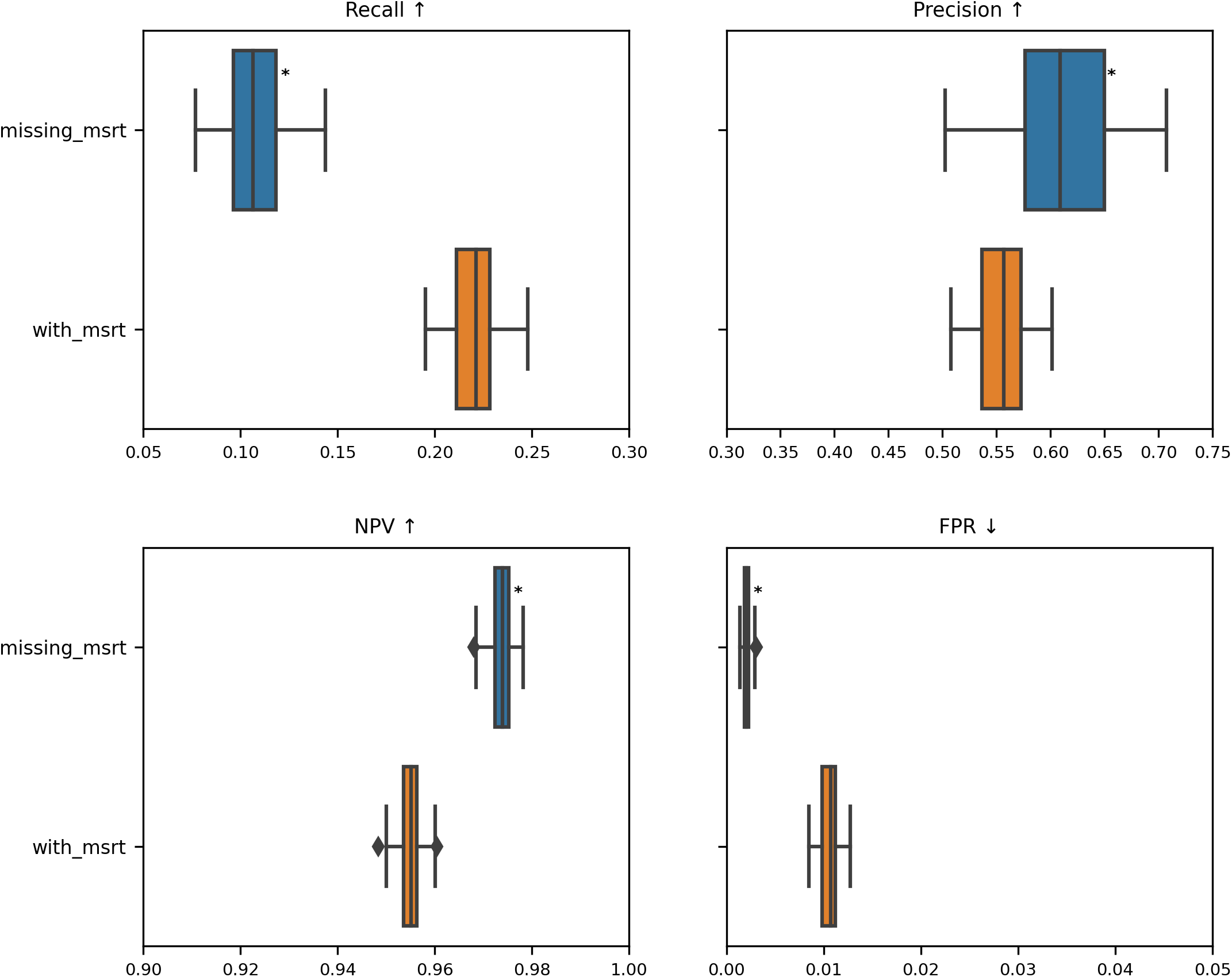
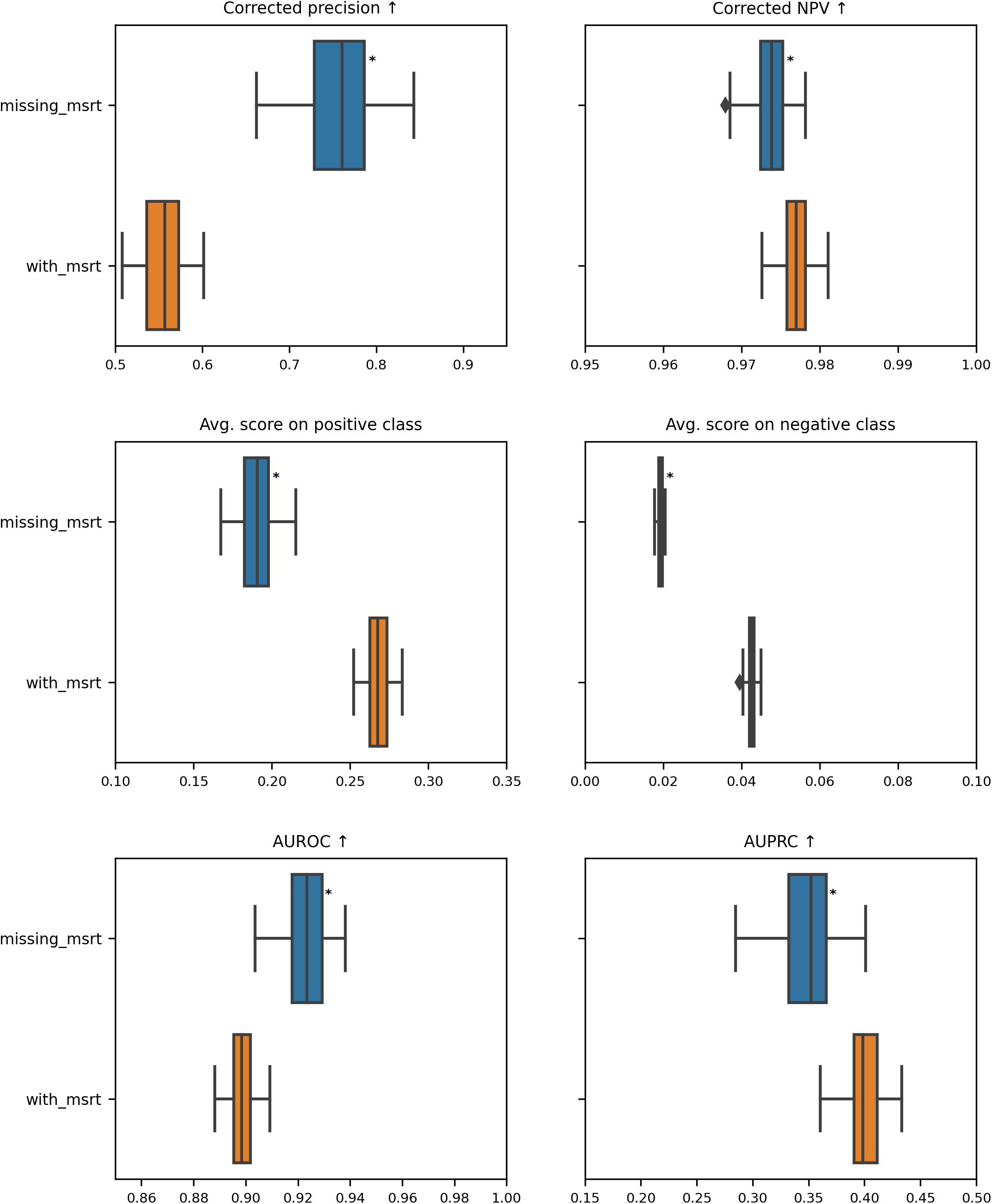
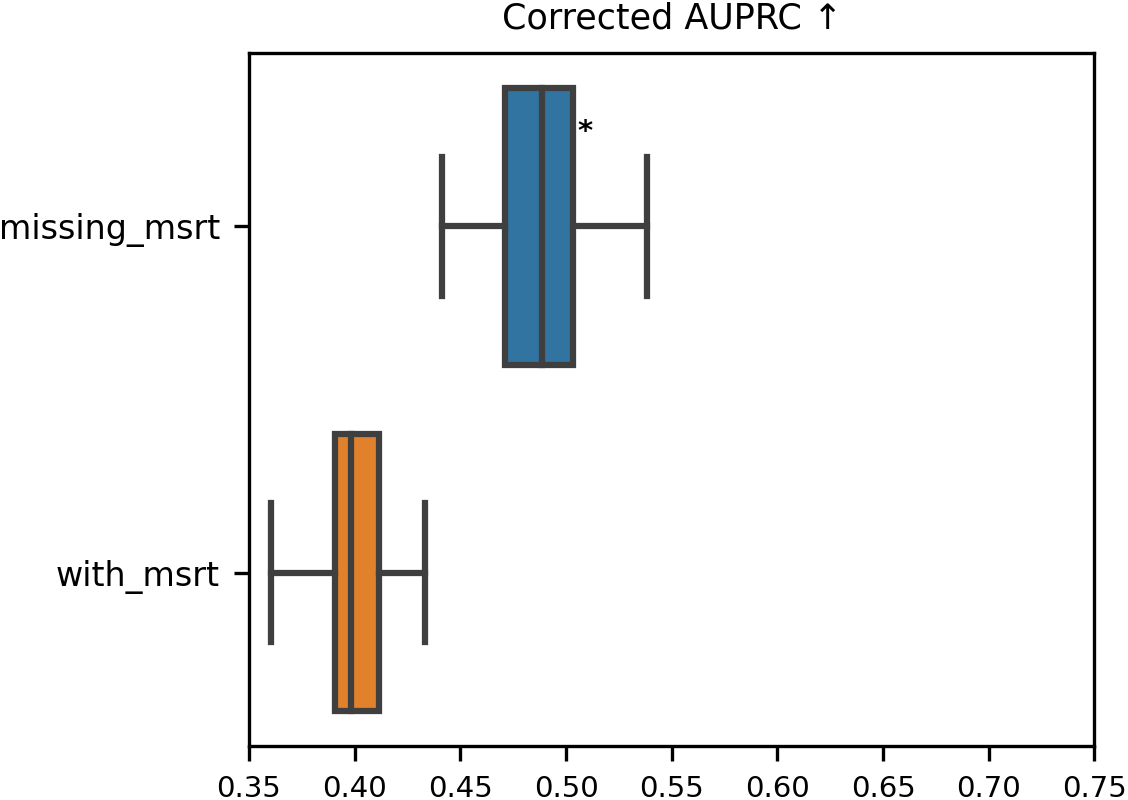

**Table 6.2.2.a.**
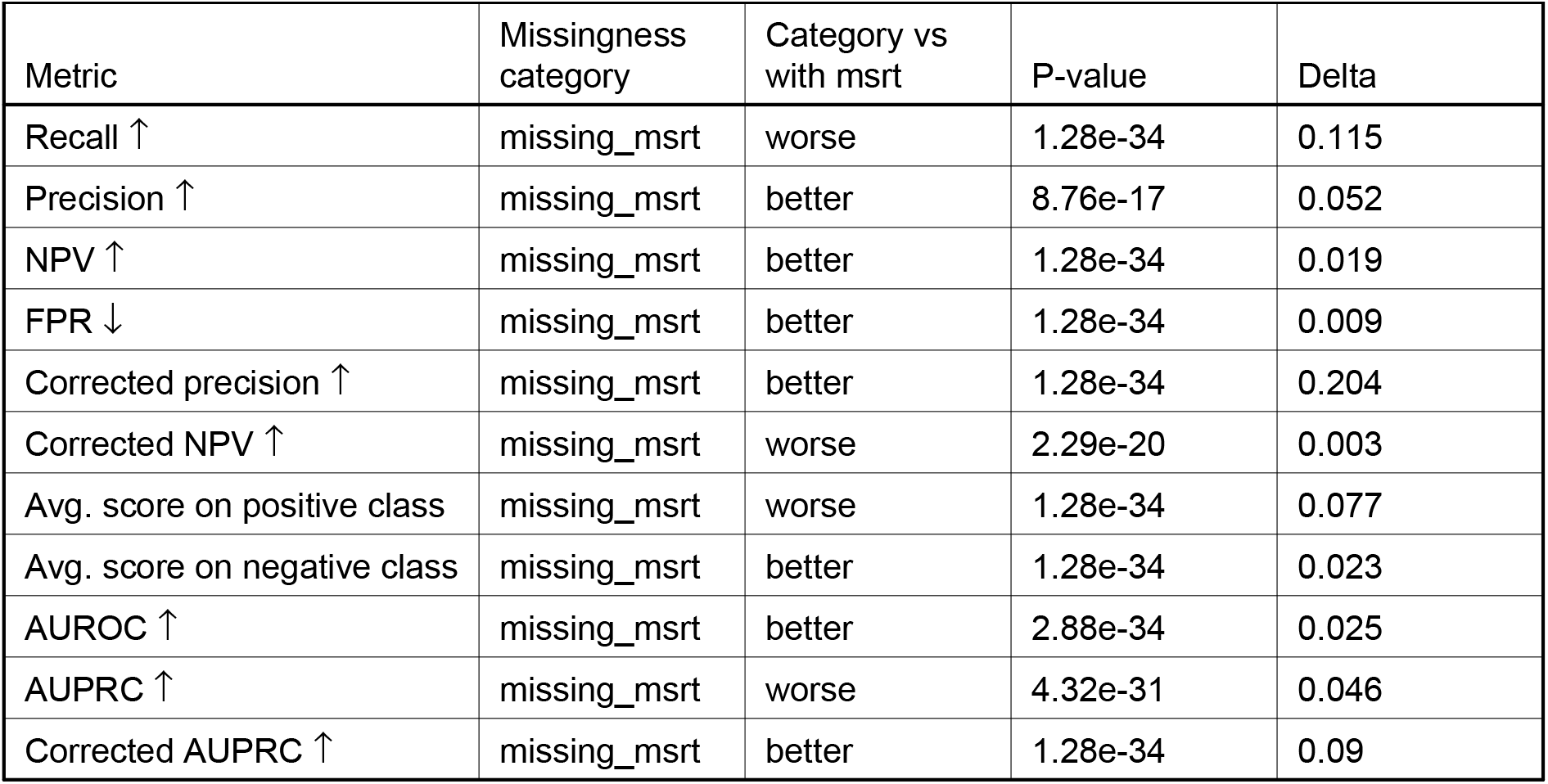

| Metric | Missingness category | Category vs with msrt | P-value | Delta |
| --- | --- | --- | --- | --- |
| Recall ↑ | missing_msrt | worse | 1.28e-34 | 0.115 |
| Precision ↑ | missing_msrt | better | 8.76e-17 | 0.052 |
| NPV ↑ | missing_msrt | better | 1.28e-34 | 0.019 |
| FPR ↓ | missing_msrt | better | 1.28e-34 | 0.009 |
| Corrected precision ↑ | missing_msrt | better | 1.28e-34 | 0.204 |
| Corrected NPV ↑ | missing_msrt | worse | 2.29e-20 | 0.003 |
| Avg. score on positive class | missing_msrt | worse | 1.28e-34 | 0.077 |
| Avg. score on negative class | missing_msrt | better | 1.28e-34 | 0.023 |
| AUROC ↑ | missing_msrt | better | 2.88e-34 | 0.025 |
| AUPRC ↑ | missing_msrt | worse | 4.32e-31 | 0.046 |
| Corrected AUPRC ↑ | missing_msrt | better | 1.28e-34 | 0.09 |

#### 6.3. Study of the variable Spitzendruck

##### 6.3.1. Intensity of measurement per grouping

###### Grouping by sex

**Figure 6.3.1.a.**
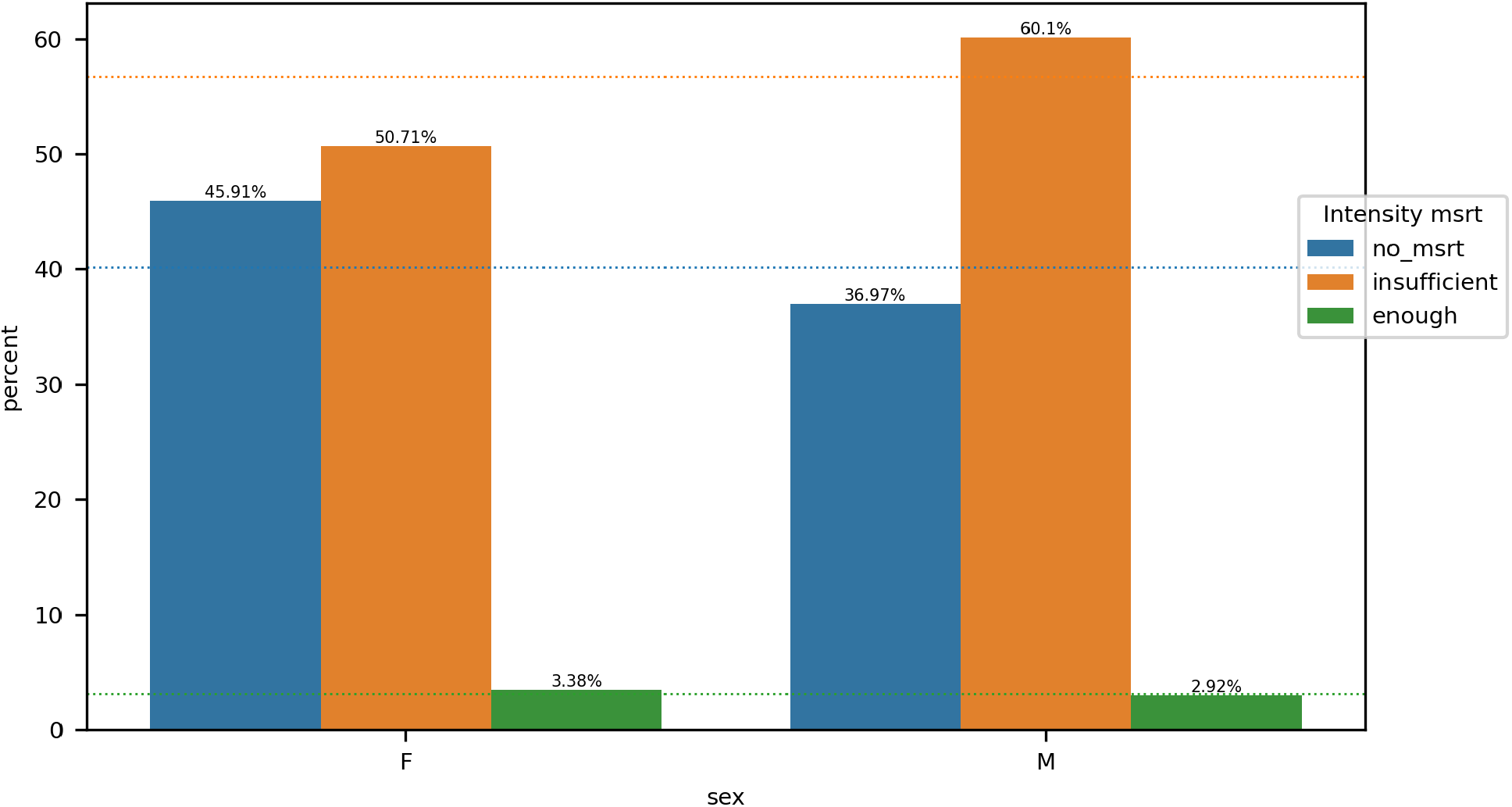

The intensity of measurements of Spitzendruck and sex attributes are <u>dependent</u>.

###### Grouping by age_group

**Figure 6.3.1.b.**
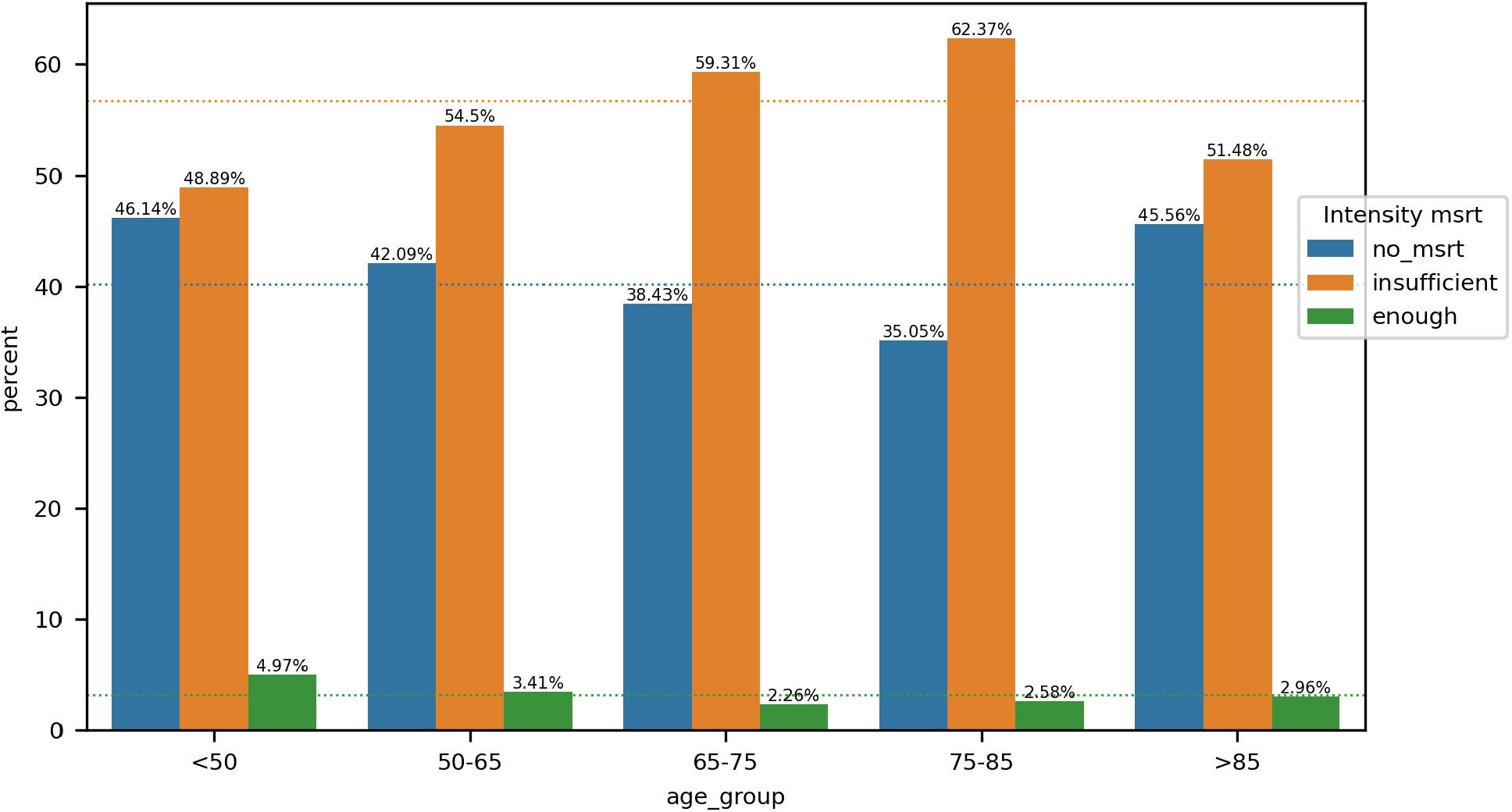

The intensity of measurements of Spitzendruck and age_group attributes are <u>dependent</u>.

###### Grouping by APACHE_group

**Figure 6.3.1.c.**
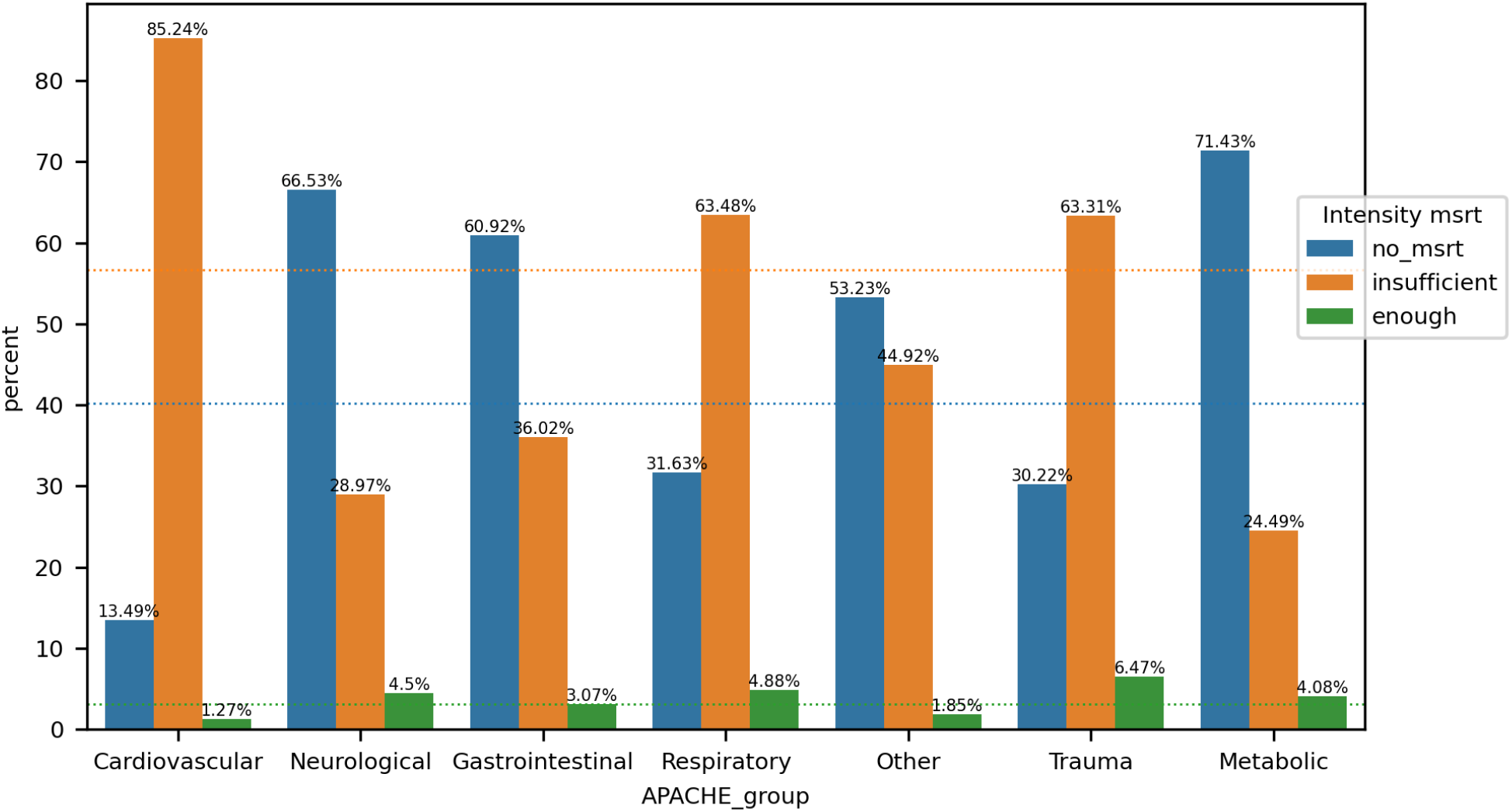

The intensity of measurements of Spitzendruck and APACHE_group attributes are <u>dependent</u>.

###### Grouping by surgical_status

**Figure 6.3.1.d.**
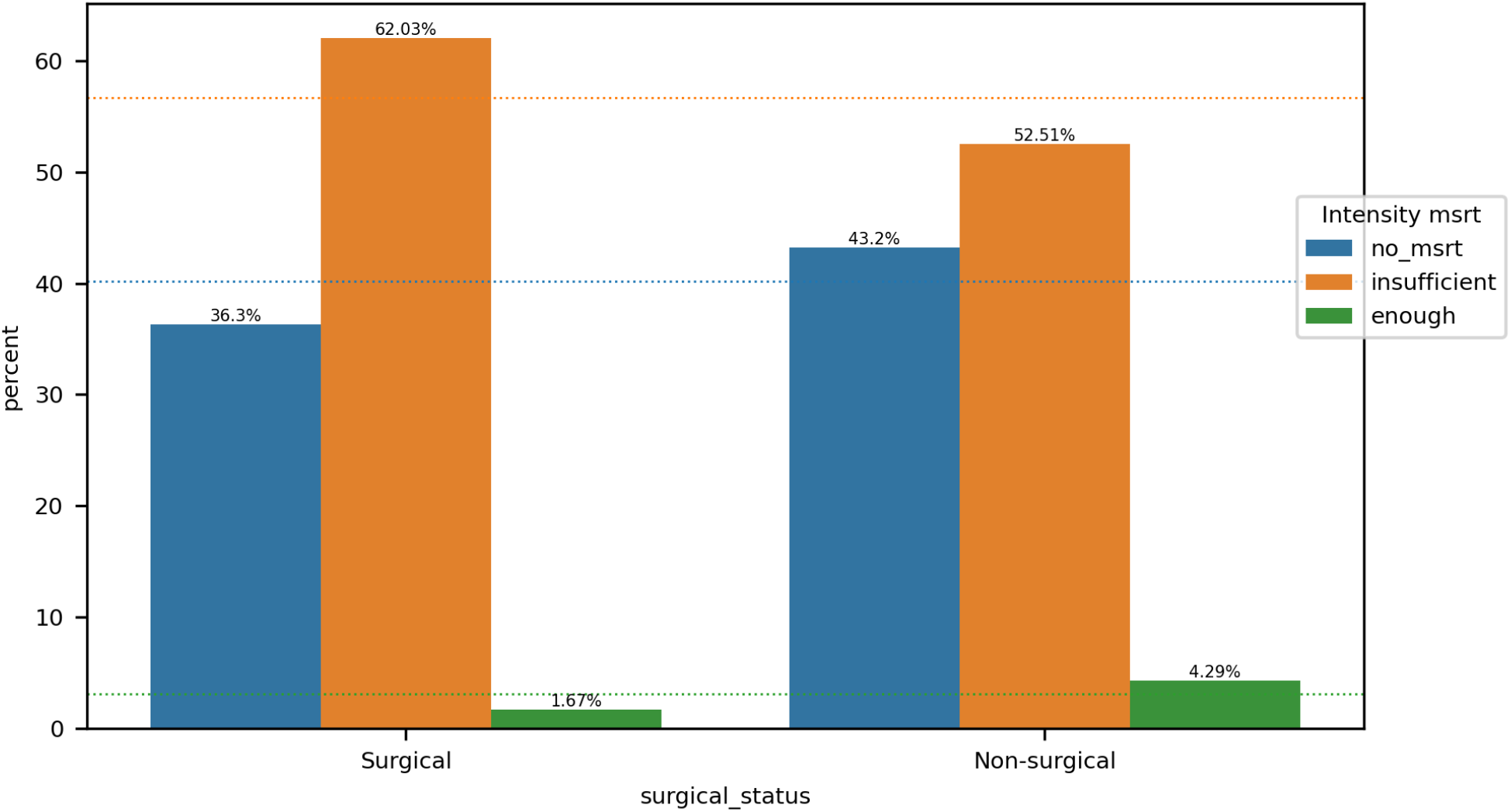

The intensity of measurements of Spitzendruck and surgical_status attributes are <u>dependent</u>.

##### 6.3.2. Impact on performance

**Figure 6.3.2.a.**
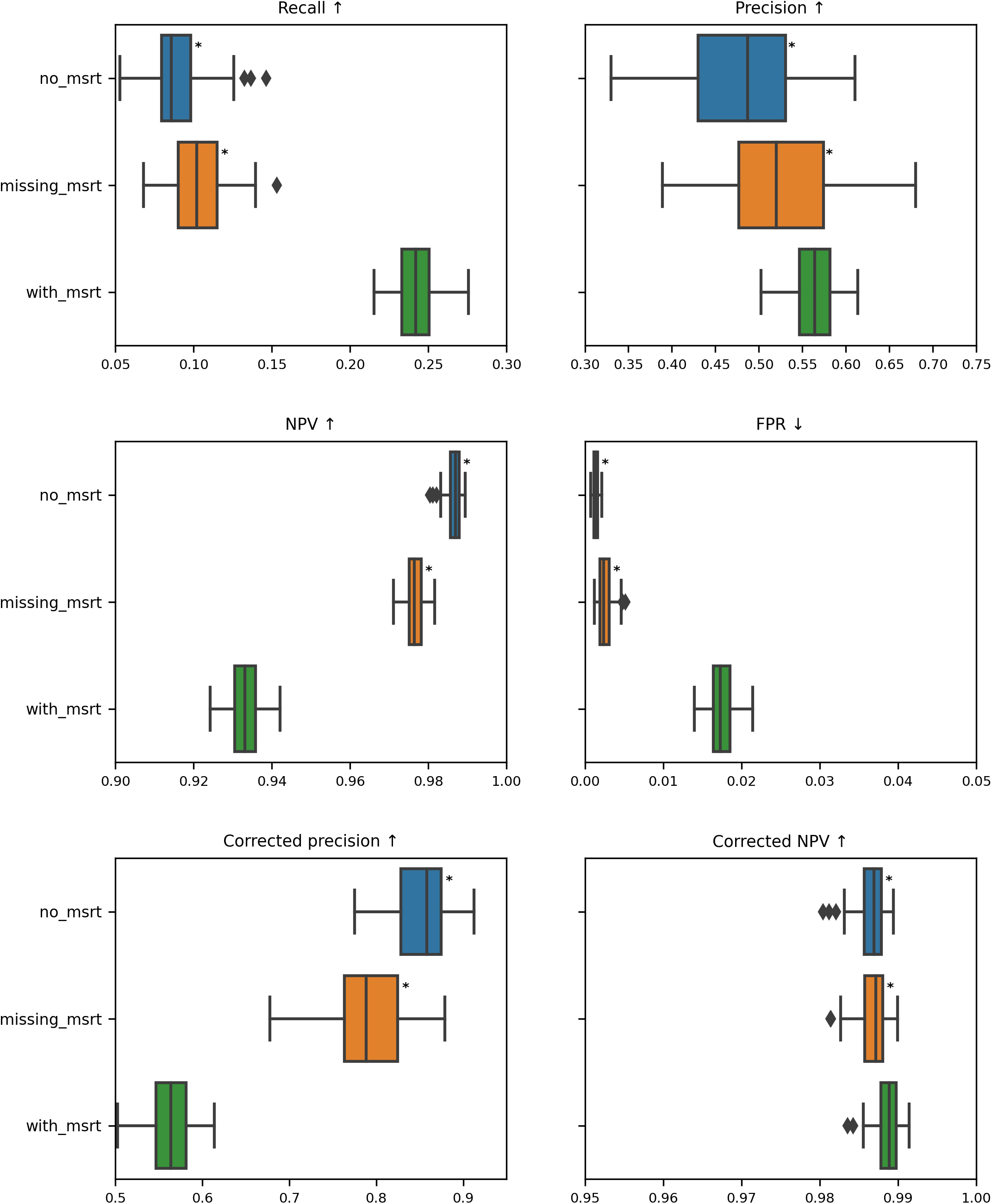
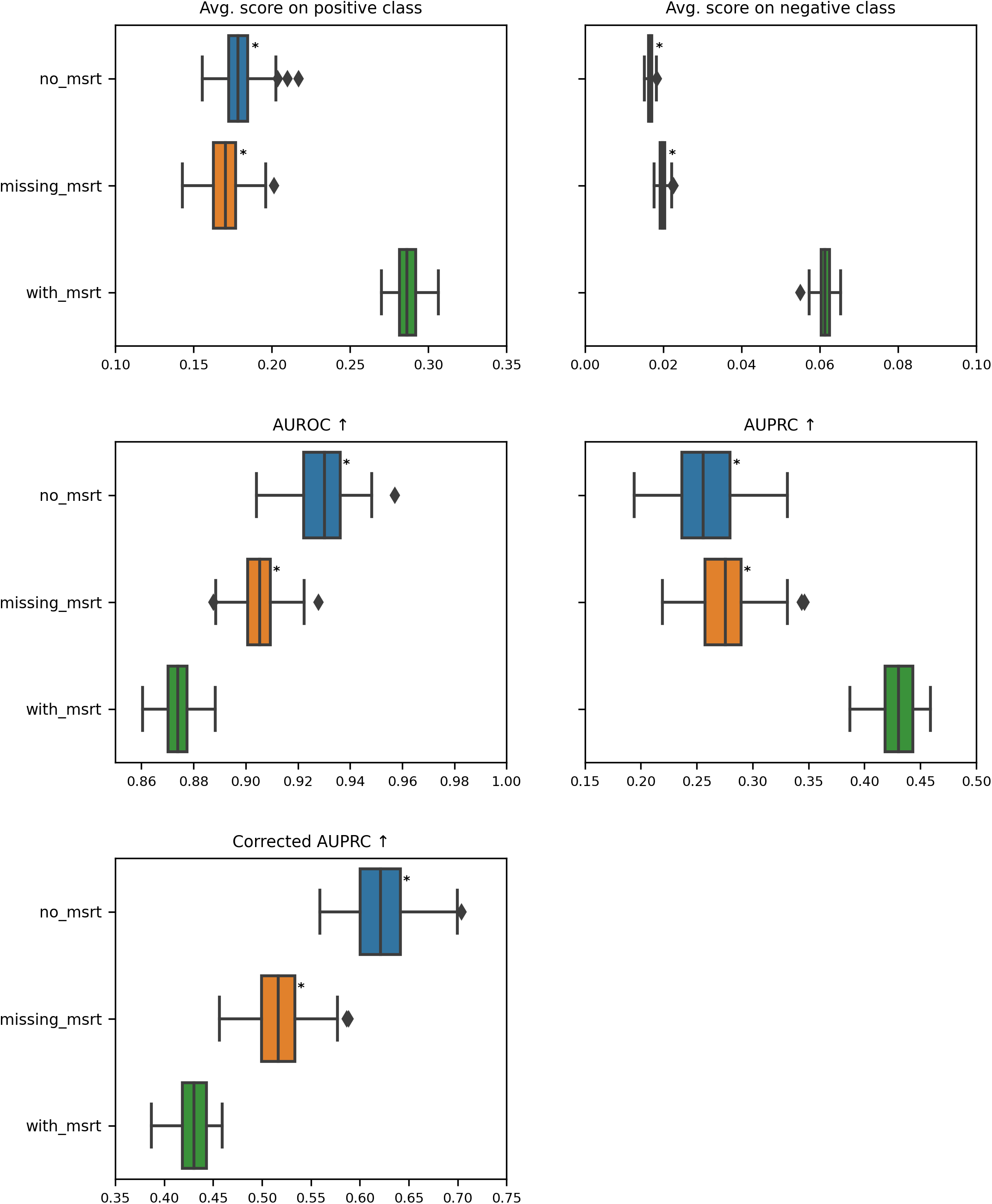

**Table 6.3.2.a.**

| Metric | Missingness category | Category vs with msrt | P-value | Delta |
| --- | --- | --- | --- | --- |
| Recall ↑ | no_msrt | worse | 1.28e-34 | 0.156 |
| Recall ↑ | missing_msrt | worse | 1.28e-34 | 0.14 |
| Precision ↑ | no_msrt | worse | 2.56e-19 | 0.077 |
| Precision ↑ | missing_msrt | worse | 6.21e-07 | 0.044 |
| NPV ↑ | no_msrt | better | 1.28e-34 | 0.054 |
| NPV ↑ | missing_msrt | better | 1.28e-34 | 0.043 |
| FPR ↓ | no_msrt | better | 1.28e-34 | 0.016 |
| FPR ↓ | missing_msrt | better | 1.28e-34 | 0.015 |
| Corrected precision ↑ | no_msrt | better | 1.28e-34 | 0.294 |
| Corrected precision ↑ | missing_msrt | better | 1.28e-34 | 0.225 |
| Corrected NPV ↑ | no_msrt | worse | 1.79e-15 | 0.002 |
| Corrected NPV ↑ | missing_msrt | worse | 1.16e-13 | 0.002 |
| Avg. score on positive class | no_msrt | worse | 1.28e-34 | 0.108 |
| Avg. score on positive class | missing_msrt | worse | 1.28e-34 | 0.116 |
| Avg. score on negative class | no_msrt | better | 1.28e-34 | 0.045 |
| Avg. score on negative class | missing_msrt | better | 1.28e-34 | 0.042 |
| AUROC ↑ | no_msrt | better | 1.28e-34 | 0.056 |
| AUROC ↑ | missing_msrt | better | 1.32e-34 | 0.031 |
| AUPRC ↑ | no_msrt | worse | 1.28e-34 | 0.175 |
| AUPRC ↑ | missing_msrt | worse | 1.28e-34 | 0.155 |
| Corrected AUPRC ↑ | no_msrt | better | 1.28e-34 | 0.191 |
| Corrected AUPRC ↑ | missing_msrt | better | 1.44e-34 | 0.086 |

### 7. Glossary

#### 7.1. General concepts

##### Event

Failure or more generally health condition that the model aims to predict. We assume that it has some duration.

##### Grouping / Group name

This refers to an attribute used to form the cohorts of patients.

##### Category

(abbreviation: **Cat**.) This refers to the value taken by the grouping attribute, it characterizes a specific cohort. It can also be used to directly designate a cohort.

##### Cohort

This is used to designate a particular category of patients (i.e. a set of patients that share a common grouping attribute value).

##### Macro-average

Consider a grouping with *n* categories, and each category *i* has a metric value *m_i*, then the macro-average is *(m_1 + m_2 +* … *+ m_n)/n*.

##### Delta

(abbreviation: Δ) Each stage is associated with certain metrics, the delta for a metric and a cohort corresponds to the absolute difference in median metric between patients of this cohort and the rest of the patients.

##### Threshold on score

Binary classifier outputs probability between 0 and 1, to obtain a binary output the user has to decide on a threshold value below which the output class will be 0 and above which it will be 1.

#### 7.2. Model Performance Analysis concepts

##### Metrics Definitions

**P** number of positive labels, **N** number of negative labels, **TP** number of correctly predicted positive labels, **TN** number of correctly predicted negative labels, **FP** number of instances with true negative labels but that were incorrectly predicted as positive by the model, **FN** number of instances with true positive labels but that were incorrectly predicted as negative by the model.

↑: Means that the larger the metric value, the better it is.

↓: Means that the lower the metric value, the better it is.

##### Recall

*TP/P*

##### Precision

*TP/(TP+FP)*

##### NPV

Negative predictive value, *TN/(TN+FN)*

##### FPR

False positive rate, *FP/(FP+TN)*

##### Corrected precision

Precision corrected for the cohort prevalence of positive labels, *TP/(TP+s*FP)* with *s* the correcting factor that depends on the cohort prevalence and the maximum prevalence for the grouping.

##### Corrected NPV

NPV corrected for the cohort prevalence of positive labels, *TN/(TN+s*FN)* with *s* the correcting factor that depends on the cohort prevalence and the minimum prevalence for the grouping.

##### Event-based recall

Number of detected events over the total number of events.

##### Calibration curve

Illustrates how well the probabilistic predictions of the model are calibrated (whether they can be interpreted as true probabilities), x-axis mean predicted probabilities, y-axis frequency of positive labels. The perfect calibration line (dashed line in the figures) acts as a reference.

##### Calibration error

Area between the calibration curve and the perfect calibration line.

##### Avg. score on positive class

for all positive labels, average of the output scores.

##### Avg. score on negative class

for all negative labels, average of the output scores.

##### ROC curve

Receiver operating characteristic curve, x-axis FPR, y-axis TPR.

##### AUROC

Area under the ROC curve.

##### PR curve

Precision-recall curve, x-axis recall, y-axis precision. It can be drawn also for event-based recall and corrected precision.

##### AUPRC

Area under the PR curve. It can be computed for the PR curve drawn with event-based recall and/or corrected precision.

##### Ratio of significantly worse metrics

For a specific category of patients, it refers to the number of metrics for which the category is significantly worse off compared to the rest of the population divided by the total number of metrics.

##### Worst ratio

Refers to the largest **ratio of significantly worse metrics** (for a grouping or for the overall analysis).

##### Worst delta

Refers to the largest **delta** in performance metrics (for a grouping or for the overall analysis).

#### 7.3. Time Gap Analysis concepts

##### Time gap

Amount of time between the trigger of the first correct alarm and the event occurence.

##### Start event

Considered split of the alarm horizon. We split the alarm horizon into different windows (chosen by the user) based on how much time in advance the alarm can be triggered. The available prediction horizon can not be longer than the time between the start of the considered event and the start of the stay or between the start of the considered event and the time when the previous event finished.

#### 7.4. Medical Variable Analysis concepts

##### Not in event

Refers to the median value computed on time points when patients aren’t undergoing an event.

##### Never in event

Refers to the median value computed for patients without any event during their stay.

#### 7.5. Feature Importance Analysis concepts

##### Feature importance

Approximates how useful is a feature for the prediction task. We use SHAP values to estimate it.

##### RBO (Rank-biased overlap)

Similarity measure between two lists that focuses more on the head of the list (i.e it penalizes more mismatches that occur at the beginning). We use this measure to compare two feature rankings.

##### General feature ranking

Refers to the ranking of features based on their importance (from the most important to the least important), obtained on the entire set of patients. In contrast to cohort-based rankings, that are obtained on a specific cohort of patients.

##### Delta of inverse rank

For a feature that has rank *rk_0* in the cohort-based ranking and *rk_all* in the general ranking, it is defined as |*1/rk_0 - 1/rk_all*|. If it is big enough, we consider the change in rank of the feature from the general to the cohort-based ranking to be significant.

##### Top 15 (cohort)

refers to the first 15 features of the general (or cohort-based) ranking.

#### 7.6. Missingness Analysis concepts

##### Performance metrics definitions

All metrics have already been defined in the **Model Performance Analysis concepts**.

##### Intensity of measurement categories

*no_msrt*: Refers to patients without any measurement for a variable.

*insufficient*: Refers to patients with between 0% (not included) and 90% of valid measurements (over the number of expected measurements).

The number of expected measurements is computed from the medical variable’s expected sampling interval *t_e* (input from the user) and the patient’s length of stay *los* as *los* / *t_e*.

*enough*: Refers to patients with between 90% (not included) and 100% of valid measurements (over the number of expected measurements).

The number of expected measurements is computed from the medical variable’s expected sampling interval *t_e* (input from the user) and the patient’s length of stay *los* as *los* / *t_e*.

##### Missingness categories

*no_msrt*: Refers to patients without any measurement for a variable (before full data imputation). *missing_msrt*: Refers to data points without valid measurement for a variable (before full data imputation but after forward propagation of measurements based on the variable’s expected sampling interval).

*with_msrt*: Refers to data points with valid measurements for a variable (before full data imputation but after forward propagation of measurements based on the variable’s expected sampling interval).

##### Dependent/Independent

Refers to the result of the Chi-squared independence test.

## Footnotes

1. https://europarl.europa.eu/doceo/document/TA-9-2022-0140_EN.html

2. https://data.consilium.europa.eu/doc/document/ST-15698-2022-INIT/EN/pdf

